# A qualitative meta-synthesis of service users’ and carers’ experiences of assessment and involuntary hospital admissions under mental health legislations: a five-year update

**DOI:** 10.1101/2024.03.27.24304909

**Authors:** Gergely Bartl, Ruth Stuart, Nafiso Ahmed, Katherine Saunders, Sofia Loizou, Grainne Brady, Hannah Gray, Andrew Grundy, Tamar Jeynes, Patrick Nyikavaranda, Karen Persaud, Ari Raad, Una Foye, Alan Simpson, Sonia Johnson, Brynmor Lloyd-Evans

## Abstract

**Background:** Compulsory admissions occur in psychiatric hospitals around the world. They result in coercive and sometimes traumatic experiences for service users and carers. Legal and service reforms in various countries are intended to reduce rates of detention and improve service user experience. We aimed to inform policy and service delivery by providing an up-to-date synthesis of qualitative evidence on service users’ and carers’ experiences of assessment and detention under mental health legislation, updating previous reviews in which we searched for literature published up to 2018.

**Methods:** We searched five bibliographic databases for studies published between January 2018 and March 2023. We identified 24 additional studies reporting qualitative investigations of service users’ or carers’ experiences of assessment or detention under mental health legislation. A team including researchers with relevant personal experience analysed and synthesised data using a thematic synthesis approach.

**Results:** Findings suggest that views on compulsory admissions and assessment varied: many reports highlighted its often negative, traumatic impacts on emotional well-being and self-worth, with fewer accounts of it as an opportunity to access help and support, accompanied by feelings of relief. Experiences of racial discrimination, inequality of access, and dissatisfaction with support before and after hospital stay were more prominent than in our previous reviews.

**Conclusions:** Increasing service user and carer involvement in treatment decisions, provision of timely information at key stages of the admission process, training of key personnel, addressing the issue of discrimination, and investing in community alternatives of inpatient care may contribute to and lead to better overall treatment experiences.

## Background

Involuntary treatment and compulsory hospital admissions are commonly used in psychiatric services in many countries, and present service users, carers, medical professionals, and legislators with a range of emotional, ethical and practical challenges (1, 2). There are large but at times unexplained variations between countries in compulsory admission rates, and rates have risen in some countries including England in recent years despite policies expanding community care (3, 4).

Whilst legislation requires involuntary admissions to be based on assessment of clinical acuity and risks, the decision-making process appears to be affected by several other factors. For example, variations in involuntary admission rates have been linked to service level characteristics including mental health legislation, availability of inpatient services, community-based alternatives, or public attitudes towards mental illness (5, 6, 7). Some individual characteristics also seem to increase the risk of involuntary admissions, including diagnosis, gender, age, ethnicity and race, migrant status, treatment received before admission and contact with police and legal system (8, 9, 10, 11). Previous decisions in an individual’s care are also likely to influence future assessments, e.g. making decisions that colleagues have made previously may feel safer for detaining clinicians (11).

Involuntary admissions remain a complex process to navigate for service users and staff, where service users may find it difficult to exercise their rights and express preferences due to being in a ward environment, staff attitudes, or lack of communication (12). Admissions are often associated by experiences of coercion (13), which in turn affects service users’ attitudes towards psychiatric treatment (14). Thus, compulsory admissions present a challenging situation where service users’ well-being, as well as freedom and rights need to be taken into account and respected (15) in addition to clinical considerations.

At the same time, detention is thought to lead to only modest improvements in clinical and social outcomes (16). Service users who undergo compulsory admission (compared to those who are admitted voluntarily) have higher rates of suicide and greater dissatisfaction with care, as well as increased risk of readmission, especially compulsory admission (17). Rates of service users’ agreement with the decision to detain them vary internationally, with more than half of patients in some countries still disagreeing with the decision to detain them three months retrospectively (18). Experiences of coercion vary substantially both for voluntary and detained patients (19). It is therefore important to understand service users’ experience and what contributes to particularly negative experiences of detention, to inform efforts to improve detained service users’ experience in future. As one contributor to the call for evidence to the 2018 Independent Review of the Mental Health Act in England put it:

> *‘I can understand looking back why I needed to be detained at that moment in my life, but what I can’t understand is why it was such a needlessly unpleasant experience’* (*20*).

Disproportionally high rates of involuntary admissions of people from marginalised groups is also an increasingly recognised issue, reported internationally (8), including in the UK and US (21, 22). In England, people from Black ethnic groups are 3.5 times more likely to be detained than White British people (23).

In our two previous reviews (24, 25) we reported findings from papers published in or before 2017 which explored service users’ and carers’ experiences of compulsory treatment and hospital admissions in psychiatric care. These reviews highlighted gaps in research on experiences of the assessment process for compulsory admission, and experiences of people from a minority ethnic background. Therefore, we aimed to carry out an update of the two previously published systematic reviews on service user and carer experiences of compulsory admissions, to understand these experiences in contemporary mental health contexts and provide an updated evidence synthesis. Using the new studies over the last 5 years, including those with a focus on specific groups (e.g. marginalised groups, forensic services) and broadening our search to also include children and young people (under 18s), we carried out a synthesis of evidence to address two research questions: 1) what are service users’ experiences of being formally assessed for involuntary hospital admission and/or legal detention in a psychiatric hospital, and 2) what are carers’ experiences of the formal assessment of their family and friends for involuntary admission and/or legal detention in a psychiatric hospital?

## Methods

### Protocol and registration

We prospectively registered the study protocol in the publicly accessible PROSPERO database on 30^th^ May 2023 (CRD42023423439). The protocol differed from our previous reviews (24, 25) by including service users under 18 and their family carers. We adhered to the updated PRISMA reporting principles (26) and included the completed PRISMA Checklist 2020 in Supplementary Material.

### Eligibility Criteria

Inclusion criteria were peer-reviewed qualitative studies published in any language from 1^st^ January 2018. We excluded books, systematic reviews, commentaries, editorials or grey literature such as pre-prints, dissertations, PhD theses, government reports, conference abstracts. We included studies that collected data through interviews or focus groups as well as auto-ethnographic or case studies. Studies that collected data using questionnaires or surveys were excluded. No restrictions were placed on language or country of study.

There were no age limits for participants. We included papers with service users who had been assessed for compulsory admission or legally detained to a psychiatric hospital and unpaid carers who provided practical and emotional support to such service users. Paid carers and other professionals were excluded.

Key outcomes for which data were sought were service users’ or informal carers’ experiences of either (a) assessment for compulsory admission to a psychiatric hospital, (b) legal detention in a psychiatric hospital and/or (c) appeal and tribunal processes related to detention in a psychiatric hospital. We excluded studies reporting experiences of voluntary and involuntary admission which did not separate these conditions in analyses, and studies which only reported involuntary outpatient treatment in the community.

### Data sources

We searched five electronic databases (MEDLINE, PsycINFO, HMIC and Embase via the Ovid platform, and the Social Sciences Citation Index via the Web of Science platform) for articles published between 1st January 2018 and 1st March 2023 (see the search strategy for each database in Supplementary Materials). In addition, we screened the reference lists of all eligible papers for other relevant studies and carried out a forward citation search on all included papers and our previous reviews (24, 25) using Web of Science. Manual screening was completed on 6th July 2023.

### Selection process

The resulting list of records was deduplicated in EndNote software before being imported into Rayyan (27) for independent screening. We piloted the inclusion and exclusion criteria on 10 titles and abstracts. All others were then screened independently by one reviewer. A second reviewer double-screened some of each reviewers’ allocation of titles and abstracts, 10% in total. Any discrepancies were resolved between the reviewers. Potentially eligible, full articles were subsequently reviewed independently by two reviewers. Uncertainties were resolved by discussion and in some cases by consultation with a third reviewer.

### Data extraction

We adapted the data extraction table developed in the previous reviews for use in the current review. Data extraction was piloted and revised based on 10% included papers using Microsoft Excel. Data extraction was carried out by one member of the research team and checked for accuracy by another researcher. Extracted information included study authors, year of publication, study focus (service user, carer, or both), study setting (country and whether single-site or multi-site), total sample, gender and/or sex, age-range, ethnicity, diagnosis, method of data collection, and method of analysis. We summarised main themes identified by the authors of papers at this stage, prior to meta-synthesis of papers.

### Risk of bias and quality appraisal

We used the Critical Appraisal Skills Programme (28) qualitative checklist to appraise the quality of included studies. This included an appraisal of papers along several domains covering research ethics, suitability of methods, recruitment and data collection processes, data analysis, reflection on sources of bias, clarity and validity of findings. Two reviewers conducted quality appraisal independently and discrepancies were resolved through discussion.

### Assessment of Confidence in Findings

Two reviewers independently assessed certainty of evidence for each individual finding, defined as the sub-themes, using an adapted GRADE Confidence in the Evidence from Reviews of Qualitative research approach (GRADE-CERQual, (29)). We operationalised components of the GRADE-CERQual to the context of the review as described in Supplementary Materials.

### Data analysis and synthesis

We analysed findings (‘Results’ or ‘Findings’ sections extracted verbatim) from included studies using a two-stage process, informed by Thematic Synthesis (30). In stage one, we developed a separate service user and carer initial coding frame to reflect the over-arching themes developed through thematic synthesis and reported in the two previous reviews (24, 25). Following discussion and agreement by the team, we defined and added to the coding frames sub-themes implicit in the published reviews. We tested and refined the two coding frameworks and coding process by double-coding and reviewing two service user and two carer papers. Thereafter, we adopted a deductive approach to code findings from all included studies to the overarching themes. In stage two, the wider project team reviewed relevant data which could not be satisfactorily coded to existing themes and developed additional themes inductively and iteratively. We then modified the initial framework to accommodate newly identified themes and sub-themes, where necessary. All coding was carried out in NVivo 14 (31).

We planned to analyse and report findings regarding service users’ and carers’ experiences’ separately. No formal comparison of sub-groups was planned, but we identified and specifically considered findings regarding groups of people and stages of the detention process for which evidence was limited in our previous reviews (24, 25). These included: young people, people from minority ethnic communities, and the process of assessment for compulsory admission.

### Reflexivity

Our research team included people of different backgrounds with prior interest and experience in the topic of investigation. It included senior and junior researchers, people with experience of delivering and receiving mental health care, and people with different ethnic and cultural backgrounds. The team included authors with practitioner experience (psychiatry, nursing and social work), and authors with personal experience of undergoing compulsory detention, or caring for someone undergoing compulsory detention. The team met regularly to plan the study, review study screening and inclusion decisions, contribute to analysis and decisions about themes, and to comment on drafts of the paper – and thus utilised the variety of perspectives to inform the study.

## Results

### Search results

We report the number of studies identified, screened, and included/excluded through the database search and citation searches in the PRISMA Flow Diagram in Figure 1. A total of 24 eligible papers were identified. Sixteen of these reported experiences of service users only, three of them of carers only. There were five additional articles that contained data regarding both groups. We report study characteristics and our thematic synthesis separately below: first from papers reporting service users’, then carers’ experiences. Papers which reported on both groups are included in both tables, with their relevant characteristics (e.g. number of service user, or carer participants) indicated where appropriate. The quality appraisal of papers is reported in Table 1. Twenty-three studies were rated as high quality, i.e. scored 7 or more on the CASP tool, and one study was rated as medium quality, scoring 4 or above, but below 7.

**Figure 1.**
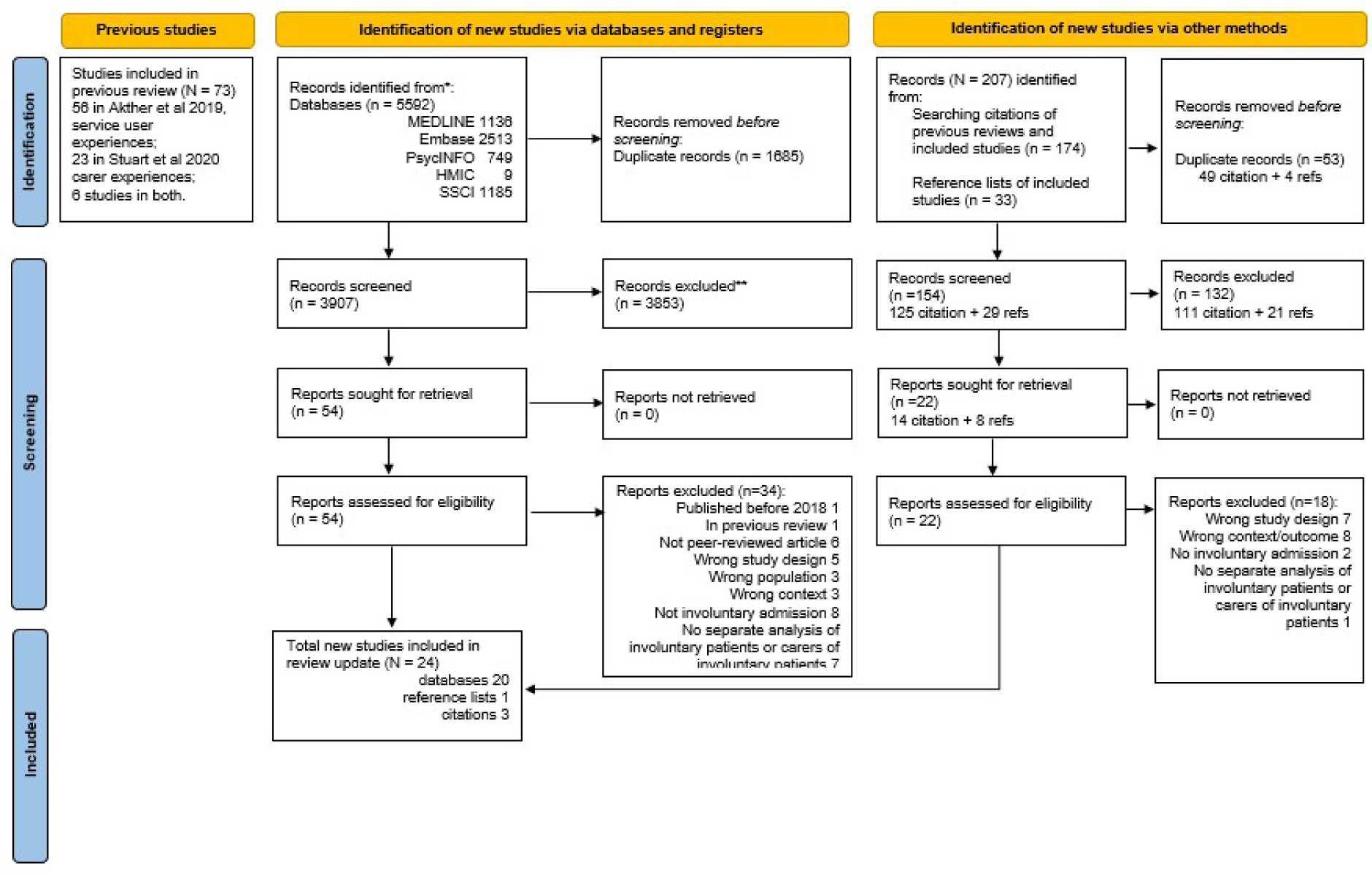
PRISMA Flow Diagram

**Table 1.** Quality appraisal of included studies.

| Quality criteria | Aluh et al. (2022) | Bendelow et al. (2019) | Blakley et al. (2022) | Dixon et al. (2022) | Goodall et al. (2019) | Jaeger et al. (2019) | Jones et al. (2021a) | Jones et al. (2021b) | Kalagi et al. (2018) | Lawrence et al. (2019) | MacDonald et al. (2023) | McDonnaugh et al. (2020) | McGuiness et al. (2018) | O'Connor et al. (2021) | Pothoff et al. (2022) | Ranieri et al. (2018) | Smyth et al. (2021) | Solanki et al. (2023) | Sondhi et al. (2018) | Vallarino et al. (2019) | Verstegen et al. (2022) | Wormdahl et al. (2021) | Wyder et al. (2018) | Yu et al. (2022) |
| --- | --- | --- | --- | --- | --- | --- | --- | --- | --- | --- | --- | --- | --- | --- | --- | --- | --- | --- | --- | --- | --- | --- | --- | --- |
| 1. Was there a clear statement of the aims of the research? | Y | Y | Y | Y | Y | Y | Y | Y | Y | Y | Y | Y | Y | Y | Y | Y | Y | Y | Y | Y | Y | Y | Y | Y |
| 2. Is a qualitative methodology appropriate? | Y | Y | Y | Y | Y | Y | Y | Y | Y | Y | Y | Y | Y | Y | Y | Y | Y | Y | Y | Y | Y | Y | Y | Y |
| 3. Was the research design appropriate to address the aims of the research? | Y | Y | Y | Y | Y | Y | Y | Y | Y | Y | Y | Y | Y | Y | Y | Y | Y | Y | Y | Y | Y | Y | Y | Y |
| 4. Was the recruitment strategy appropriate to the aims of the research? | Y | Y | Y | Y | Y | - | Y | Y | - | Y | Y | Y | Y | Y | Y | Y | Y | Y | Y | Y | Y | Y | Y | - |
| 5. Was the data collected in a way that addressed the research issue? | Y | Y | Y | Y | - | Y | Y | Y | Y | Y | Y | Y | Y | Y | Y | Y | Y | Y | Y | Y | Y | Y | Y | Y |
| 6. Has the relationship between researcher and participants been adequately considered? | Y | Y | Y | Y | N | - | Y | Y | Y | Y | Y | Y | Y | Y | Y | N | - | Y | - | - | - | - | - | - |
| 7. Have ethical issues been taken into consideration? | Y | Y | Y | Y | Y | Y | Y | Y | Y | Y | - | Y | Y | Y | Y | Y | Y | Y | Y | Y | Y | Y | Y | Y |
| 8. Was the analysis sufficiently rigorous? | Y | - | Y | Y | Y | Y | Y | Y | Y | N | Y | Y | Y | Y | Y | N | - | Y | - | Y | Y | Y | Y | - |
| 9. Is there a clear statement of findings? | Y | Y | Y | Y | Y | Y | Y | Y | Y | N | Y | Y | Y | Y | Y | Y | Y | Y | Y | Y | Y | Y | Y | Y |
| Number of criteria met: | 9 | 8 | 9 | 9 | 7 | 7 | 9 | 9 | 8 | 7 | 8 | 9 | 9 | 9 | 9 | 7 | 7 | 9 | 7 | 8 | 8 | 8 | 8 | 6 |

Criteria met: Yes (Y), No (N), Can't tell (-)

### Service users

#### Overview of included studies

Key characteristics of included service user papers are reported in Table 2. Most studies were conducted in Europe (UK: 7, Germany: 2, Ireland: 2, Denmark, Italy, Netherlands, Norway 1 each), whilst three were based in the US (Florida and New York) and one each in Australia, Nigeria and South Korea. All studies were published in English.

**Table 2.** Key characteristics of included studies: service user experiences.

| Authors | Study Focus | Sample size | Gender (female/male/non-binary) | Age in years Range or Mean | Ethnicity | Diagnoses of patients | Method of data collection | Method of data analysis | Lived experience involvement |
| --- | --- | --- | --- | --- | --- | --- | --- | --- | --- |
| Aluh et al., 2022<br>Nigeria, multi-site | To investigate service users' perceptions and experiences of coercive practices in Nigerian psychiatric hospitals. | 30 | 11/19/0 | M=34.67 | NR | Schizophrenia (12), Schizoaffective disorder (2), Bipolar disorder (3), Depression (3), Mental & Behavioural Disorder due to psychoactive substance (10) | Focus groups | Thematic Analysis | None |
| Bendelow et al., 2019<br>Sussex, UK, single-site | To investigate the complexities underlying high rates of Section 136 detention in Sussex highlighted by Home Office review in 2014. | 34 | NR | NR (sub-sample 19-65) | NR | Complex histories of often multiple diagnoses, including Borderline or Emotionally Unstable Personality Disorder, Dissociative Identity Disorder, Bipolar Disorder and Complex Post-Traumatic Stress Disorder. (Exact numbers NR) | Semi-structured narrative interviews | Thematic Analysis | Patient involvement advisory group (input in analysis) |
| Blakley et al., 2022<br>England, UK, single-site | To explore the subjective experience of a Mental Health Act Assessment from service users' perspectives | 10 | 7/3/0* | 22-59 | 8 White, 2 Black Afro-Caribbean | NR | Structured interviews | Framework Analysis | Co-designed study, LE input in conception, interview schedule, pilot and analysis. |
| Goodall et al., 2019<br>England, UK, multi-site | To gain insight and understanding of the process of being detained under S136 of the Mental Health Act from the detained person's perspective. To identify critical factors that helped or worsened the experience from the service user's perspective. Explore of wish list of potential improvements. | 15 | 2/13/0 | 18-64 | NR | NR | Semi-structured interviews | Critical Incident Technique | Participants invited to check interpretations reflect their lived experience. |
| Jaeger et al., 2019<br>Germany, multi-site | To explore perspectives of patients service users who currently or previously refused antipsychotic medication during inpatient treatment, their family members, physicians, and nursing staff during an interim period in Germany when, due to legislative change, involuntary medication of patients was not regulated and thus not available as an option. | 11 | NR/6/NR | 25-60 | NR | Schizophrenia Spectrum Disorder (82%), Affective Disorder (18%) | Guideline-based, problem-centred interviews | Grounded Theory | NR |
| Jones et al., 2021<br>Florida, US, multi-site | To investigate how initial involuntary hospitalizations impact youth and young adult treatment pathways following discharge | 40 | 28/NR/1 | 16-27 | 18 White, 4 African-American, 5 Asian-American, 12 Latinx, 1 Multiracial | NR | Semi-structured interviews | Grounded Theory | Broader team included service users with LE of involuntary admissions, e.g. in interpretation. |
| Jones et al., 2022<br>Florida, US, multi-site | To better understand youths' and young adults' experiences of police involvement in situations in which police officers served as first responders to a psychiatric crisis. | 40 | 20/NR/1 | 16-27 | 11 White, 3 Black, Asian American, 11 Latinx, 1 Multiracial | NR | Semi-structured interviews | Grounded Theory and Thematic Analysis | LE members of research team. |
| Kalagi et al., 2018<br>Germany, single-site | To assess the opinions and values of relevant stakeholders with regard to the requirements for implementing open wards in psychiatric hospitals. | 15 | 3/12/0 | 20-60 | NR | Psychotic Disorder (6), Affective Disorder (6), Substance dependence (3) | Semi-structured interviews | Qualitative Content Analysis | NR |
| Lawrence et al., 2019<br>New York, US, multi-site | To understand how experiences with coercion, e.g. involuntary hospitalisation and locked facilities, may affect the inpatient treatment alliance. | 50 | 19/31/0 | 23-88 | 22 White, 14 Black, 11 Hispanic, 3 Other | Schizophrenia (6), Schizoaffective disorder (6), Substance-induced psychotic disorder (2), Major depression (25), Bipolar disorder (11) | Semi-structured interviews | Grounded Theory | NR |
| MacDonald et al., 2023<br>Denmark, multi-site | To explore the experiences and perspectives of anorexia nervosa patients with regard to involuntary detention with an exclusive focus on multiple IT events, to enhance our understanding of IT and potentially inform treatment | 7 | 7/0/0 | 20s to 30s | NR | Anorexia, Nervosa. Comorbid psychiatric diagnoses (6): Personality Disorders, Depression, ASD, Obsessive Compulsive Disorder, ADHD, and Schizotypal Disorder | Semi-structured interviews | Reflexive Thematic Analysis | NR |
| McDonnau gh et al., 2020<br>UK, multi-site | To explore the experiences of mentally disordered male offenders conditionally discharged from secure hospitals on a restrictive Section of the Mental Health Act (Section 37/41). | 7 | 0/7/0 | 28-44 | 6 White British, 1 Black Caribbean British | NR | Semi-structured interviews | Thematic analysis | NR |
| McGuinness et al., 2018<br>Ireland, multi-site | To develop an understanding of individuals' experiences over the course of the involuntary admission process. | 50 | 21/29/0 | 23-85 | NR | Bipolar disorder (14), Schizophrenia (13), Schizoaffective disorder (10), Alcohol Dependence Syndrome (3), Recurrent Depressive Disorder (2), Acute and Transient Psychotic Disorder (2), Schizophreniform Psychosis (1), Substance-Induced Psychotic Disorder (1), and | Semi-structured interviews | Grounded Theory | Reference to self-experience influencing topic guide design. |
|  |  |  |  |  |  | Anorexia Nervosa (1), and Other (3) |  |  |  |
| O'Connor et al., 2021<br>Australia, single-site | To explore service user experiences of a forensic service that endorsed a recovery model | 8 | 3/5/0 | 30–51 | NR | Schizophrenia (7), Schizophrenia and Borderline Personality Disorder/Schizoaffective Disorder (1) | Semi-structured interviews | Thematic Analysis | NR |
| Potthoff et al., 2022<br>Germany, multi-site | To develop a conceptual model of psychological pressure based on the perspectives of service users | 14 | 7/7/0 | 21-63 | NR | Psychotic Disorders (6), Affective Disorders (6), Substance Dependence (1), Personality Disorder (1) | Semi-structured interviews | Grounded Theory | NR |
| Smyth et al., 2021<br>Ireland, multi-site | To examine and compare retrospective qualitative perceptions of service-users in relation to their involuntary admission with their levels of clinical insight, using a mixed methods approach | 42 | 20/22/0 | 18-24 to 65+ | NR | Bipolar disorder (14), Paranoid Schizophrenia (12), Schizo-Affective Disorder (9), Acute and Transient Psychotic Disorder (2), Schizophreniform Psychosis (1), Substance Induced Psychotic Disorder (1), Major Depressive Disorder (1), Anorexia Nervosa (1), Other (1) | Semi-structured interviews | Content Analysis | NR |
| Solanki et al., 2023<br>England, UK, single-site | To explore the lived experiences of adults from a Black ethnic background who have been detained as inpatients under the MHA. | 12 | 8/4/0 | 18-60 | 2 Black African, 1 Black African/Black Caribbean/Black Other, 3 Black British, 2 Black Other, 1 Caribbean, 1 Caribbean and African, 1 Mixed Race, 1 Black British/Black African | Acute Stress Disorder (1), Paranoid Schizophrenia (1), Psychosis (4), Schizophrenia (1), Split Personality (1), Don't know (4) | Semi-structured interviews | Thematic Analysis | NR. Feedback sought from one participant with LE. |
| Sondhi et al., 2018<br>London, UK, multi-site | To describe the views and perceptions of the process for people with lived experience of mental distress who have been detained under Section 136 of the Mental Health Act 1983. | 54 | 40/NR/NR | M=44.1 | white 52 (of whole sample of 58 inc. 4 carers) | NR | Semi-structured interviews | Grounded Theory and Thematic Analysis | Service user group (pilot, interview guide) |
| Vallarino et al., 2018<br>Milan, ITA, single-site | To explore the experiences of mental health care in a sample of Italian adults with BD, to gain a broad understanding of their expectations, views and evaluations of key aspects of services received. (Involuntary experiences sub-sample) | 9 | NR | 37-56 | NR | Bipolar Disorder | Unstructured interviews | Thematic Analysis | NR |
| Verstegen et al., 2022<br>Netherlands, single-site | To increase insight into patient experiences with victimization during clinical forensic psychiatric treatment | 9 | 2/7/0 | 28-51 | NR | NR (aimed to include schizophrenia spectrum disorders and cluster B personality disorders) | Semi-structured interviews | Grounded Theory approach | NR |
| Wormdahl et al., 2021<br>Norway, multi-site | To explore the characteristics of the paths toward referral to involuntary psychiatric admission of adults with severe mental illness. | 16 | NR | NR | NR | NR | Focus groups (SU&C) | Grounded Theory | Peer researcher involved in design, data collection, analysis. |
| Yu et al., 2022<br>South Korea, multi-site | To explore the perspectives of persons with mental illness and their family members who had first-hand experience with involuntary admission after the revision of the Mental Health and Welfare Act 2016 and investigate whether the MHWFA 2016 has improved the human rights of persons with mental illness, based on their experiences. | 7 | 2/5/0 | 30s-60s | NR | Schizophrenia (5), Bipolar Disorder (2) | Semi-structured interviews | Thematic Analysis | NR |

Sample sizes varied from 7 to 54 for service users; 13 of the 21 papers with service user experiences had a sample size of interest of 20 or less. The majority of service user papers reported on gender (n = 18), age (n = 19) and diagnosis (n = 14), but fewer reported ethnicity (n = 7). The samples included experiences from people with a variety of mental health diagnoses including but not limited to schizophrenia, depression, bipolar disorder, personality disorder.

Papers investigating service user perspectives focused on various aspects of the involuntary assessment and treatment process, e.g. looked specifically at assessment/referral experiences, admission under mental health legislation involving police, detention in general acute psychiatric hospital or forensic settings, and some had a focus on the experience of coercion, discharge or specifically the experiences of black ethnic service users in the UK.

#### Thematic synthesis

The review and update of the thematic framework for service user experiences has been carried out based on 21 identified papers (including 5 mixed papers on service user and carer reports). Most studies investigated either assessment or inpatient stay experience, with some focusing on more specific aspects or experiences, e.g. the nearest relative role, medication use, coercive practices, open-wards or discharge. There were two papers that looked in more detail at experiences of young people (32, 33), and one that specifically explored experiences of people from minoritised ethnic backgrounds (34). The corresponding, updated thematic framework including in-depth sub-theme descriptions as illustrated in Table 4. Main themes and subthemes (indicated in *italics*) are also summarised in the text below.

**Table 3.** Key characteristics of included studies: carer experiences.

| Authors/<br>Year of<br>publication/<br>Location | Study aim | Carer<br>participants /Total<br>sample<br>size | Carer<br>gender<br>female/<br>male | Relationship<br>to<br>person<br>cared for | Carer<br>Age<br>range | Carer<br>Ethnicity | Method<br>of data<br>collection<br>from<br>carers | Method<br>of data<br>analysis | Lived<br>experience<br>involvement |
| --- | --- | --- | --- | --- | --- | --- | --- | --- | --- |
| Bendelow, G.<br>Warrington, CA<br>Jones, AM<br>Markham, S<br>**2019**<br>Sussex, UK,<br>single site | To investigate the complexities underlying high rates of Section 136 detention in Sussex highlighted by Home Office review in 2014. | 3/ 62 | NR | Carers of services users who died from suicide. Carer relationship to service user not reported. | NR | NR | Semi-structured narrative interviews (face-to-face or over telephone) | Thematic Analysis | Patient involvement advisory group (input in analysis) |
| Dixon, J.<br>Stone, K.<br>Laing, J.<br>**2022**<br>England, UK,<br>multi-site | To discover how individuals experience the Nearest Relatives role when involved in a Mental Health Act assessment and what kind of supports may be needed to help them to exercise it appropriately. | 19 / 19 | 11 / 8 | 8 Mothers<br>1 Daughter<br>2 Wives<br>2 Fathers<br>1 Brother<br>2 Sons<br>3 Husband | 34-72 | 19 White | Semi-structured interviews | Thematic Analysis | NR |
| Jaeger, S.<br>Huther, F.<br>Steinert, T.<br>**2019**<br>Germany<br>multi-site | To explore perspectives of service users who currently or previously refused antipsychotic medication during inpatient treatment, their family members, physicians, and nursing staff during an interim period in Germany when, due to legislative change, involuntary medication of patients was not regulated and thus not available as an option. | 8 / 33 | 5 / 3 | 2 fathers<br>1 mother<br>1 sister<br>2 spouses<br>2 daughters | 19-72 | NR | Guideline-based, problem-centred interviews | Grounded Theory | NR |
| Ranieri, V.<br>Wilson, C.<br>Davidson, L.<br>**2018.**<br>Ireland,<br>Connecticut<br>USA,<br>multi-site | To provide a preliminary and exploratory post-discharge account of mental health caregivers' experiences of securing involuntary admission of a relative, particularly caregivers' perceptions of coercion surrounding the admission and events leading up to it, and how they interpret such perceptions after discharge. The secondary aim is to observe whether caregivers revealed different experiences depending on their location and legislation. | 14 / 14<br>(9 Ireland, 5 USA) | 14 / 0 | 13 mothers<br>1 daughter | >18 | NR | Semi-structured interviews | Thematic Analysis | NR |
| Sondhi, A.<br>Luger, L.<br>Toleikyte, L.<br>Williams, E.<br>**2018**<br>Greater London, UK, multi-site | To describe the views and perceptions of the process for people with lived experience of mental distress who have been detained under Section 136 of the Mental Health Act 1983. | 4 / 58 | reported for total sample, not separately for carers | NR | reported for total sample, not separately for carers | reported for total sample, not separately for carers | Semi-structured interviews | Grounded Theory and Thematic Analysis | Service user group (pilot, interview guide) |
| Wormdahl, I.<br>Husum, T.L.<br>Kjus, S.H.H.<br>Rugkasa, J.<br>Hatling, T.<br>Rise, M.B.<br>**2021**<br>Norway, multi-site, | To explore the characteristics of the paths toward referral to involuntary psychiatric admission of adults with severe mental illness. | 21 / 103 | reported for total sample, not separately for carers | parent, sibling, spouse (numbers not specified) | NR | NR | 2 interviews, 19 carers in focus groups with service users | Grounded Theory | Peer researcher (design, data collection, analysis) |
| Wyder, M.<br>Bland, R.<br>McCann, K.<br>Crompton, D.<br>**2018**<br>Brisbane, Queensland, Australia single site | To explore the family's experience of admission under an involuntary treatment order and the impact this had on their caring ability and wellbeing. | 19 / 19 | 12 / 7 | 6 fathers<br>9 mothers<br>3 partners<br>1 sibling | NR | NR | Semi-structured interviews | Inductive analysis (Strauss and Corbin, 1998) | NR |
| Yu, S. Y.<br>Heo, J.<br>Yoon, N. H. | To explore the perspectives of persons with | 3 / 10 | 3 / 0 | 2 mothers<br>1 peer worker | 50-79 | NR | Semi-structured interviews | Thematic Analysis | NR |
| Lee, M.<br>Shin, S.<br>2022<br>South Korea,<br>multi-site | mental illness and their family members who had first-hand experience with involuntary admission after the revision of the Mental Health and Welfare Act 2016 and investigate whether the MHWA 2016 has improved the human rights of persons with mental illness, based on their experiences. |  |  |  |  |  |  |  |  |

**Table 4.**
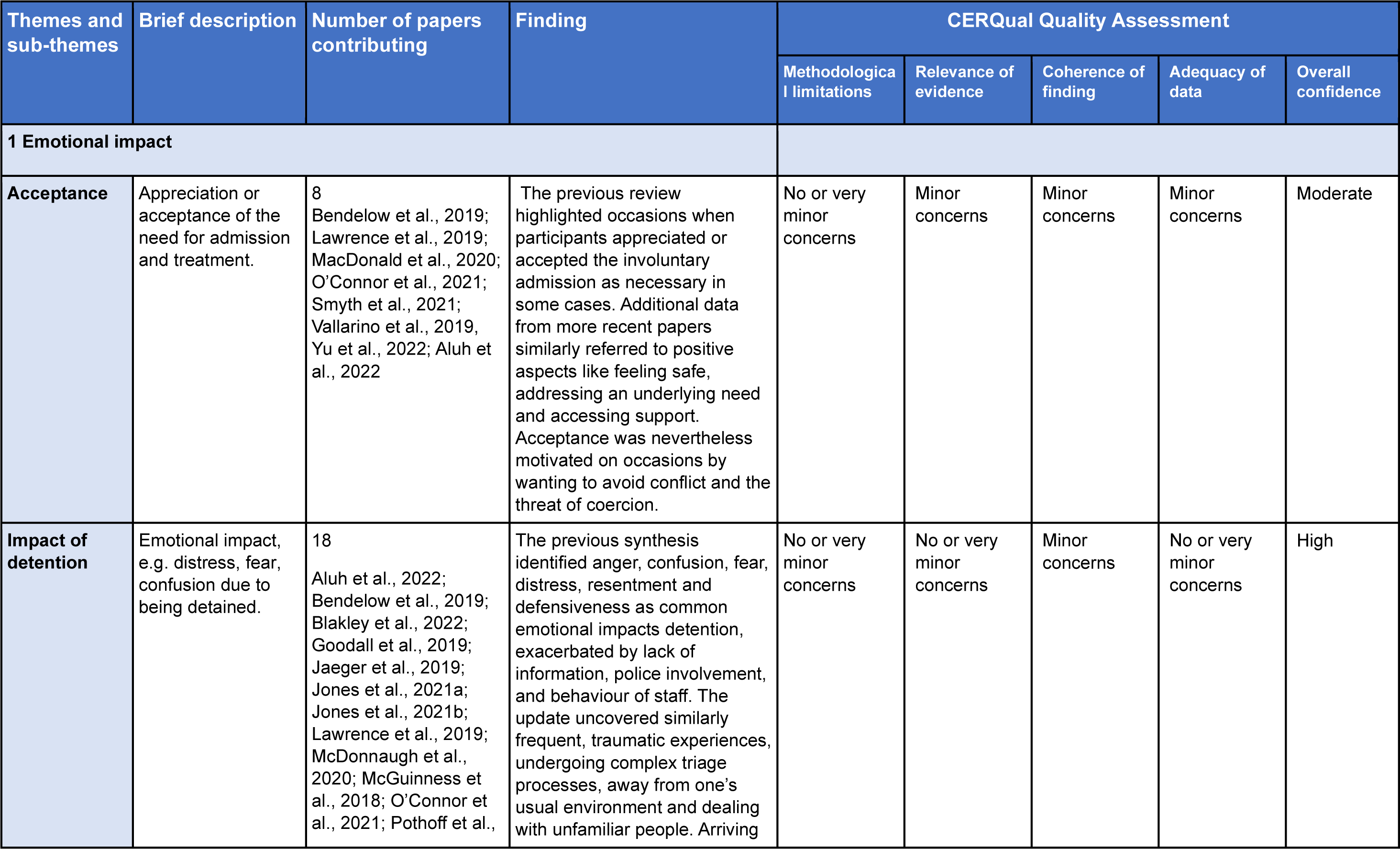
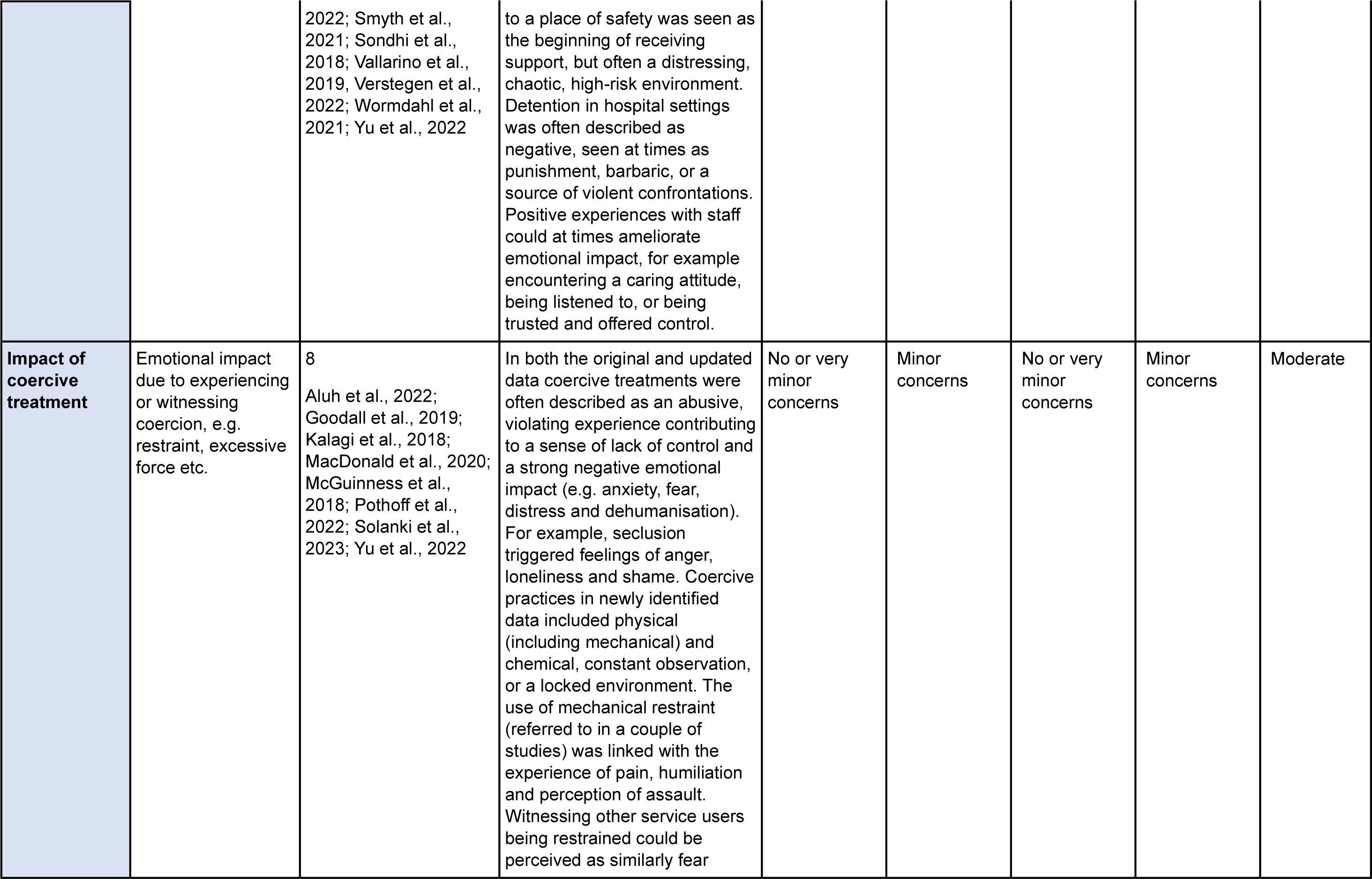
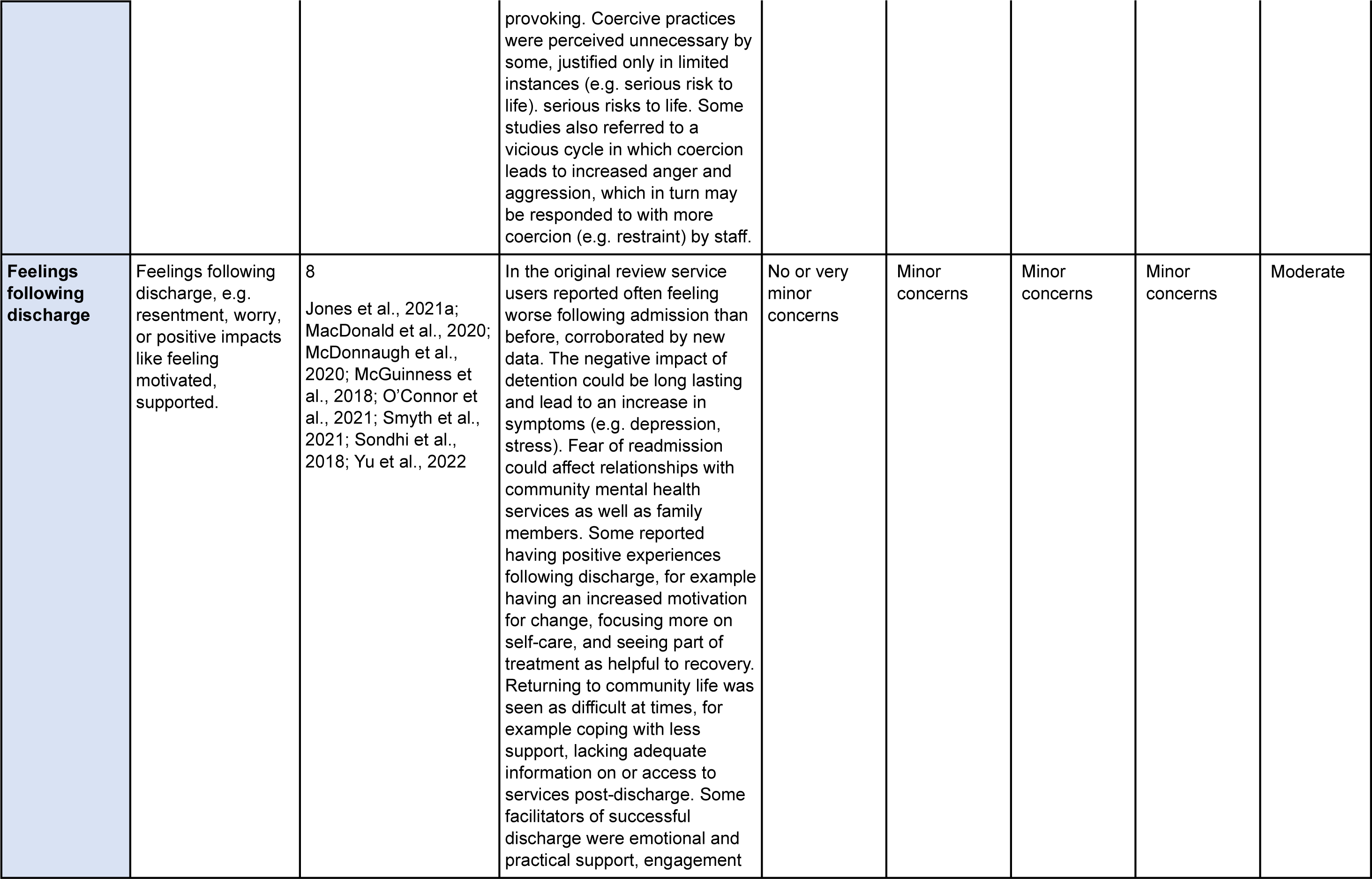
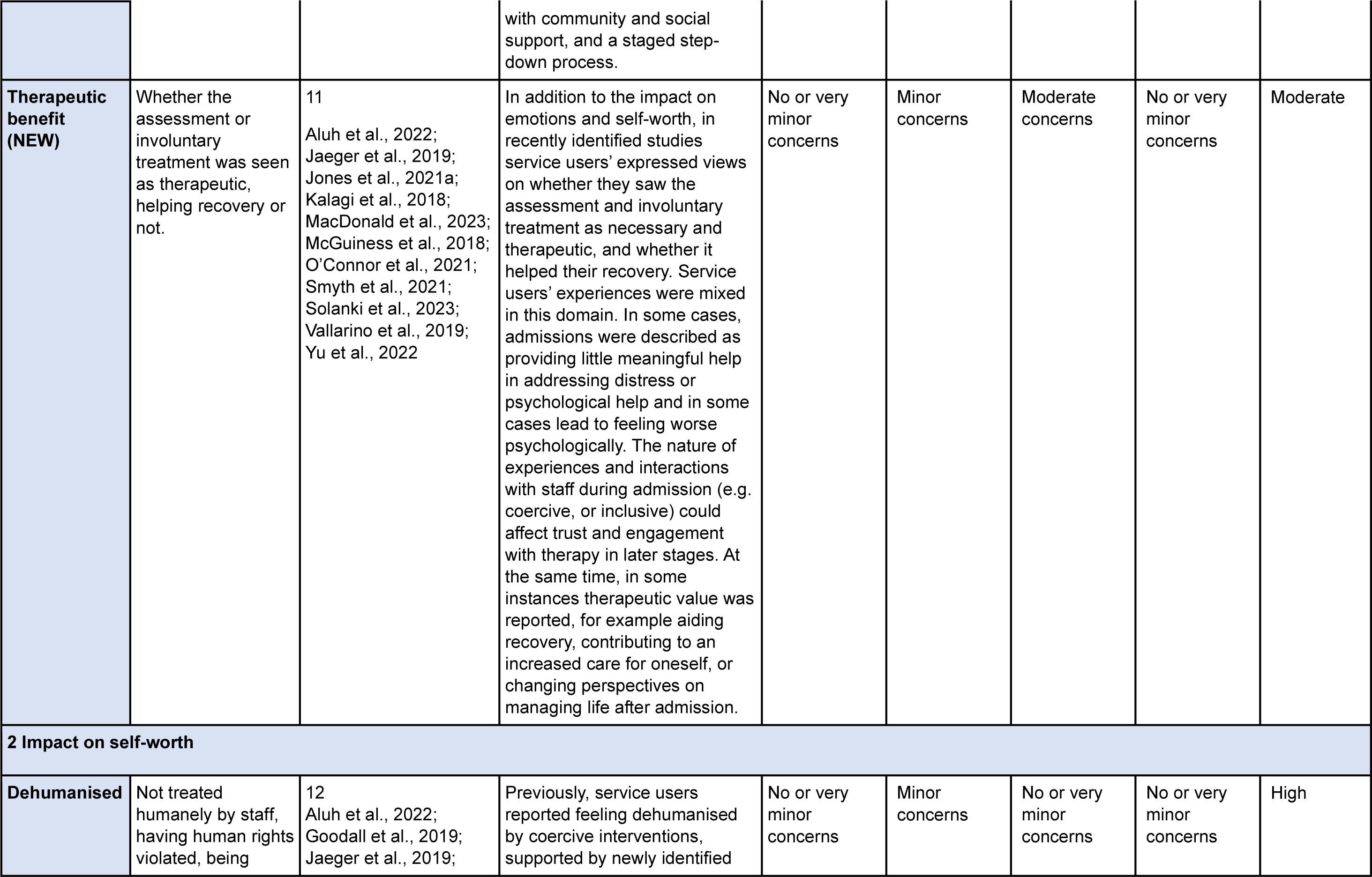
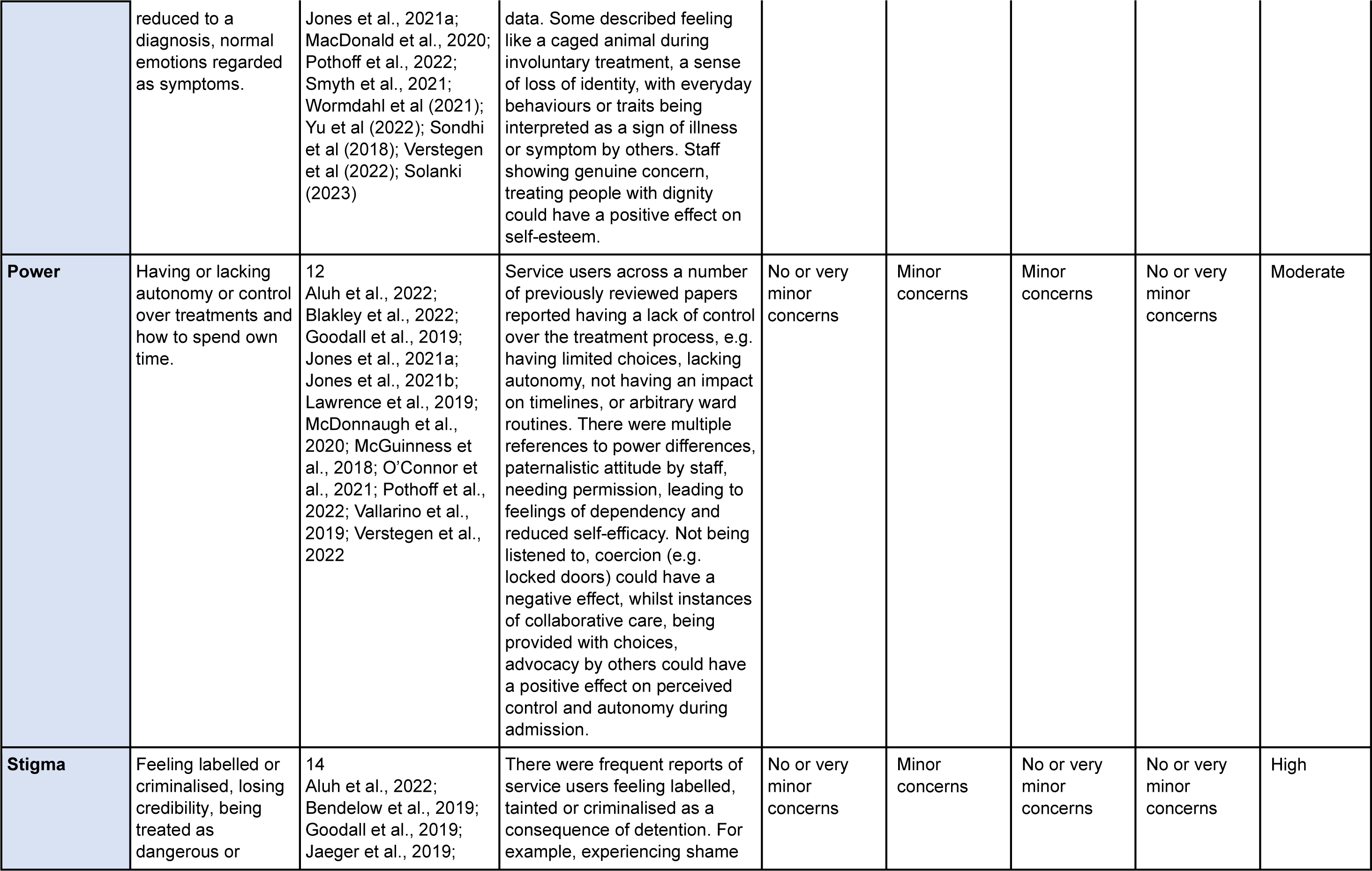
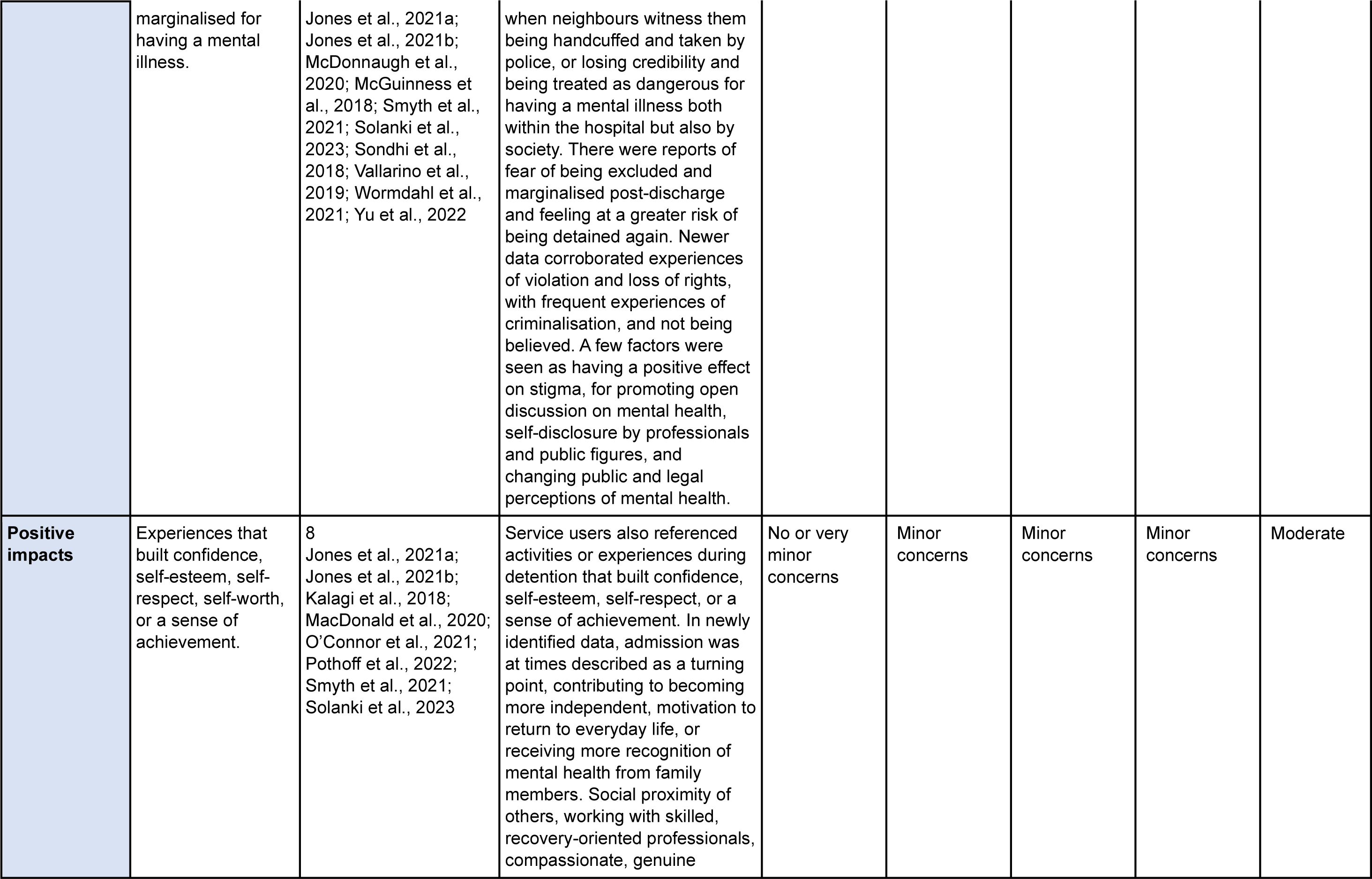
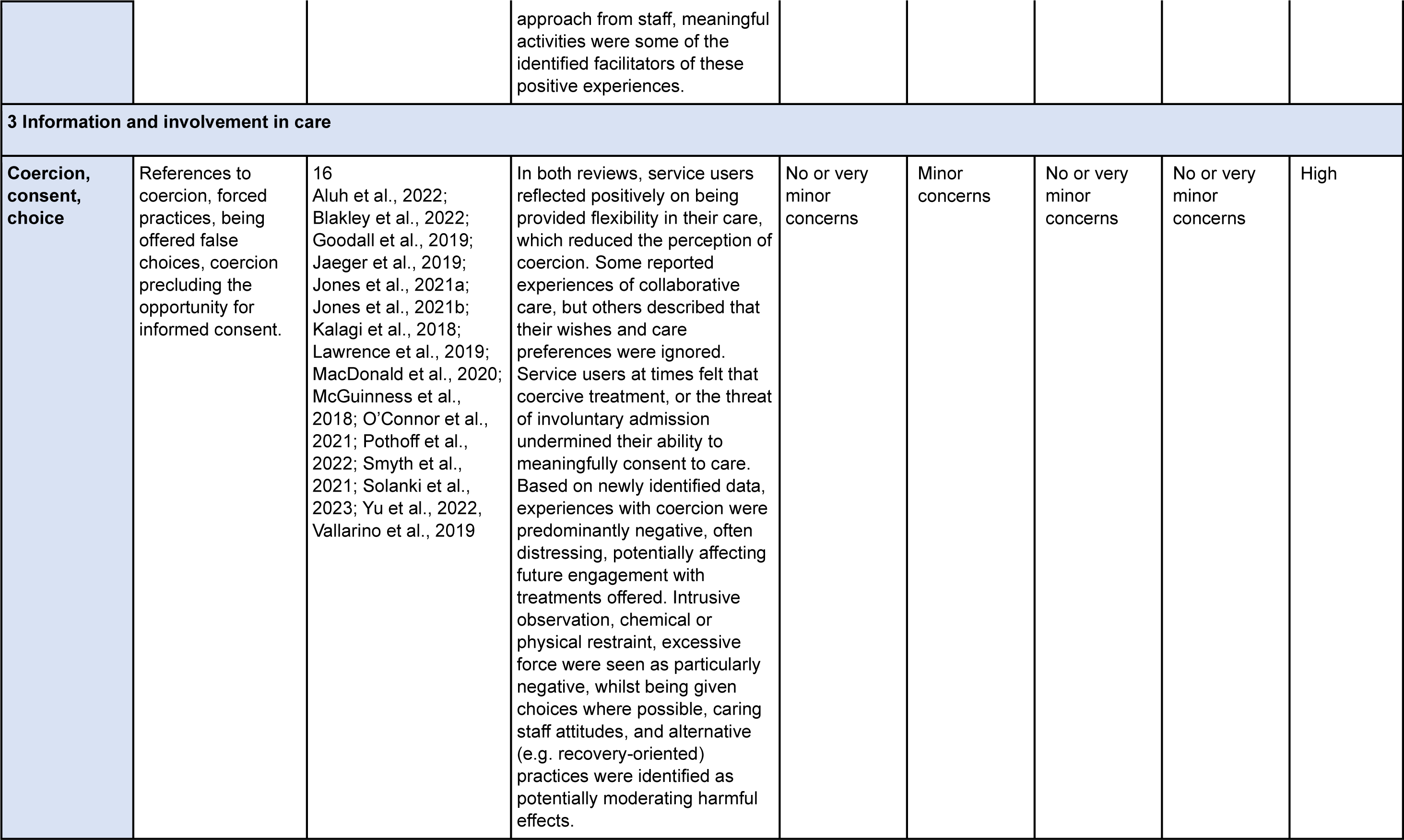
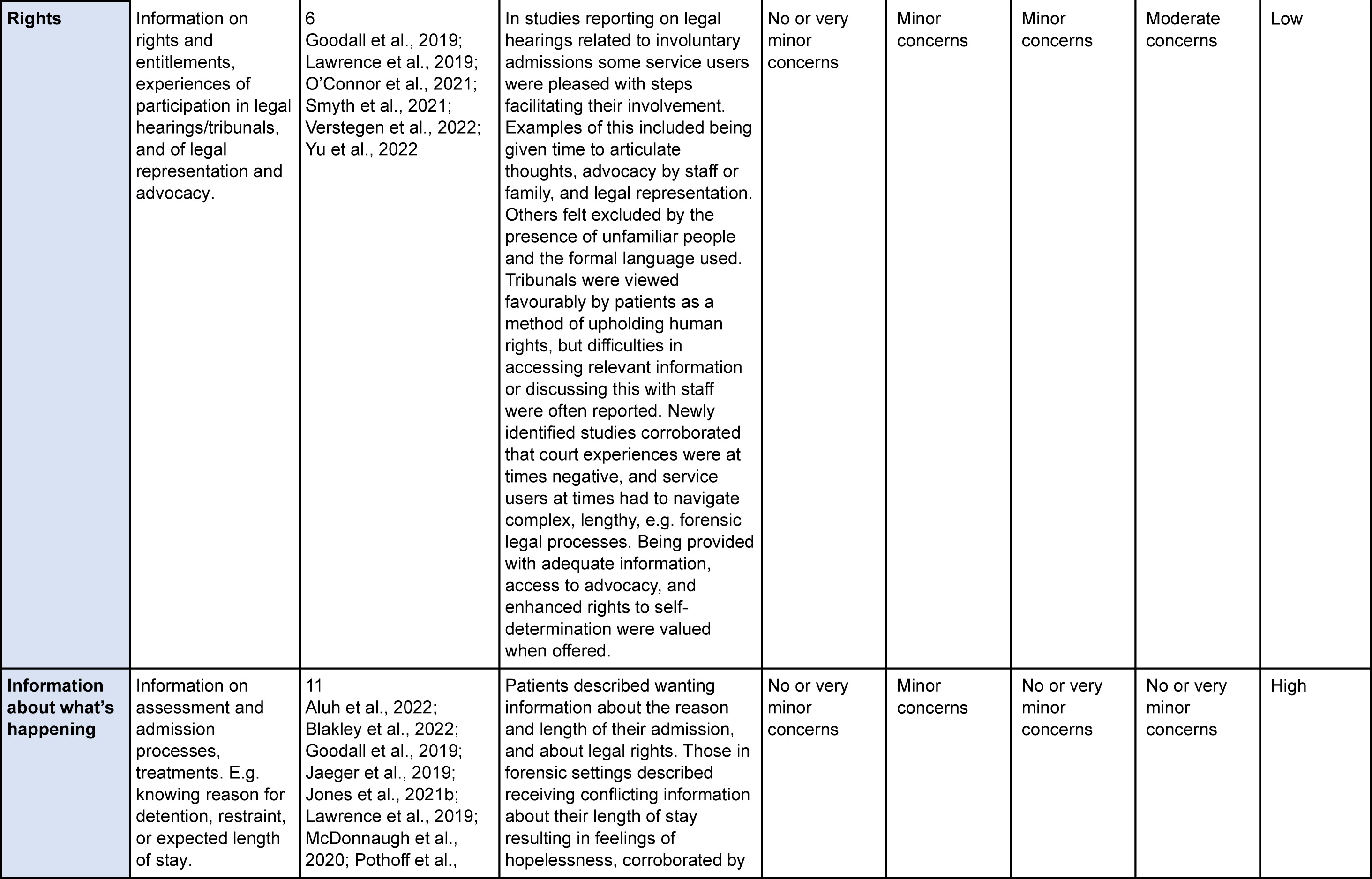
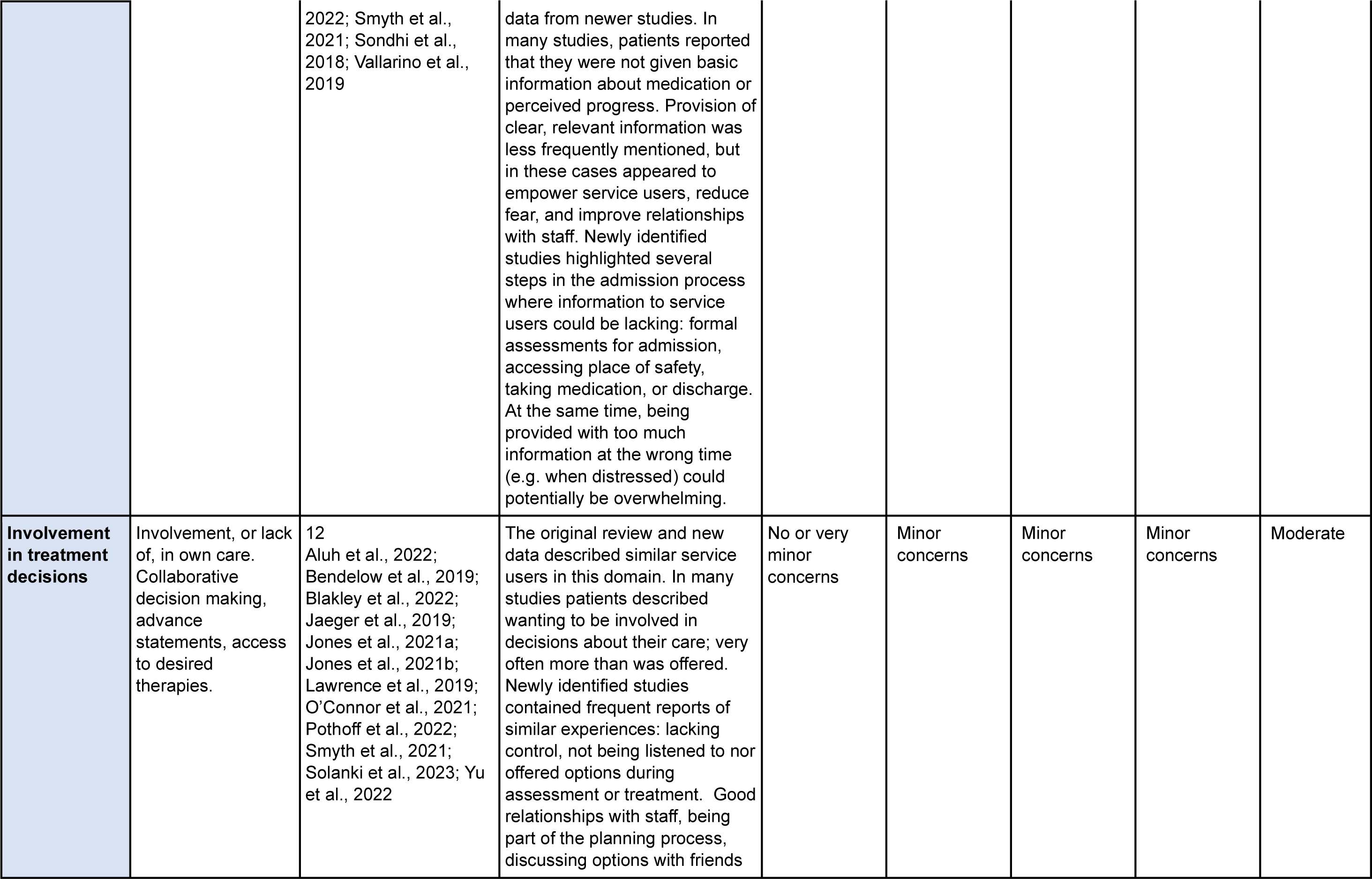
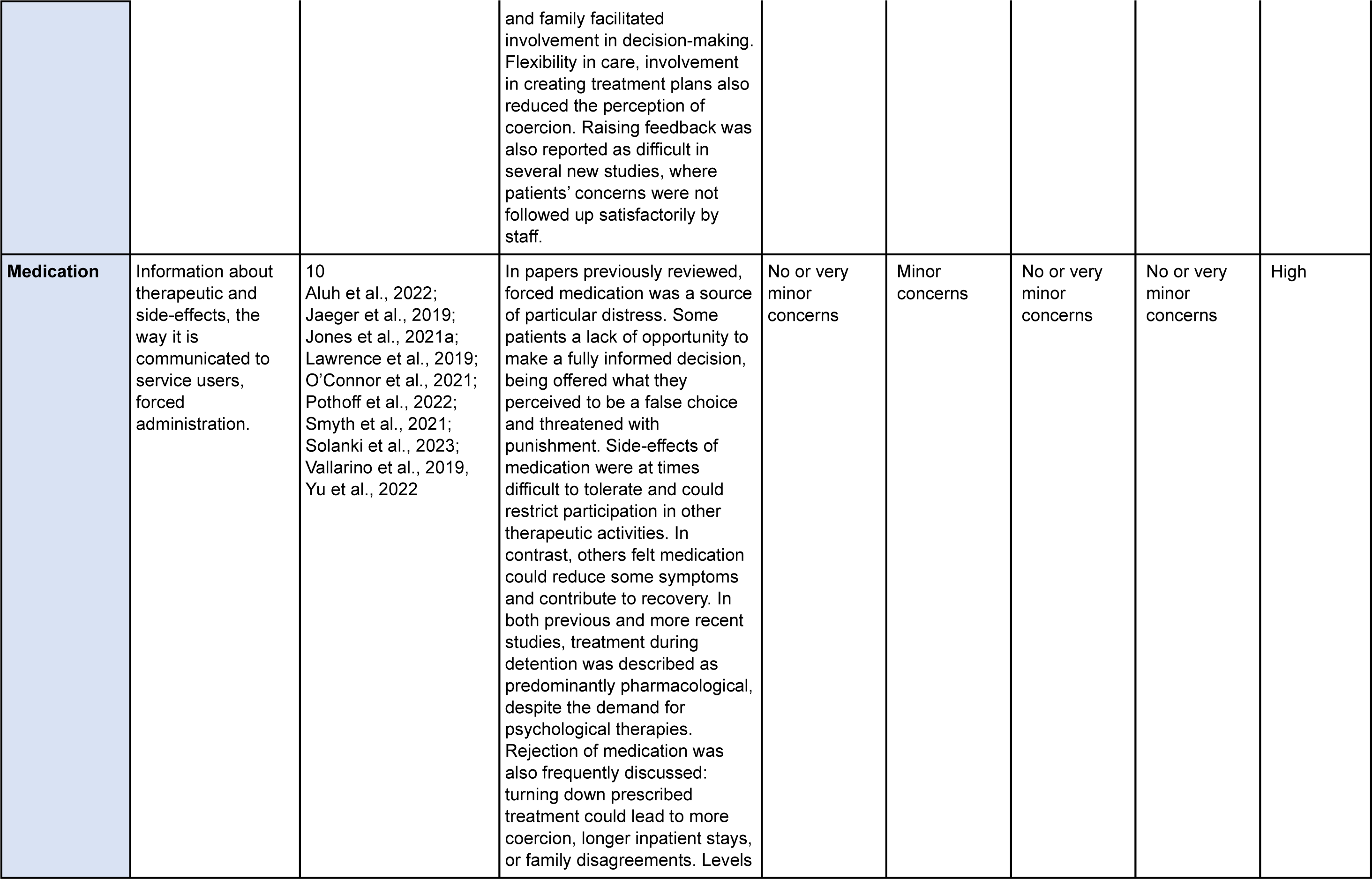
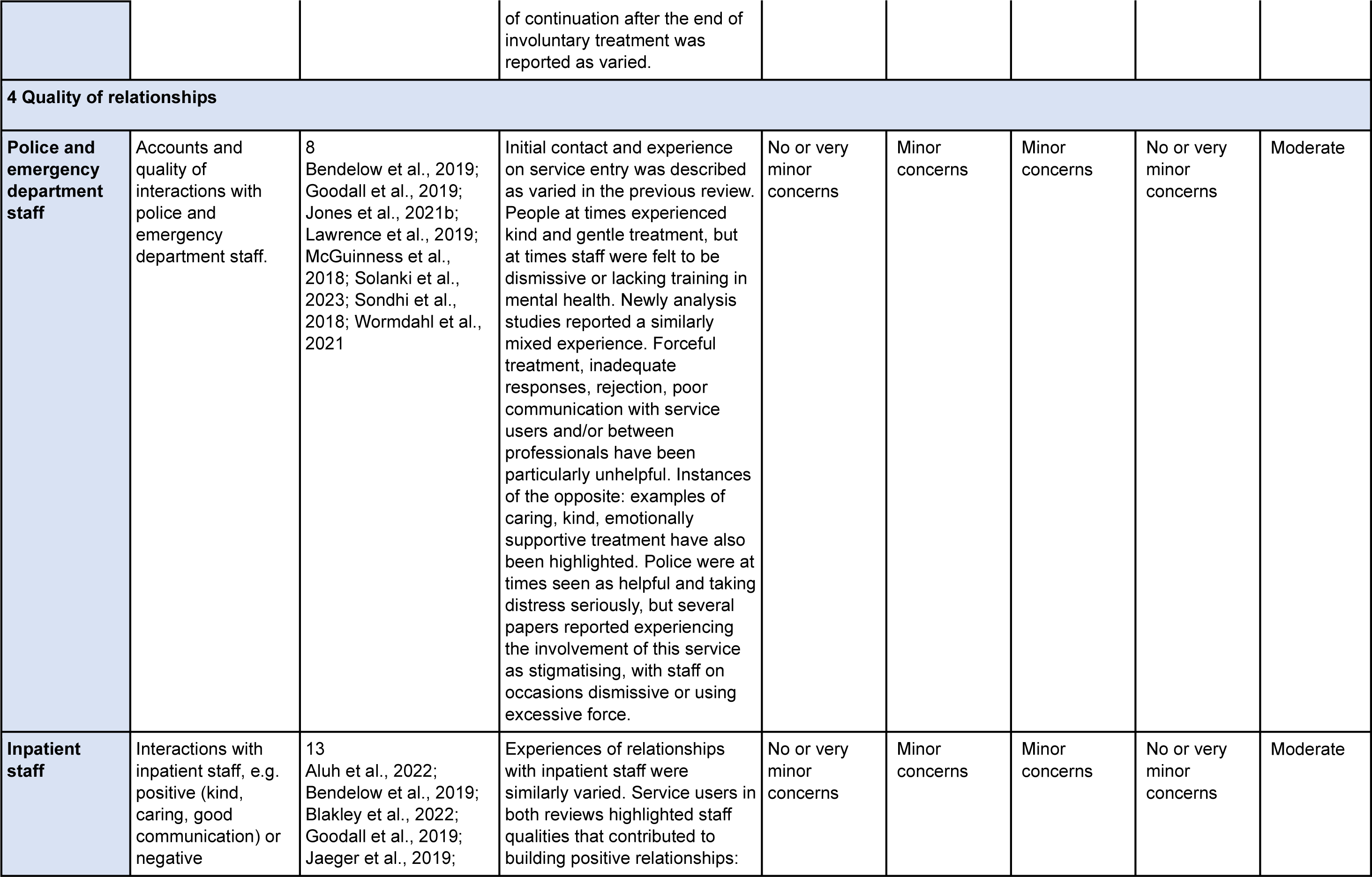
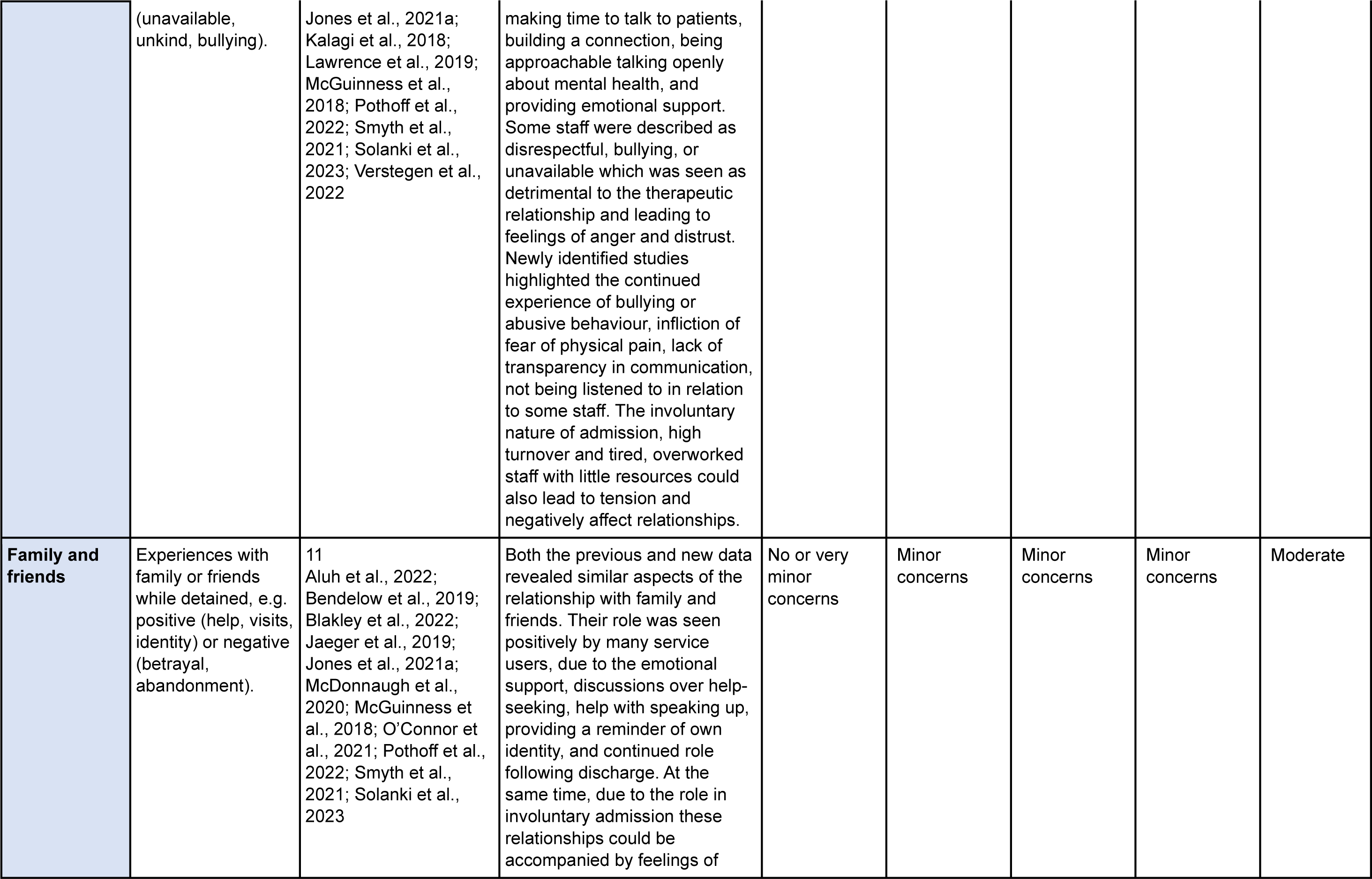
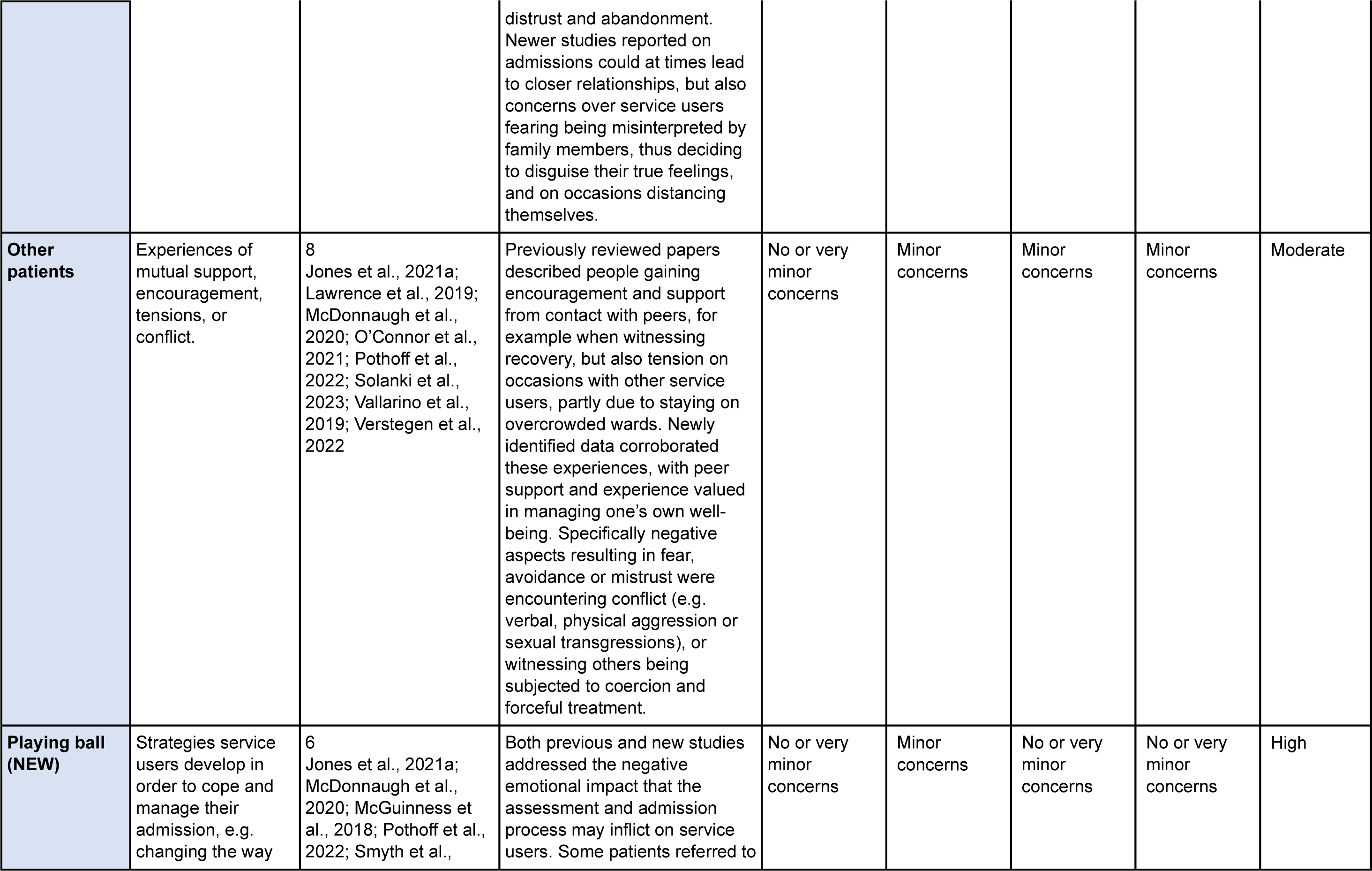
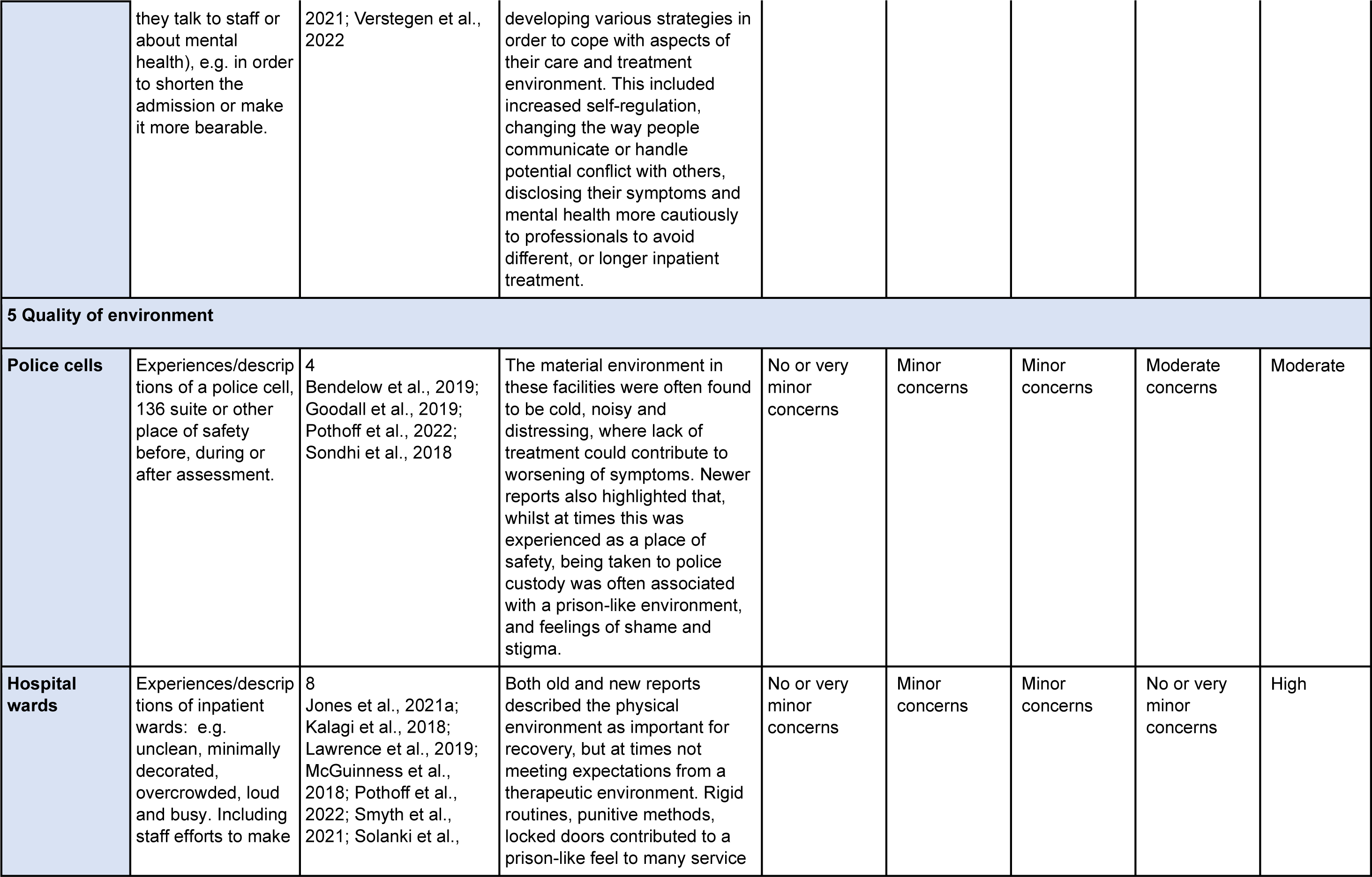
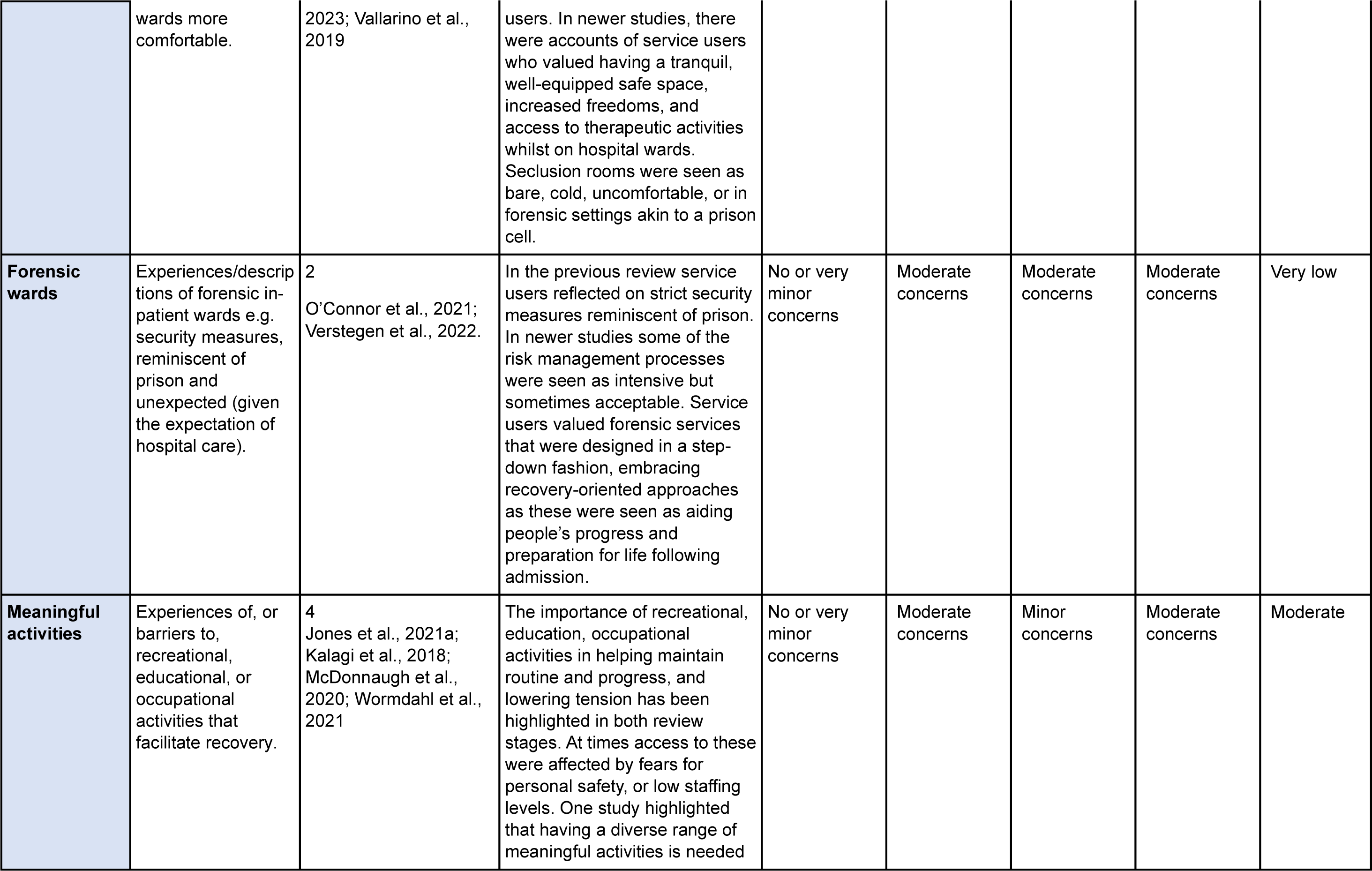
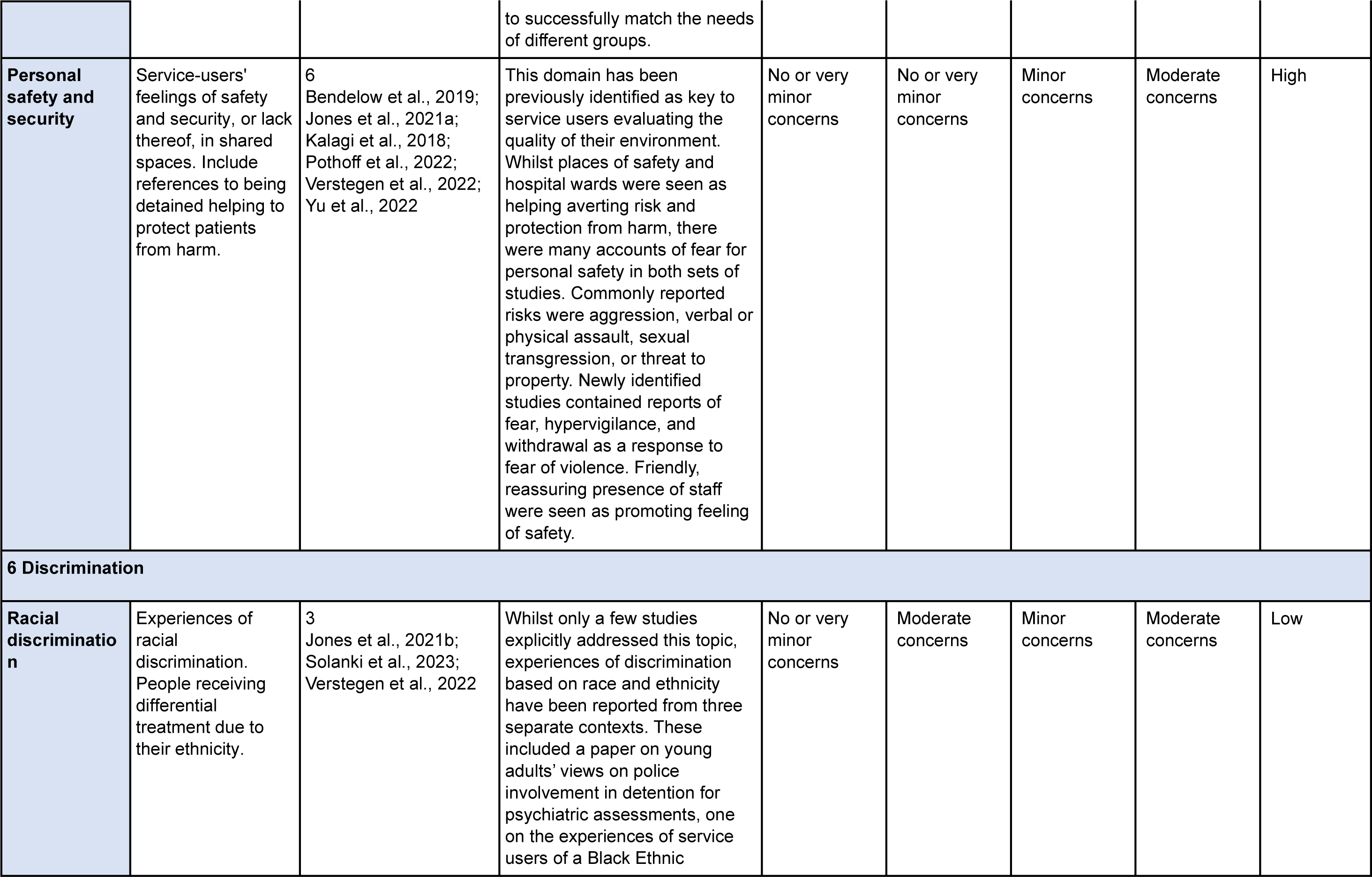
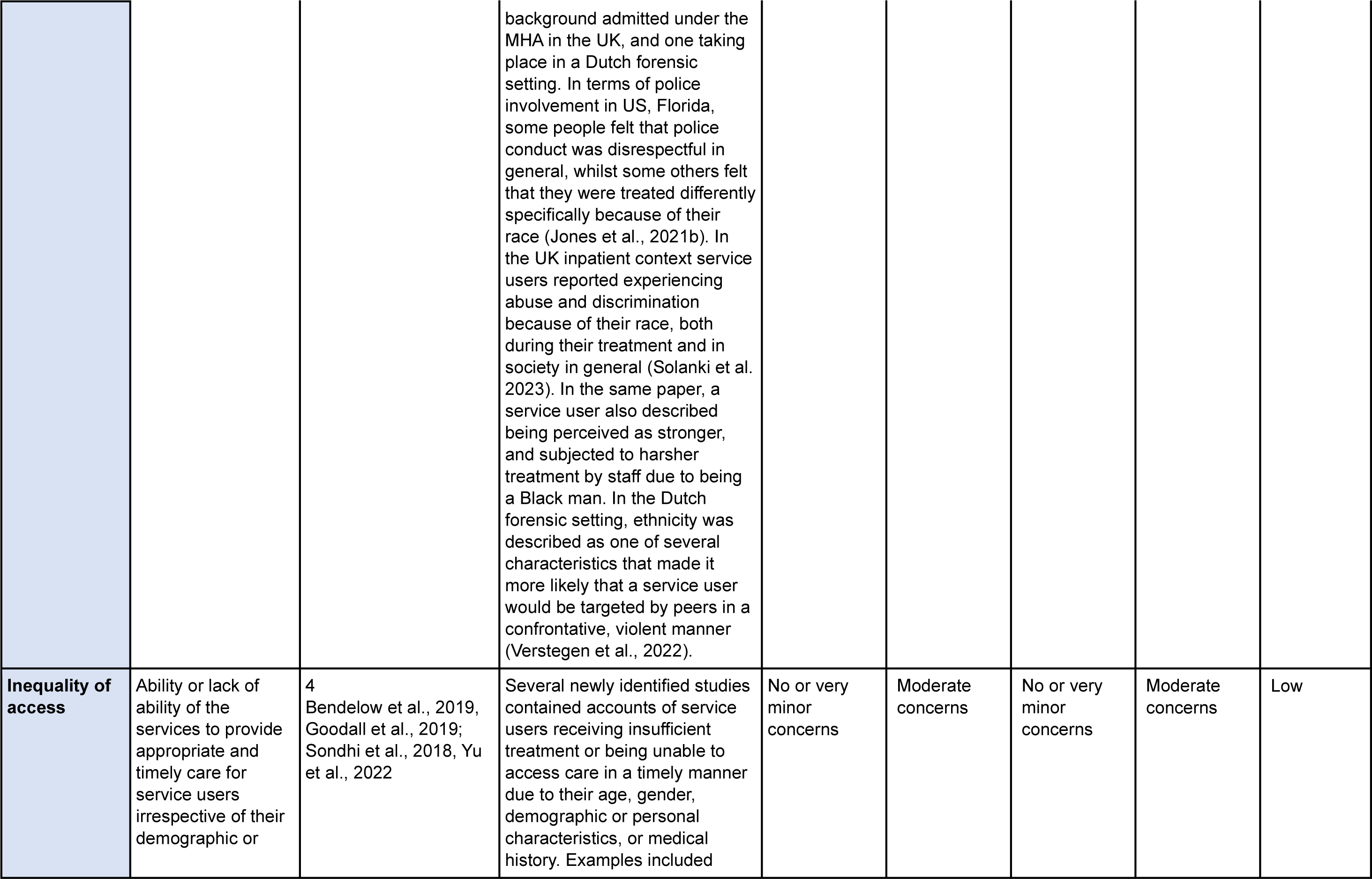
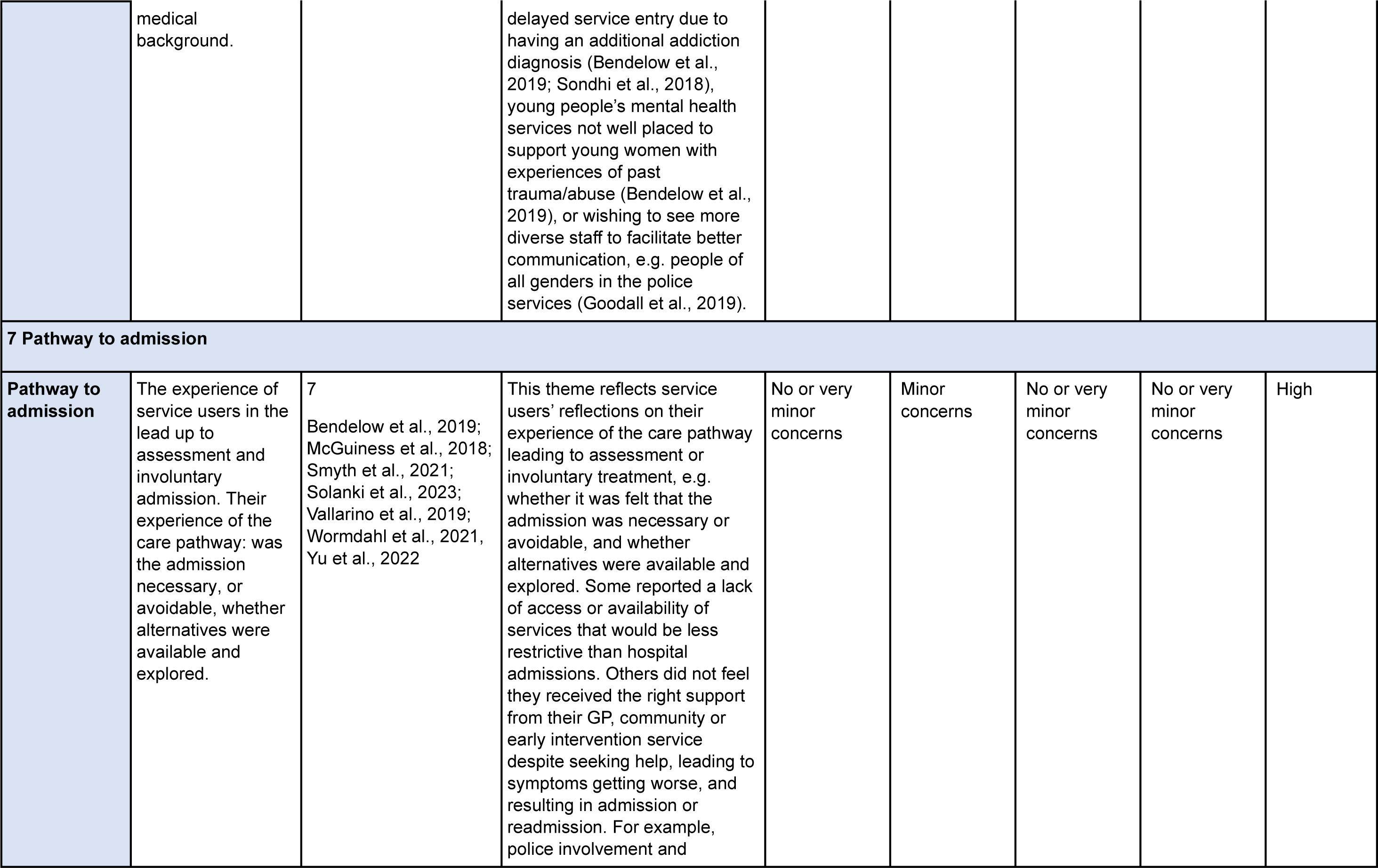
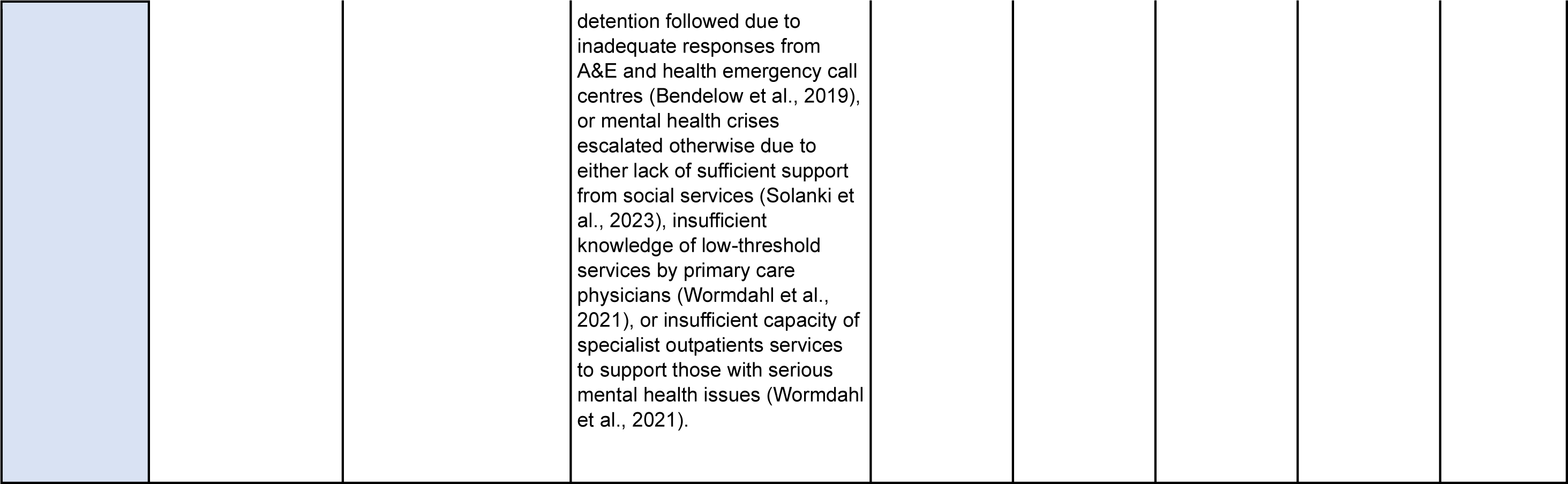
Summary of findings: Thematic framework, service user themes and sub-themes, certainty of evidence assessment.

| Themes and sub-themes | Brief description | Number of papers contributing | Finding | CERQual Quality Assessment |  |  |  |  |
| --- | --- | --- | --- | --- | --- | --- | --- | --- |
|  |  |  |  | Methodological limitations | Relevance of evidence | Coherence of finding | Adequacy of data | Overall confidence |
| 1 Emotional impact |  |  |  |  |  |  |  |  |
| Acceptance | Appreciation or acceptance of the need for admission and treatment. | 8<br>Bendelow et al., 2019; Lawrence et al., 2019; MacDonald et al., 2020; O'Connor et al., 2021; Smyth et al., 2021; Vallarino et al., 2019, Yu et al., 2022; Aluh et al., 2022 | The previous review highlighted occasions when participants appreciated or accepted the involuntary admission as necessary in some cases. Additional data from more recent papers similarly referred to positive aspects like feeling safe, addressing an underlying need and accessing support. Acceptance was nevertheless motivated on occasions by wanting to avoid conflict and the threat of coercion. | No or very minor concerns | Minor concerns | Minor concerns | Minor concerns | Moderate |
| Impact of detention | Emotional impact, e.g. distress, fear, confusion due to being detained. | 18<br>Aluh et al., 2022; Bendelow et al., 2019; Blakley et al., 2022; Goodall et al., 2019; Jaeger et al., 2019; Jones et al., 2021a; Jones et al., 2021b; Lawrence et al., 2019; McDonnaugh et al., 2020; McGuinness et al., 2018; O'Connor et al., 2021; Pothoff et al., | The previous synthesis identified anger, confusion, fear, distress, resentment and defensiveness as common emotional impacts detention, exacerbated by lack of information, police involvement, and behaviour of staff. The update uncovered similarly frequent, traumatic experiences, undergoing complex triage processes, away from one's usual environment and dealing with unfamiliar people. Arriving | No or very minor concerns | No or very minor concerns | Minor concerns | No or very minor concerns | High |
|  |  | 2022; Smyth et al., 2021; Sondhi et al., 2018; Vallarino et al., 2019, Verstegen et al., 2022; Wormdahl et al., 2021; Yu et al., 2022 | to a place of safety was seen as the beginning of receiving support, but often a distressing, chaotic, high-risk environment. Detention in hospital settings was often described as negative, seen at times as punishment, barbaric, or a source of violent confrontations. Positive experiences with staff could at times ameliorate emotional impact, for example encountering a caring attitude, being listened to, or being trusted and offered control. |  |  |  |  |  |
| **Impact of coercive treatment** | Emotional impact due to experiencing or witnessing coercion, e.g. restraint, excessive force etc. | 8<br>Aluh et al., 2022; Goodall et al., 2019; Kalagi et al., 2018; MacDonald et al., 2020; McGuinness et al., 2018; Pothoff et al., 2022; Solanki et al., 2023; Yu et al., 2022 | In both the original and updated data coercive treatments were often described as an abusive, violating experience contributing to a sense of lack of control and a strong negative emotional impact (e.g. anxiety, fear, distress and dehumanisation). For example, seclusion triggered feelings of anger, loneliness and shame. Coercive practices in newly identified data included physical (including mechanical) and chemical, constant observation, or a locked environment. The use of mechanical restraint (referred to in a couple of studies) was linked with the experience of pain, humiliation and perception of assault. Witnessing other service users being restrained could be perceived as similarly fear provoking. Coercive practices were perceived unnecessary by some, justified only in limited instances (e.g. serious risk to life). serious risks to life. Some studies also referred to a vicious cycle in which coercion leads to increased anger and aggression, which in turn may be responded to with more coercion (e.g. restraint) by staff. | No or very minor concerns | Minor concerns | No or very minor concerns | Minor concerns | Moderate |
| **Feelings following discharge** | Feelings following discharge, e.g. resentment, worry, or positive impacts like feeling motivated, supported. | 8<br>Jones et al., 2021a; MacDonald et al., 2020; McDonnaugh et al., 2020; McGuinness et al., 2018; O'Connor et al., 2021; Smyth et al., 2021; Sondhi et al., 2018; Yu et al., 2022 | In the original review service users reported often feeling worse following admission than before, corroborated by new data. The negative impact of detention could be long lasting and lead to an increase in symptoms (e.g. depression, stress). Fear of readmission could affect relationships with community mental health services as well as family members. Some reported having positive experiences following discharge, for example having an increased motivation for change, focusing more on self-care, and seeing part of treatment as helpful to recovery. Returning to community life was seen as difficult at times, for example coping with less support, lacking adequate information on or access to services post-discharge. Some facilitators of successful discharge were emotional and practical support, engagement with community and social support, and a staged step-down process. | No or very minor concerns | Minor concerns | Minor concerns | Minor concerns | Moderate |
| **Therapeutic benefit (NEW)** | Whether the assessment or involuntary treatment was seen as therapeutic, helping recovery or not. | 11<br>Aluh et al., 2022; Jaeger et al., 2019; Jones et al., 2021a; Kalagi et al., 2018; MacDonald et al., 2023; McGuinness et al., 2018; O'Connor et al., 2021; Smyth et al., 2021; Solanki et al., 2023; Vallarino et al., 2019; Yu et al., 2022 | In addition to the impact on emotions and self-worth, in recently identified studies service users' expressed views on whether they saw the assessment and involuntary treatment as necessary and therapeutic, and whether it helped their recovery. Service users' experiences were mixed in this domain. In some cases, admissions were described as providing little meaningful help in addressing distress or psychological help and in some cases lead to feeling worse psychologically. The nature of experiences and interactions with staff during admission (e.g. coercive, or inclusive) could affect trust and engagement with therapy in later stages. At the same time, in some instances therapeutic value was reported, for example aiding recovery, contributing to an increased care for oneself, or changing perspectives on managing life after admission. | No or very minor concerns | Minor concerns | Moderate concerns | No or very minor concerns | Moderate |
| **2 Impact on self-worth** |  |  |  |  |  |  |  |  |
| **Dehumanised** | Not treated humanely by staff, having human rights violated, being | 12<br>Aluh et al., 2022; Goodall et al., 2019; Jaeger et al., 2019; | Previously, service users reported feeling dehumanised by coercive interventions, supported by newly identified | No or very minor concerns | Minor concerns | No or very minor concerns | No or very minor concerns | High |
|  | reduced to a diagnosis, normal emotions regarded as symptoms. | Jones et al., 2021a; MacDonald et al., 2020; Pothoff et al., 2022; Smyth et al., 2021; Wormdahl et al (2021); Yu et al (2022); Sondhi et al (2018); Verstegen et al (2022); Solanki (2023) | data. Some described feeling like a caged animal during involuntary treatment, a sense of loss of identity, with everyday behaviours or traits being interpreted as a sign of illness or symptom by others. Staff showing genuine concern, treating people with dignity could have a positive effect on self-esteem. |  |  |  |  |  |
| **Power** | Having or lacking autonomy or control over treatments and how to spend own time. | 12<br>Aluh et al., 2022; Blakley et al., 2022; Goodall et al., 2019; Jones et al., 2021a; Jones et al., 2021b; Lawrence et al., 2019; McDonnaugh et al., 2020; McGuinness et al., 2018; O'Connor et al., 2021; Pothoff et al., 2022; Vallarino et al., 2019; Verstegen et al., 2022 | Service users across a number of previously reviewed papers reported having a lack of control over the treatment process, e.g. having limited choices, lacking autonomy, not having an impact on timelines, or arbitrary ward routines. There were multiple references to power differences, paternalistic attitude by staff, needing permission, leading to feelings of dependency and reduced self-efficacy. Not being listened to, coercion (e.g. locked doors) could have a negative effect, whilst instances of collaborative care, being provided with choices, advocacy by others could have a positive effect on perceived control and autonomy during admission. | No or very minor concerns | Minor concerns | Minor concerns | No or very minor concerns | Moderate |
| **Stigma** | Feeling labelled or criminalised, losing credibility, being treated as dangerous or | 14<br>Aluh et al., 2022; Bendelow et al., 2019; Goodall et al., 2019; Jaeger et al., 2019; | There were frequent reports of service users feeling labelled, tainted or criminalised as a consequence of detention. For example, experiencing shame | No or very minor concerns | Minor concerns | No or very minor concerns | No or very minor concerns | High |
|  | marginalised for having a mental illness. | Jones et al., 2021a; Jones et al., 2021b; McDonnaugh et al., 2020; McGuinness et al., 2018; Smyth et al., 2021; Solanki et al., 2023; Sondhi et al., 2018; Vallarino et al., 2019; Wormdahl et al., 2021; Yu et al., 2022 | when neighbours witness them being handcuffed and taken by police, or losing credibility and being treated as dangerous for having a mental illness both within the hospital but also by society. There were reports of fear of being excluded and marginalised post-discharge and feeling at a greater risk of being detained again. Newer data corroborated experiences of violation and loss of rights, with frequent experiences of criminalisation, and not being believed. A few factors were seen as having a positive effect on stigma, for promoting open discussion on mental health, self-disclosure by professionals and public figures, and changing public and legal perceptions of mental health. |  |  |  |  |  |
| **Positive impacts** | Experiences that built confidence, self-esteem, self-respect, self-worth, or a sense of achievement. | 8<br>Jones et al., 2021a; Jones et al., 2021b; Kalagi et al., 2018; MacDonald et al., 2020; O'Connor et al., 2021; Pothoff et al., 2022; Smyth et al., 2021; Solanki et al., 2023 | Service users also referenced activities or experiences during detention that built confidence, self-esteem, self-respect, or a sense of achievement. In newly identified data, admission was at times described as a turning point, contributing to becoming more independent, motivation to return to everyday life, or receiving more recognition of mental health from family members. Social proximity of others, working with skilled, recovery-oriented professionals, compassionate, genuine approach from staff, meaningful activities were some of the identified facilitators of these positive experiences. | No or very minor concerns | Minor concerns | Minor concerns | Minor concerns | Moderate |
| **3 Information and involvement in care** |  |  |  |  |  |  |  |  |
| **Coercion, consent, choice** | References to coercion, forced practices, being offered false choices, coercion precluding the opportunity for informed consent. | 16<br>Aluh et al., 2022; Blakley et al., 2022; Goodall et al., 2019; Jaeger et al., 2019; Jones et al., 2021a; Jones et al., 2021b; Kalagi et al., 2018; Lawrence et al., 2019; MacDonald et al., 2020; McGuinness et al., 2018; O'Connor et al., 2021; Pothoff et al., 2022; Smyth et al., 2021; Solanki et al., 2023; Yu et al., 2022, Vallarino et al., 2019 | In both reviews, service users reflected positively on being provided flexibility in their care, which reduced the perception of coercion. Some reported experiences of collaborative care, but others described that their wishes and care preferences were ignored. Service users at times felt that coercive treatment, or the threat of involuntary admission undermined their ability to meaningfully consent to care. Based on newly identified data, experiences with coercion were predominantly negative, often distressing, potentially affecting future engagement with treatments offered. Intrusive observation, chemical or physical restraint, excessive force were seen as particularly negative, whilst being given choices where possible, caring staff attitudes, and alternative (e.g. recovery-oriented) practices were identified as potentially moderating harmful effects. | No or very minor concerns | Minor concerns | No or very minor concerns | No or very minor concerns | High |
| **Rights** | Information on rights and entitlements, experiences of participation in legal hearings/tribunals, and of legal representation and advocacy. | 6<br>Goodall et al., 2019;<br>Lawrence et al., 2019;<br>O'Connor et al., 2021;<br>Smyth et al., 2021;<br>Verstegen et al., 2022;<br>Yu et al., 2022 | In studies reporting on legal hearings related to involuntary admissions some service users were pleased with steps facilitating their involvement. Examples of this included being given time to articulate thoughts, advocacy by staff or family, and legal representation. Others felt excluded by the presence of unfamiliar people and the formal language used. Tribunals were viewed favourably by patients as a method of upholding human rights, but difficulties in accessing relevant information or discussing this with staff were often reported. Newly identified studies corroborated that court experiences were at times negative, and service users at times had to navigate complex, lengthy, e.g. forensic legal processes. Being provided with adequate information, access to advocacy, and enhanced rights to self-determination were valued when offered. | No or very minor concerns | Minor concerns | Minor concerns | Moderate concerns | Low |
| **Information about what's happening** | Information on assessment and admission processes, treatments. E.g. knowing reason for detention, restraint, or expected length of stay. | 11<br>Aluh et al., 2022;<br>Blakley et al., 2022;<br>Goodall et al., 2019;<br>Jaeger et al., 2019;<br>Jones et al., 2021b;<br>Lawrence et al., 2019;<br>McDonnaugh et al., 2020;<br>Pothoff et al., | Patients described wanting information about the reason and length of their admission, and about legal rights. Those in forensic settings described receiving conflicting information about their length of stay resulting in feelings of hopelessness, corroborated by | No or very minor concerns | Minor concerns | No or very minor concerns | No or very minor concerns | High |
|  |  | 2022; Smyth et al., 2021; Sondhi et al., 2018; Vallarino et al., 2019 | data from newer studies. In many studies, patients reported that they were not given basic information about medication or perceived progress. Provision of clear, relevant information was less frequently mentioned, but in these cases appeared to empower service users, reduce fear, and improve relationships with staff. Newly identified studies highlighted several steps in the admission process where information to service users could be lacking: formal assessments for admission, accessing place of safety, taking medication, or discharge. At the same time, being provided with too much information at the wrong time (e.g. when distressed) could potentially be overwhelming. |  |  |  |  |  |
| **Involvement in treatment decisions** | Involvement, or lack of, in own care. Collaborative decision making, advance statements, access to desired therapies. | 12<br>Aluh et al., 2022; Bendelow et al., 2019; Blakley et al., 2022; Jaeger et al., 2019; Jones et al., 2021a; Jones et al., 2021b; Lawrence et al., 2019; O'Connor et al., 2021; Pothoff et al., 2022; Smyth et al., 2021; Solanki et al., 2023; Yu et al., 2022 | The original review and new data described similar service users in this domain. In many studies patients described wanting to be involved in decisions about their care; very often more than was offered. Newly identified studies contained frequent reports of similar experiences: lacking control, not being listened to nor offered options during assessment or treatment. Good relationships with staff, being part of the planning process, discussing options with friends and family facilitated involvement in decision-making. Flexibility in care, involvement in creating treatment plans also reduced the perception of coercion. Raising feedback was also reported as difficult in several new studies, where patients' concerns were not followed up satisfactorily by staff. | No or very minor concerns | Minor concerns | Minor concerns | Minor concerns | Moderate |
| **Medication** | Information about therapeutic and side-effects, the way it is communicated to service users, forced administration. | 10<br>Aluh et al., 2022;<br>Jaeger et al., 2019;<br>Jones et al., 2021a;<br>Lawrence et al., 2019;<br>O'Connor et al., 2021;<br>Pothoff et al., 2022;<br>Smyth et al., 2021;<br>Solanki et al., 2023;<br>Vallarino et al., 2019,<br>Yu et al., 2022 | In papers previously reviewed, forced medication was a source of particular distress. Some patients a lack of opportunity to make a fully informed decision, being offered what they perceived to be a false choice and threatened with punishment. Side-effects of medication were at times difficult to tolerate and could restrict participation in other therapeutic activities. In contrast, others felt medication could reduce some symptoms and contribute to recovery. In both previous and more recent studies, treatment during detention was described as predominantly pharmacological, despite the demand for psychological therapies. Rejection of medication was also frequently discussed: turning down prescribed treatment could lead to more coercion, longer inpatient stays, or family disagreements. Levels of continuation after the end of involuntary treatment was reported as varied. | No or very minor concerns | Minor concerns | No or very minor concerns | No or very minor concerns | High |
| **4 Quality of relationships** |  |  |  |  |  |  |  |  |
| **Police and emergency department staff** | Accounts and quality of interactions with police and emergency department staff. | 8<br>Bendelow et al., 2019; Goodall et al., 2019; Jones et al., 2021b; Lawrence et al., 2019; McGuinness et al., 2018; Solanki et al., 2023; Sondhi et al., 2018; Wormdahl et al., 2021 | Initial contact and experience on service entry was described as varied in the previous review. People at times experienced kind and gentle treatment, but at times staff were felt to be dismissive or lacking training in mental health. Newly analysis studies reported a similarly mixed experience. Forceful treatment, inadequate responses, rejection, poor communication with service users and/or between professionals have been particularly unhelpful. Instances of the opposite: examples of caring, kind, emotionally supportive treatment have also been highlighted. Police were at times seen as helpful and taking distress seriously, but several papers reported experiencing the involvement of this service as stigmatising, with staff on occasions dismissive or using excessive force. | No or very minor concerns | Minor concerns | Minor concerns | No or very minor concerns | Moderate |
| **Inpatient staff** | Interactions with inpatient staff, e.g. positive (kind, caring, good communication) or negative | 13<br>Aluh et al., 2022; Bendelow et al., 2019; Blakley et al., 2022; Goodall et al., 2019; Jaeger et al., 2019; | Experiences of relationships with inpatient staff were similarly varied. Service users in both reviews highlighted staff qualities that contributed to building positive relationships: | No or very minor concerns | Minor concerns | Minor concerns | No or very minor concerns | Moderate |
|  | (unavailable, unkind, bullying). | Jones et al., 2021a; Kalagi et al., 2018; Lawrence et al., 2019; McGuinness et al., 2018; Pothoff et al., 2022; Smyth et al., 2021; Solanki et al., 2023; Verstegen et al., 2022 | making time to talk to patients, building a connection, being approachable talking openly about mental health, and providing emotional support. Some staff were described as disrespectful, bullying, or unavailable which was seen as detrimental to the therapeutic relationship and leading to feelings of anger and distrust. Newly identified studies highlighted the continued experience of bullying or abusive behaviour, infliction of fear of physical pain, lack of transparency in communication, not being listened to in relation to some staff. The involuntary nature of admission, high turnover and tired, overworked staff with little resources could also lead to tension and negatively affect relationships. |  |  |  |  |  |
| **Family and friends** | Experiences with family or friends while detained, e.g. positive (help, visits, identity) or negative (betrayal, abandonment). | 11<br>Aluh et al., 2022; Bendelow et al., 2019; Blakley et al., 2022; Jaeger et al., 2019; Jones et al., 2021a; McDonnaugh et al., 2020; McGuinness et al., 2018; O'Connor et al., 2021; Pothoff et al., 2022; Smyth et al., 2021; Solanki et al., 2023 | Both the previous and new data revealed similar aspects of the relationship with family and friends. Their role was seen positively by many service users, due to the emotional support, discussions over help-seeking, help with speaking up, providing a reminder of own identity, and continued role following discharge. At the same time, due to the role in involuntary admission these relationships could be accompanied by feelings of distrust and abandonment. Newer studies reported on admissions could at times lead to closer relationships, but also concerns over service users fearing being misinterpreted by family members, thus deciding to disguise their true feelings, and on occasions distancing themselves. | No or very minor concerns | Minor concerns | Minor concerns | Minor concerns | Moderate |
| **Other patients** | Experiences of mutual support, encouragement, tensions, or conflict. | 8<br>Jones et al., 2021a; Lawrence et al., 2019; McDonnaugh et al., 2020; O'Connor et al., 2021; Pothoff et al., 2022; Solanki et al., 2023; Vallarino et al., 2019; Versteegen et al., 2022 | Previously reviewed papers described people gaining encouragement and support from contact with peers, for example when witnessing recovery, but also tension on occasions with other service users, partly due to staying on overcrowded wards. Newly identified data corroborated these experiences, with peer support and experience valued in managing one's own well-being. Specifically negative aspects resulting in fear, avoidance or mistrust were encountering conflict (e.g. verbal, physical aggression or sexual transgressions), or witnessing others being subjected to coercion and forceful treatment. | No or very minor concerns | Minor concerns | Minor concerns | Minor concerns | Moderate |
| **Playing ball (NEW)** | Strategies service users develop in order to cope and manage their admission, e.g. changing the way | 6<br>Jones et al., 2021a; McDonnaugh et al., 2020; McGuinness et al., 2018; Pothoff et al., 2022; Smyth et al., | Both previous and new studies addressed the negative emotional impact that the assessment and admission process may inflict on service users. Some patients referred to | No or very minor concerns | Minor concerns | No or very minor concerns | No or very minor concerns | High |
|  | they talk to staff or about mental health), e.g. in order to shorten the admission or make it more bearable. | 2021; Verstegen et al., 2022 | developing various strategies in order to cope with aspects of their care and treatment environment. This included increased self-regulation, changing the way people communicate or handle potential conflict with others, disclosing their symptoms and mental health more cautiously to professionals to avoid different, or longer inpatient treatment. |  |  |  |  |  |
| **5 Quality of environment** |  |  |  |  |  |  |  |  |
| **Police cells** | Experiences/descriptions of a police cell, 136 suite or other place of safety before, during or after assessment. | 4<br>Bendelow et al., 2019; Goodall et al., 2019; Pothoff et al., 2022; Sondhi et al., 2018 | The material environment in these facilities were often found to be cold, noisy and distressing, where lack of treatment could contribute to worsening of symptoms. Newer reports also highlighted that, whilst at times this was experienced as a place of safety, being taken to police custody was often associated with a prison-like environment, and feelings of shame and stigma. | No or very minor concerns | Minor concerns | Minor concerns | Moderate concerns | Moderate |
| **Hospital wards** | Experiences/descriptions of inpatient wards: e.g. unclean, minimally decorated, overcrowded, loud and busy. Including staff efforts to make | 8<br>Jones et al., 2021a; Kalagi et al., 2018; Lawrence et al., 2019; McGuinness et al., 2018; Pothoff et al., 2022; Smyth et al., 2021; Solanki et al., | Both old and new reports described the physical environment as important for recovery, but at times not meeting expectations from a therapeutic environment. Rigid routines, punitive methods, locked doors contributed to a prison-like feel to many service | No or very minor concerns | Minor concerns | Minor concerns | No or very minor concerns | High |
|  | wards more comfortable. | 2023; Vallarino et al., 2019 | users. In newer studies, there were accounts of service users who valued having a tranquil, well-equipped safe space, increased freedoms, and access to therapeutic activities whilst on hospital wards. Seclusion rooms were seen as bare, cold, uncomfortable, or in forensic settings akin to a prison cell. |  |  |  |  |  |
| **Forensic wards** | Experiences/descriptions of forensic in-patient wards e.g. security measures, reminiscent of prison and unexpected (given the expectation of hospital care). | 2<br>O'Connor et al., 2021; Verstegen et al., 2022. | In the previous review service users reflected on strict security measures reminiscent of prison. In newer studies some of the risk management processes were seen as intensive but sometimes acceptable. Service users valued forensic services that were designed in a step-down fashion, embracing recovery-oriented approaches as these were seen as aiding people's progress and preparation for life following admission. | No or very minor concerns | Moderate concerns | Moderate concerns | Moderate concerns | Very low |
| **Meaningful activities** | Experiences of, or barriers to, recreational, educational, or occupational activities that facilitate recovery. | 4<br>Jones et al., 2021a; Kalagi et al., 2018; McDonnaugh et al., 2020; Wormdahl et al., 2021 | The importance of recreational, education, occupational activities in helping maintain routine and progress, and lowering tension has been highlighted in both review stages. At times access to these were affected by fears for personal safety, or low staffing levels. One study highlighted that having a diverse range of meaningful activities is needed to successfully match the needs of different groups. | No or very minor concerns | Moderate concerns | Minor concerns | Moderate concerns | Moderate |
| **Personal safety and security** | Service-users' feelings of safety and security, or lack thereof, in shared spaces. Include references to being detained helping to protect patients from harm. | 6<br>Bendelow et al., 2019;<br>Jones et al., 2021a;<br>Kalagi et al., 2018;<br>Pothoff et al., 2022;<br>Verstegen et al., 2022;<br>Yu et al., 2022 | This domain has been previously identified as key to service users evaluating the quality of their environment. Whilst places of safety and hospital wards were seen as helping averting risk and protection from harm, there were many accounts of fear for personal safety in both sets of studies. Commonly reported risks were aggression, verbal or physical assault, sexual transgression, or threat to property. Newly identified studies contained reports of fear, hypervigilance, and withdrawal as a response to fear of violence. Friendly, reassuring presence of staff were seen as promoting feeling of safety. | No or very minor concerns | No or very minor concerns | Minor concerns | Moderate concerns | High |
| **6 Discrimination** |  |  |  |  |  |  |  |  |
| **Racial discrimination** | Experiences of racial discrimination. People receiving differential treatment due to their ethnicity. | 3<br>Jones et al., 2021b;<br>Solanki et al., 2023;<br>Verstegen et al., 2022 | Whilst only a few studies explicitly addressed this topic, experiences of discrimination based on race and ethnicity have been reported from three separate contexts. These included a paper on young adults' views on police involvement in detention for psychiatric assessments, one on the experiences of service users of a Black Ethnic background admitted under the MHA in the UK, and one taking place in a Dutch forensic setting. In terms of police involvement in US, Florida, some people felt that police conduct was disrespectful in general, whilst some others felt that they were treated differently specifically because of their race (Jones et al., 2021b). In the UK inpatient context service users reported experiencing abuse and discrimination because of their race, both during their treatment and in society in general (Solanki et al. 2023). In the same paper, a service user also described being perceived as stronger, and subjected to harsher treatment by staff due to being a Black man. In the Dutch forensic setting, ethnicity was described as one of several characteristics that made it more likely that a service user would be targeted by peers in a confrontative, violent manner (Verstegen et al., 2022). | No or very minor concerns | Moderate concerns | Minor concerns | Moderate concerns | Low |
| **Inequality of access** | Ability or lack of ability of the services to provide appropriate and timely care for service users irrespective of their demographic or medical background. | 4<br>Bendelow et al., 2019, Goodall et al., 2019; Sondhi et al., 2018, Yu et al., 2022 | Several newly identified studies contained accounts of service users receiving insufficient treatment or being unable to access care in a timely manner due to their age, gender, demographic or personal characteristics, or medical history. Examples included delayed service entry due to having an additional addiction diagnosis (Bendelow et al., 2019; Sondhi et al., 2018), young people's mental health services not well placed to support young women with experiences of past trauma/abuse (Bendelow et al., 2019), or wishing to see more diverse staff to facilitate better communication, e.g. people of all genders in the police services (Goodall et al., 2019). | No or very minor concerns | Moderate concerns | No or very minor concerns | Moderate concerns | Low |
| **7 Pathway to admission** |  |  |  |  |  |  |  |  |
| **Pathway to admission** | The experience of service users in the lead up to assessment and involuntary admission. Their experience of the care pathway: was the admission necessary, or avoidable, whether alternatives were available and explored. | 7<br>Bendelow et al., 2019; McGuiness et al., 2018; Smyth et al., 2021; Solanki et al., 2023; Vallarino et al., 2019; Wormdahl et al., 2021, Yu et al., 2022 | This theme reflects service users' reflections on their experience of the care pathway leading to assessment or involuntary treatment, e.g. whether it was felt that the admission was necessary or avoidable, and whether alternatives were available and explored. Some reported a lack of access or availability of services that would be less restrictive than hospital admissions. Others did not feel they received the right support from their GP, community or early intervention service despite seeking help, leading to symptoms getting worse, and resulting in admission or readmission. For example, police involvement and | No or very minor concerns | Minor concerns | No or very minor concerns | No or very minor concerns | High |

##### 1 Emotional impact

Involuntary assessments and admissions were reported (35, 36, 37, 38, 39, 40, 41, 42) to be met on occasions with *acceptance* where this involved accessing support and a safe environment. At the same time, the *impact of detention* (32, 33, 35, 36, 39, 40, 41, 42, 43, 44, 45, 46, 47, 48, 49, 50, 51, 52, 53), or more specifically the *impact of coercive treatment* was presented (34, 37, 41, 42, 44, 48, 50) as a typically negative, traumatic experience for many. *Feelings following discharge* appeared to be mixed, with reported difficulties with coping in the community and managing mental health post-discharge (33, 37, 39, 41, 47, 48, 49, 51).

From papers included in this review, *Therapeutic benefit* emerged as a new subtheme (33, 34, 37, 39, 40, 41, 42, 45, 48, 49, 54). This consists of service users’ views on whether their admission was necessary, helpful for their recovery or not. These reports reflected on views of treatment as un-therapeutic and traumatic, and in some cases, even counterproductive in terms of mental health:

> *‘…that’s the thing, it makes you feel worse afterwards than you did before. I’m sitting here, I’m more depressed and stressed coming out of that, and freaked out, than I was going in before’* (*33*)

Other papers reflected on some positive experiences, for example on receiving care that is needed which contributed to managing better following admissions, as illustrated by the following reflection on a ‘Living with Psychosis’ programme delivered during admission:

> *‘Living With uh Psychosis and stuff was really beneficial . . . ‘Cause it gave a lot of discussion time, and a lot of facts. And rather than just sort of sugar-coating stuff and dumbing it down, they were happy to answer questions and air anything you wanted to discuss, so that was empowering’* (*49*)

##### 2 Impact on self-worth

In the sub-theme *Power,* several studies (32, 33, 36, 40, 42, 43, 44, 47, 48, 49, 50, 52) reported on how admission affected people’s ability to have an influence on their progress and treatment, and the common feeling of disempowerment due to depending on staff to access basic necessities on a day to day basis, and ward routines. It was reported (33, 34, 37, 39, 41, 42, 44, 45, 50, 51, 52, 53) that involuntary assessment and admission often led to feeling *dehumanised:*

> *‘It’s just one big black hole that assessment room they keep you in, nothing, no information as to what is going to happen, by when and who is doing it. Dump you in the room to be stared at like some sort of strange animal.’* (*51*)

The detention process was seen (32, 33, 34, 35, 39, 40, 41, 42, 44, 45, 47, 48, 51, 53, 55) as contributing to *stigma,* for example through being perceived less credible or as dangerous by others (e.g. in daily life, or staff on the ward) due to having a mental health condition, or being seen to be taken and/or handcuffed by police. Some studies reported on people’s experiences of positively changing perceptions of mental ill-health but this tended to be single respondents rather than the majority view (35, 41). Social proximity of others, and recovery-based and meaningful activities whilst on the ward were reported (32, 33, 34, 37, 39, 49, 50, 54, 55) as having potentially *positive impacts* on self-esteem and independence.

##### 3 Information and involvement in care

The balance between *coercion, consent and choice* was varied, with many accounts (32, 33, 34, 36, 37, 39, 40, 41, 42, 43, 44, 45, 48, 49, 50, 54) reporting service users being offered little choice or opportunity to meaningfully consent, with fewer reports of collaborative care. Healthcare staff providing information and explaining legal processes, being given better access to advocacy and other forms of representation, where available, enabled service users to exercise their *rights* and navigate complex legal processes according to some of the primary studies (36, 39, 41, 44, 49, 52).

Several papers also reported that *information on what’s happening* (32, 36, 39, 40, 42, 43, 44, 45, 47, 50, 51) and *involvement in treatment decisions* (32, 33, 34, 35, 36, 39, 41, 42, 43, 45, 49, 50) were important aspects of service users’ admission experiences. Others suggested (33, 34, 36, 39, 40, 41, 42, 45, 49, 50) the importance of the way *medication* was discussed, explained and administered.

##### 4 Quality of relationships

Positive and negative experiences were both present in reports containing reflections on the relationship with *police and emergency department staff* (32, 34, 35, 36, 44, 48, 51, 53), and *inpatient staff* (33, 34, 35, 36, 39, 42, 43, 44, 45, 48, 50, 52, 54). Approachable, caring, skilled communication was seen as valued in some reports, whilst experiencing inadequate, forceful, bullying behaviour from professionals was also often described. *Family and friends* were described as a source of emotional support and advocacy at times, although service user accounts of tension, distrust and being misinterpreted during and after admission were also often reported (33, 34, 35, 39, 42, 43, 45, 47, 48, 49, 50). Similarly, *other service users* could be a source of peer-support and positive social interaction, although there were also reports of potential conflict and violent confrontations, a source of fear (33, 34, 36, 40, 47, 49, 50, 52, 55). Some of the papers in this review (33, 39, 47, 48, 50, 52, 55) also described how coping with various aspects of involuntary treatment often led to people *‘Playing ball*’, a concept initially described in one of the studies (48) and included as a new sub-theme. This covers instances of people adapting their communication towards staff and others, e.g. becoming more guarded in discussing mental health symptoms openly, and increasingly self-regulating and adhering to ward routines to keep to a minimum the threat of coercion, confrontation, and lengthy admission:

> *‘when the doctor asked them were they still hearing the voices. In their head they’d say yeah, but then like they’d be saying no . . . Sometimes I say I’m better than I am . . . but sometimes . . . I’m not 100%, that’s all . . .They just keep you in for longer . . . Unless you’re right completely like, they just lock you up.’* (*39*)

##### 5 Quality of the environment

The importance of quality of environment in *police cells* (35, 44, 50, 51), *hospital wards* (33, 34, 36, 39, 40, 48, 50, 54), *forensic wards* (49, 52) were discussed by several papers. It was highlighted that many aspects of the material environment (e.g. being chaotic, distressing, prison-like) needed significant improvement before being seen as a temporary place of safety or part of a therapeutic environment:

> *‘In psychiatry, there are conditions that need a lot of improvement and especially in closed psychiatry. So when you’re in there, it’s really terrible that the door is closed and you’re not allowed out. I wasn’t allowed out for six to seven weeks and I walked up and down like a tiger in a cage, and I found it terrible and I find it terrible every time.’* (50)

Some identified studies suggested (33, 47, 53, 54) that *meaningful activities* were valued and seen as promoting recovery and reducing tension and boredom but were not always accessible. *Personal safety and security* were reported (33, 35, 41, 50, 52, 54) as key expectations during involuntary admissions, but experiences of aggression and assault were detailed in a number of studies.

##### 6 Discrimination

Studies in this review reported service users’ experiences of different forms of discrimination in more detail then in the previous review of service users’ experiences of detention (24). Hence this concept is discussed as a separate, new overarching theme. Related sections were coded into two subthemes, *racial discrimination* and *equality of access*.

###### Racial discrimination

Whilst only a few studies explicitly addressed this topic, experiences of discrimination based on race and ethnicity have been reported from three separate contexts. These included a paper on young adults’ views on police involvement in detention for psychiatric assessments (32), one on the experiences of service users of a Black ethnic background admitted to hospital under the Mental Health Act in the UK (34), and one taking place in a Dutch forensic setting (52). In terms of police involvement in US, Florida, the original paper reported accounts of police conduct seen as disrespectful in general, but also a more specific experience of being treated differently specifically because of one’s race (32). In the UK inpatient context, the included study reported experience of abuse and discrimination because of race, both during their treatment and in society in general (34). In the same paper, a service user’s account also described being perceived as stronger, and subjected to harsher treatment by staff due to their race:

> *‘I mean we all know, there’s no point kidding ourselves, this is generally a racist country :: : from my experience of fifteen years of having a mental illness. Being black, you are treated as if you’re superhuman, you’ve got superhuman powers ::: you just get treated differently because you’re black. They [staff] assume because you’re black that you’re stronger :: : you can take it.’* (34)

In the Dutch forensic setting, ethnicity was described as one of several characteristics that made it more likely that a service user would be targeted by peers in a confrontative, violent manner (52).

###### Inequality of access

Several studies in this review contained accounts of service users receiving insufficient treatment or being unable to access care in a timely manner prior to or during compulsory admissions due to their age, gender, demographic or personal characteristics, or medical history. These related to receiving treatment from a community service, as well as accessing mental health assessment or place of safety at a time of need. Examples included difficulties in accessing after-care due to age (41), wishing to see more diverse staff to facilitate better communication, e.g. people of all genders in the police services (44), delayed service entry due to having an additional addiction diagnosis (35, 51), or young people’s mental health services not well placed to support young women with experiences of past trauma/abuse:

*Younger women were very critical of support from Child and Adolescent Mental Health Services, especially with regard to sexual abuse, and nearly all the women with this history felt that statutory adult mental health services were unable to offer the help they needed to manage their dissociative episodes or address the traumas underpinning their mental-health problems: ‘[My community mental-health team] are underresourced, and in my most recent meeting with them, I was told that if I’m in crisis, the only option is to call the police!’* (35)

##### 7 Pathway to admission

This theme reflects service users’ experiences of the care pathway leading to assessment or involuntary treatment. It includes instances when primary studies reported experiences that admission was unnecessary or avoidable, and whether alternatives were available and explored, or to the contrary, when compulsory admission occurred because a lack of care in the community (34, 35, 39, 40, 41, 48, 53). As seen below, some studies reported a lack of access or availability of services that would be less restrictive than hospital admissions, whilst other papers reflected on experiences in which not receiving the right support (e.g. from primary physician, a community or early intervention, service) despite seeking help, led to symptoms getting worse, resulting in admission or readmission. For example, police involvement and detention followed due to inadequate responses from A&E and health emergency call centres (35), insufficient knowledge of low-threshold services by primary care physicians, or insufficient capacity of specialist outpatients’ services to support those with serious mental health issues (53). These challenges with accessing timely crisis support in the community which might help prevent the need for detention were exacerbated for people from a range of minoritised groups, as described in the *Inequality of access* sub-theme above. The following account describes how mental health crises escalate due to lack of sufficient support from primary or community services:

> *‘Months ago when I approached my general practitioner and I said to him that I was feeling depressed, I should have got help then. Rather than when it becomes too late, so that’s where I feel I’ve been let down: I think, at that time, I feel he should have taken it more seriously.’* (34)

### Carers

#### Overview of included studies

We included eight papers with carer participants. However, the following carer results are largely derived from six studies: three that interviewed carers only (56, 57, 58) and three with mixed samples of carers, service users and other stakeholders, who were interviewed or in focus groups (41, 45, 53). Two studies with mixed samples (35, 51), both about experiences of detention with police powers and/or accessing police place of safety in England, included only one or no reference to carers in their results.

Carer study characteristics are reported in Table 3. The number of carer participants in all eight papers ranged from 3 (51) to 21 (53). The carers were in Australia (Brisbane), England, Germany, Norway, Republic of Ireland, South Korea, and the USA (Connecticut). All had experience of caring for a family member.

#### Thematic Synthesis

We coded the results sections of the included studies against a framework created to represent the overarching themes and implicit subthemes of our previous carer review (25). The new data fitted this framework and were coded to all the deductively derived sub-themes except one, Hope. Because of the consistency of the new data with the previous review no new overarching themes were developed inductively. However, we have fresh examples, from the perspectives of the new studies, to illustrate the five main themes, and we have identified some new sub-themes. The words in bold relate to sub-theme headings listed in Table 5.

**Table 5.**
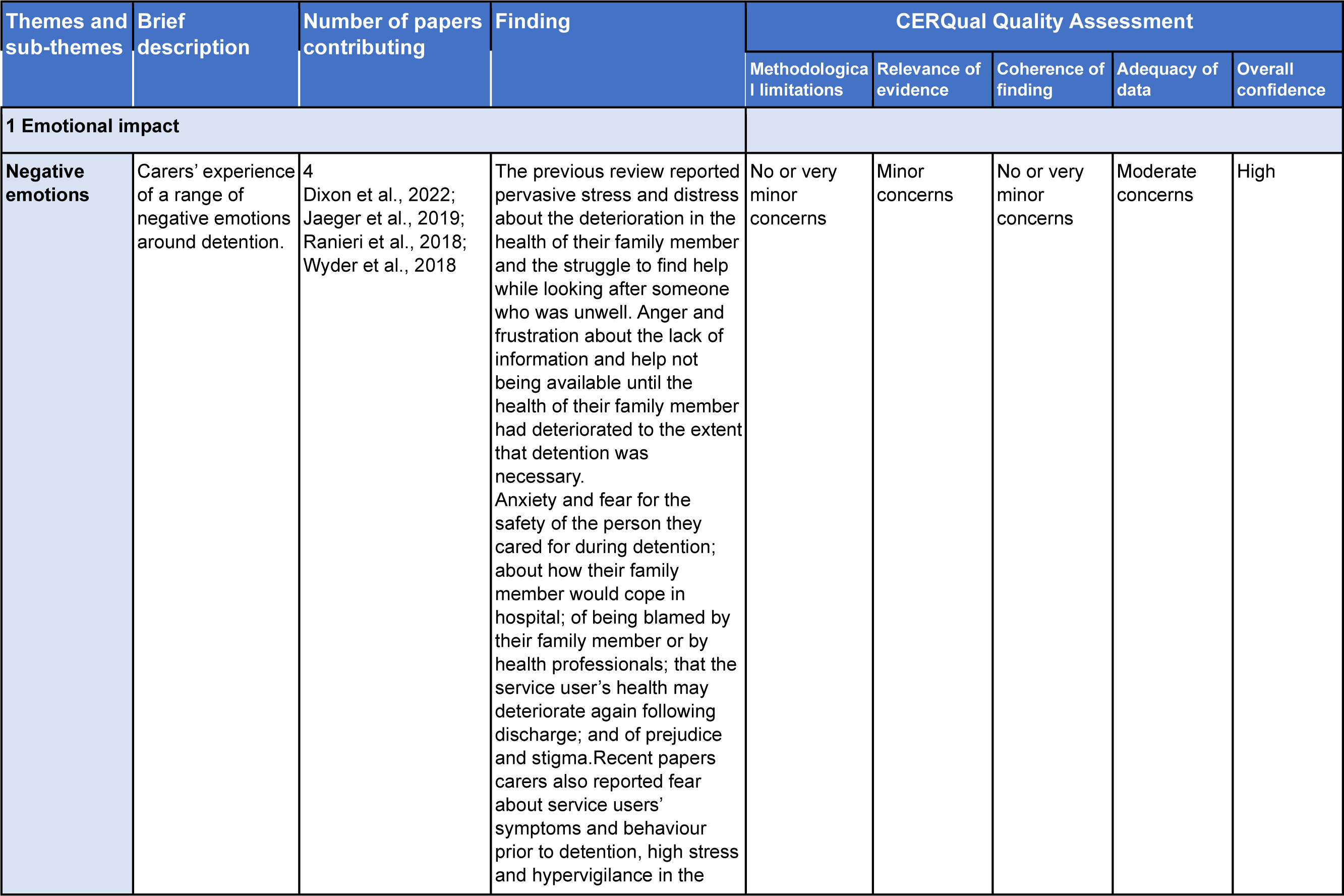
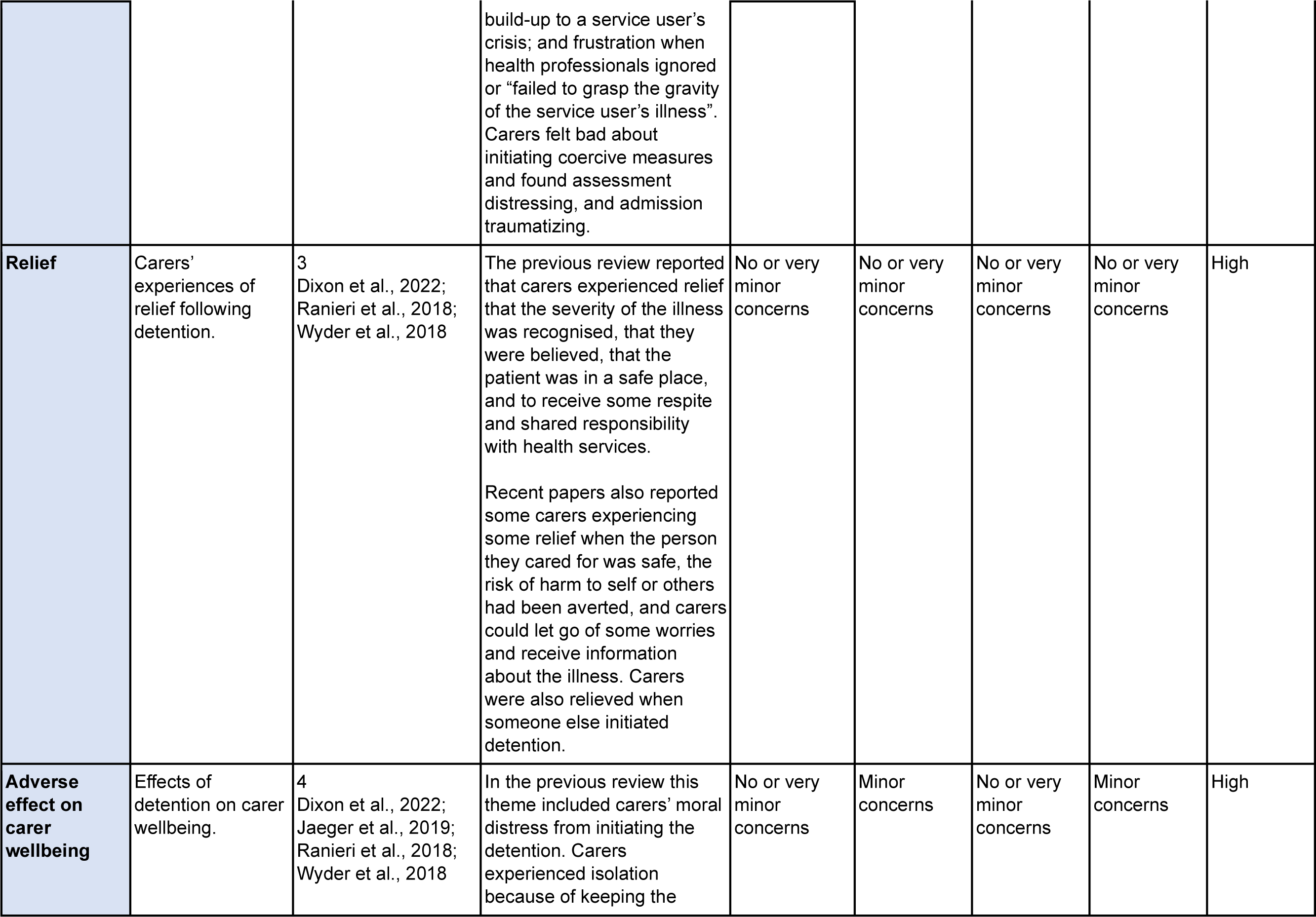
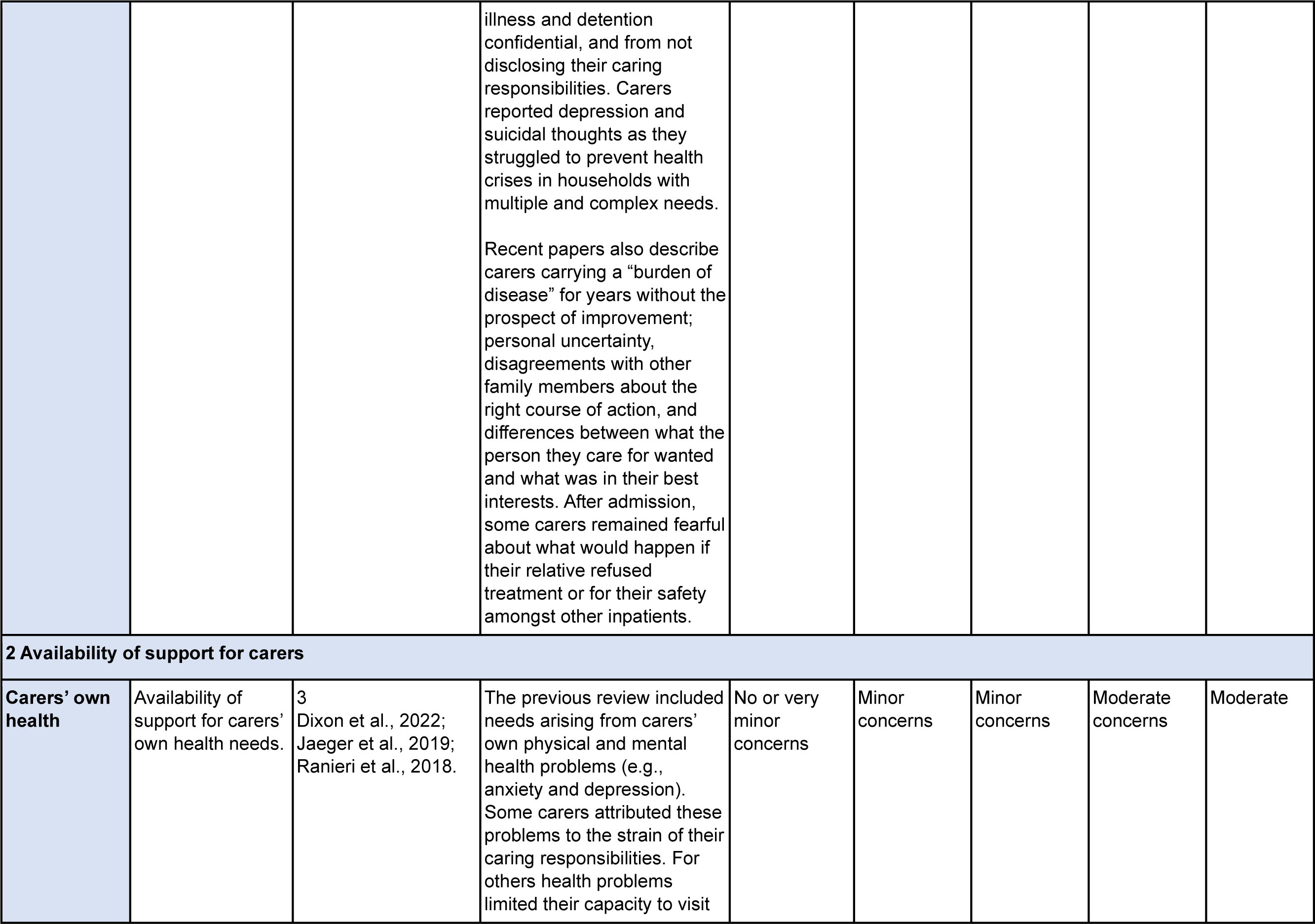
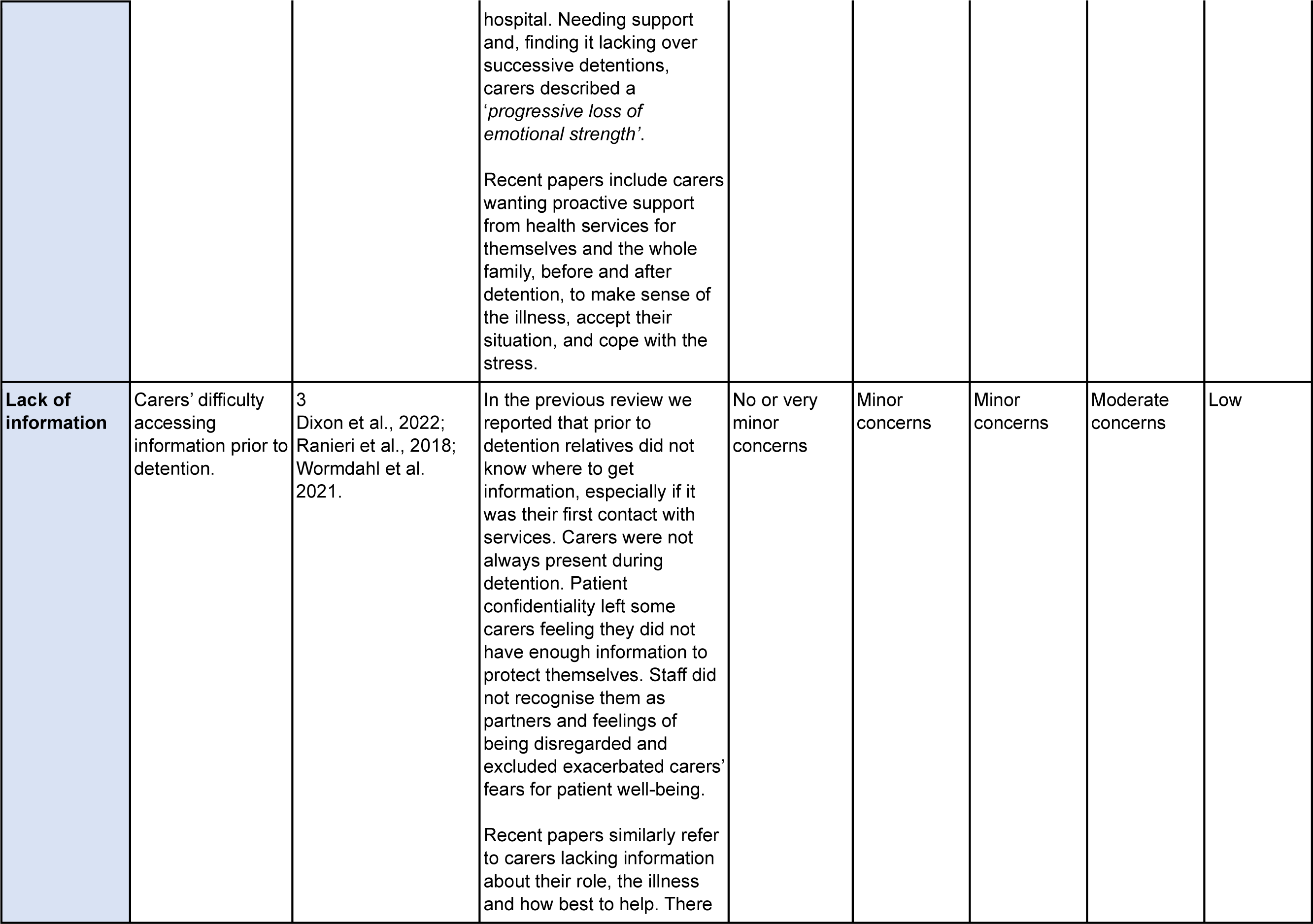
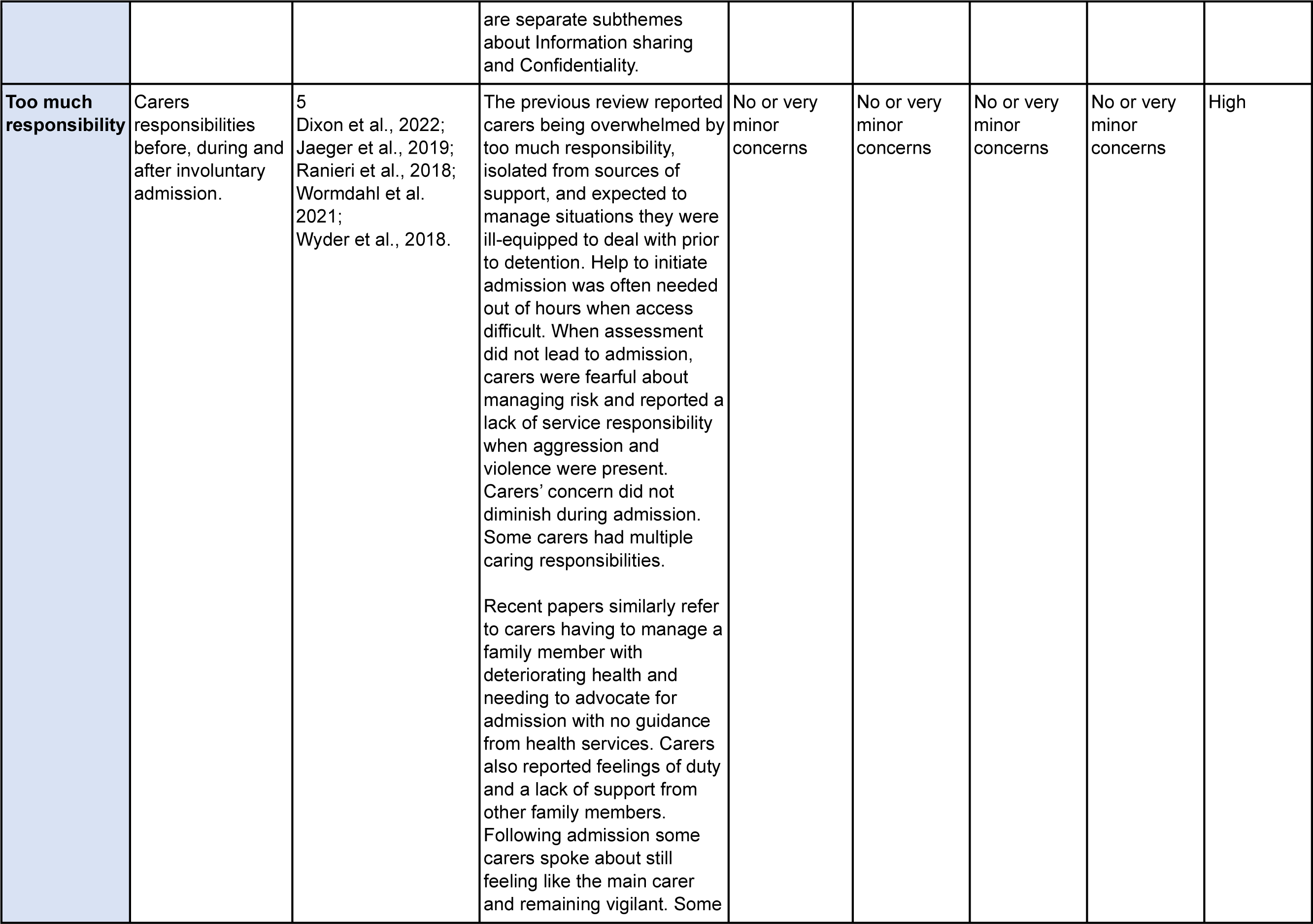
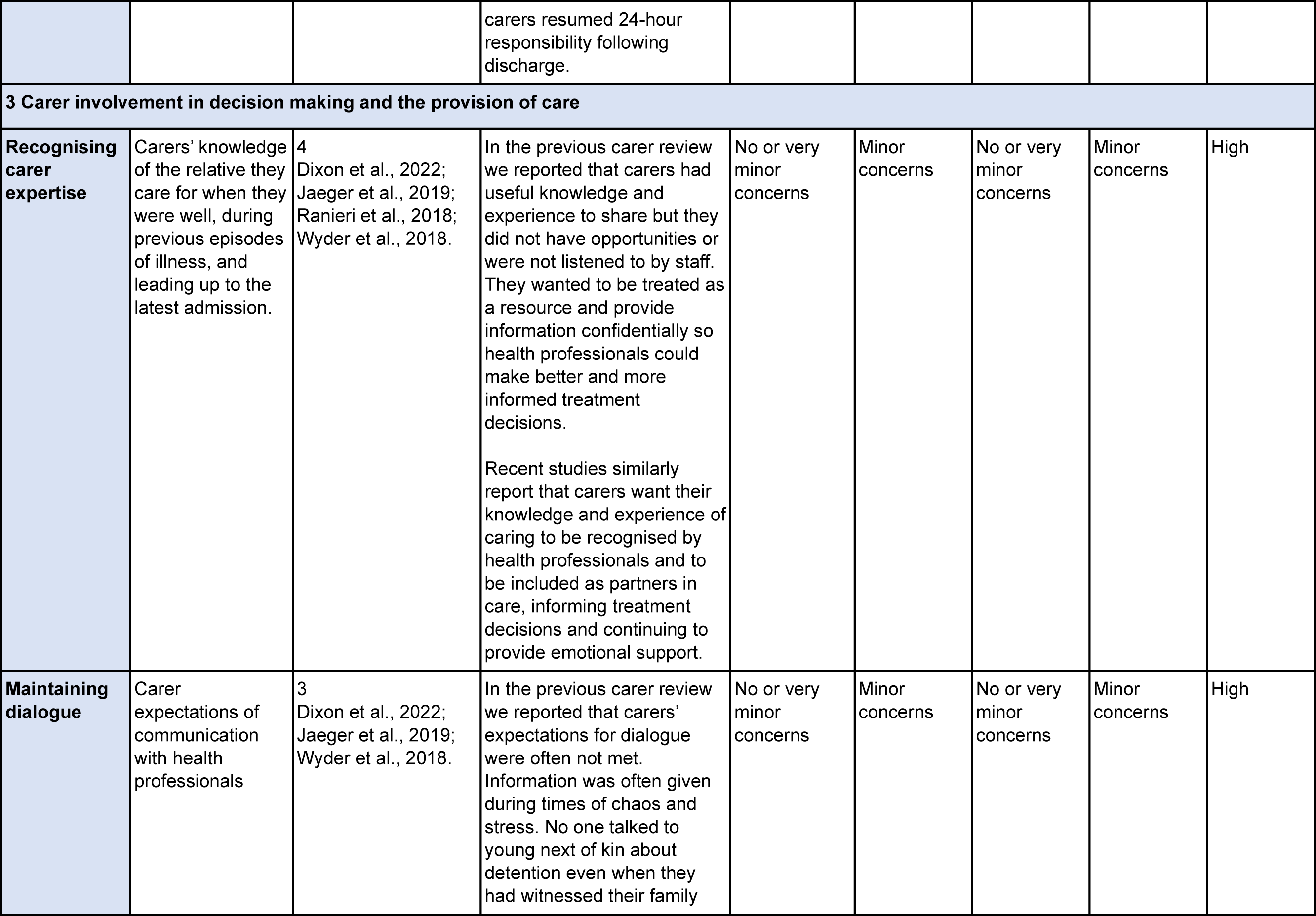
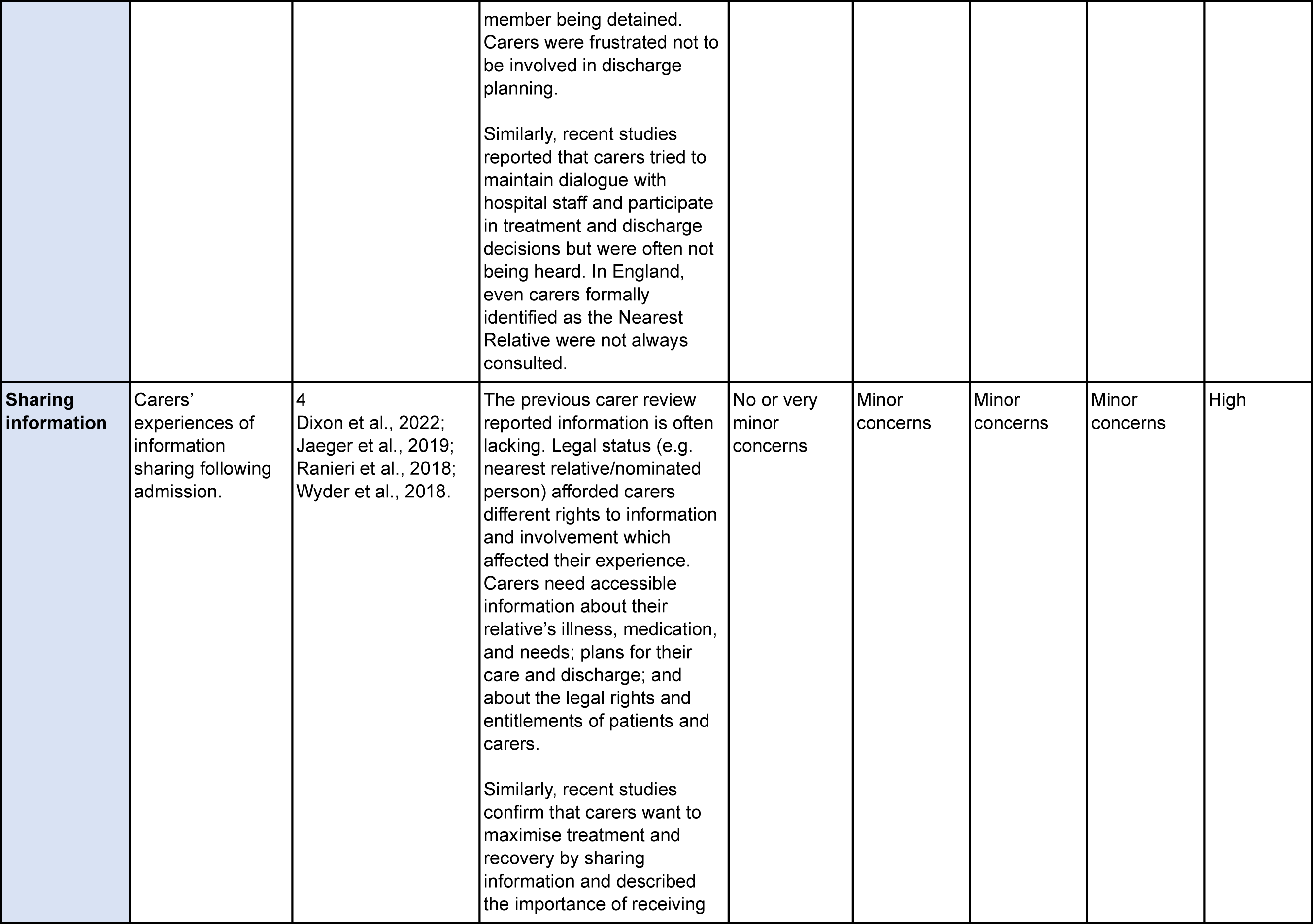
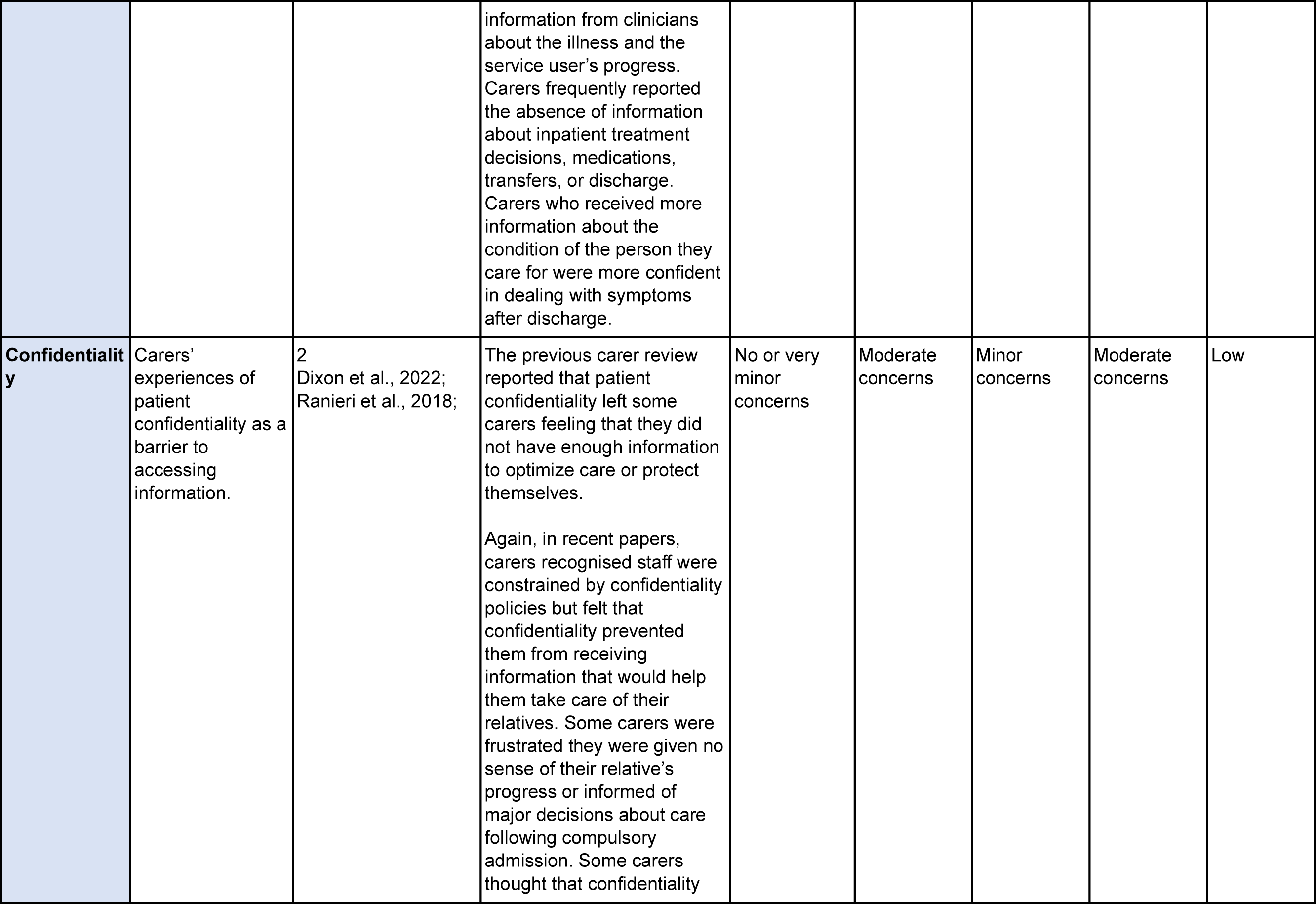
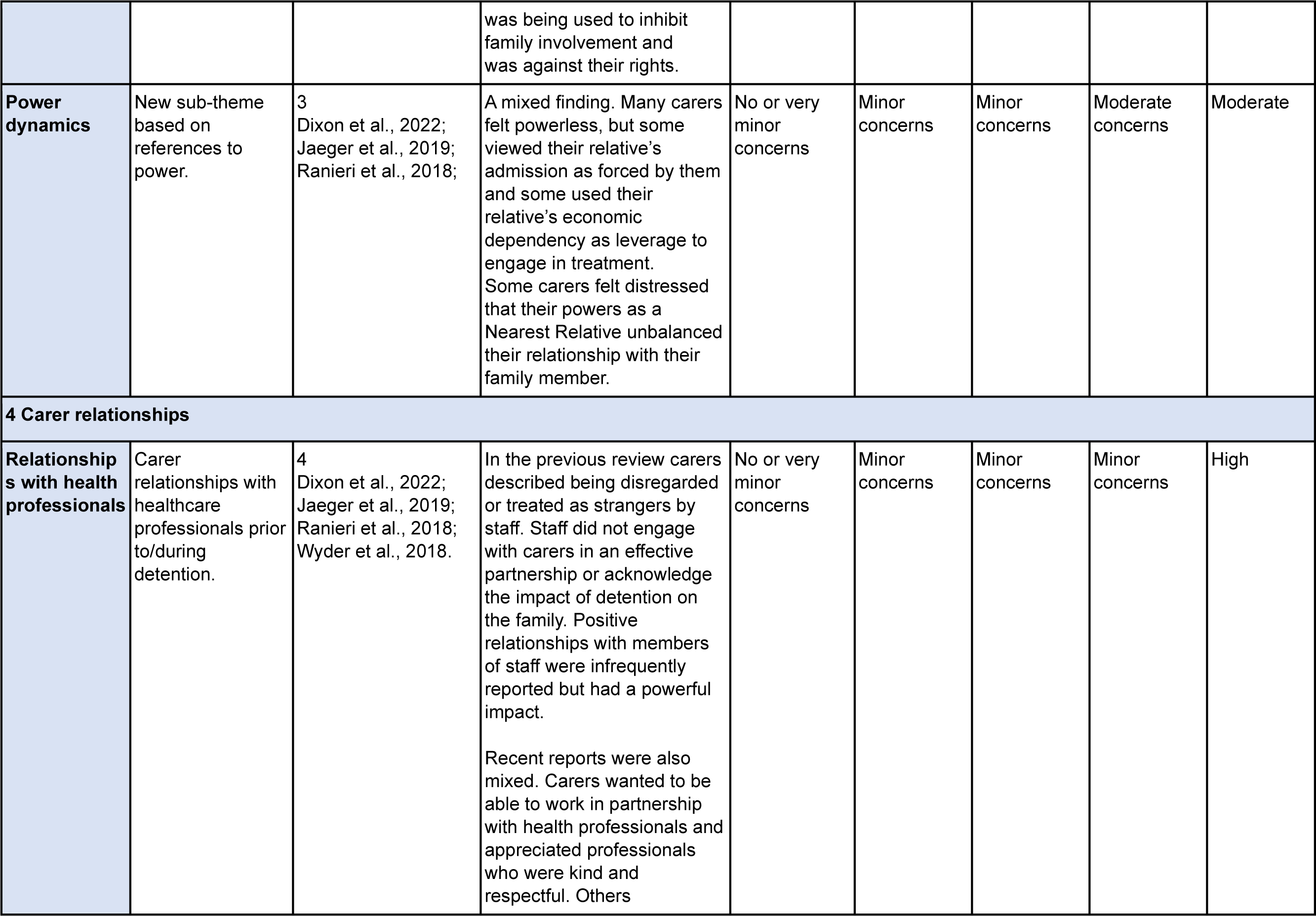
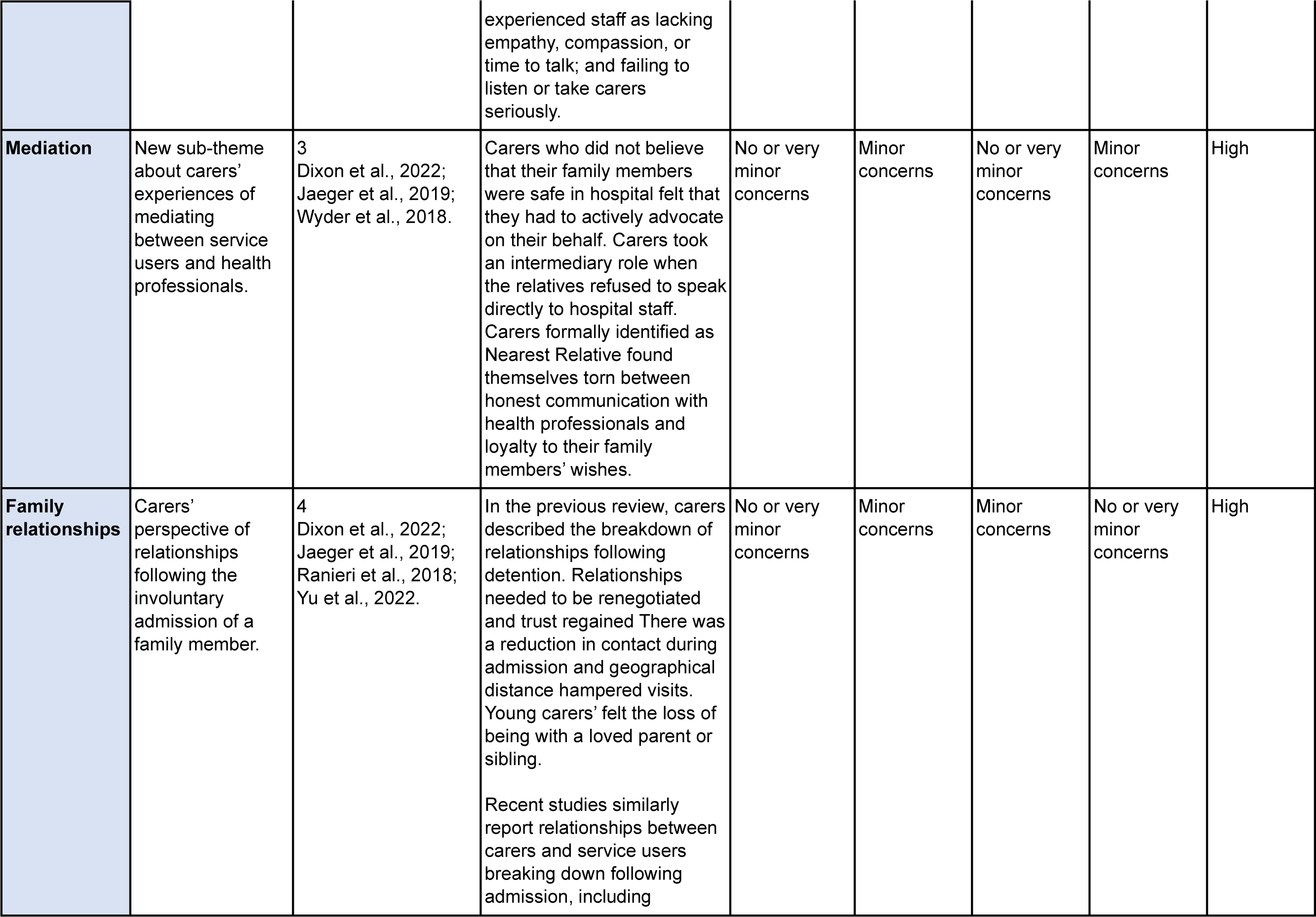
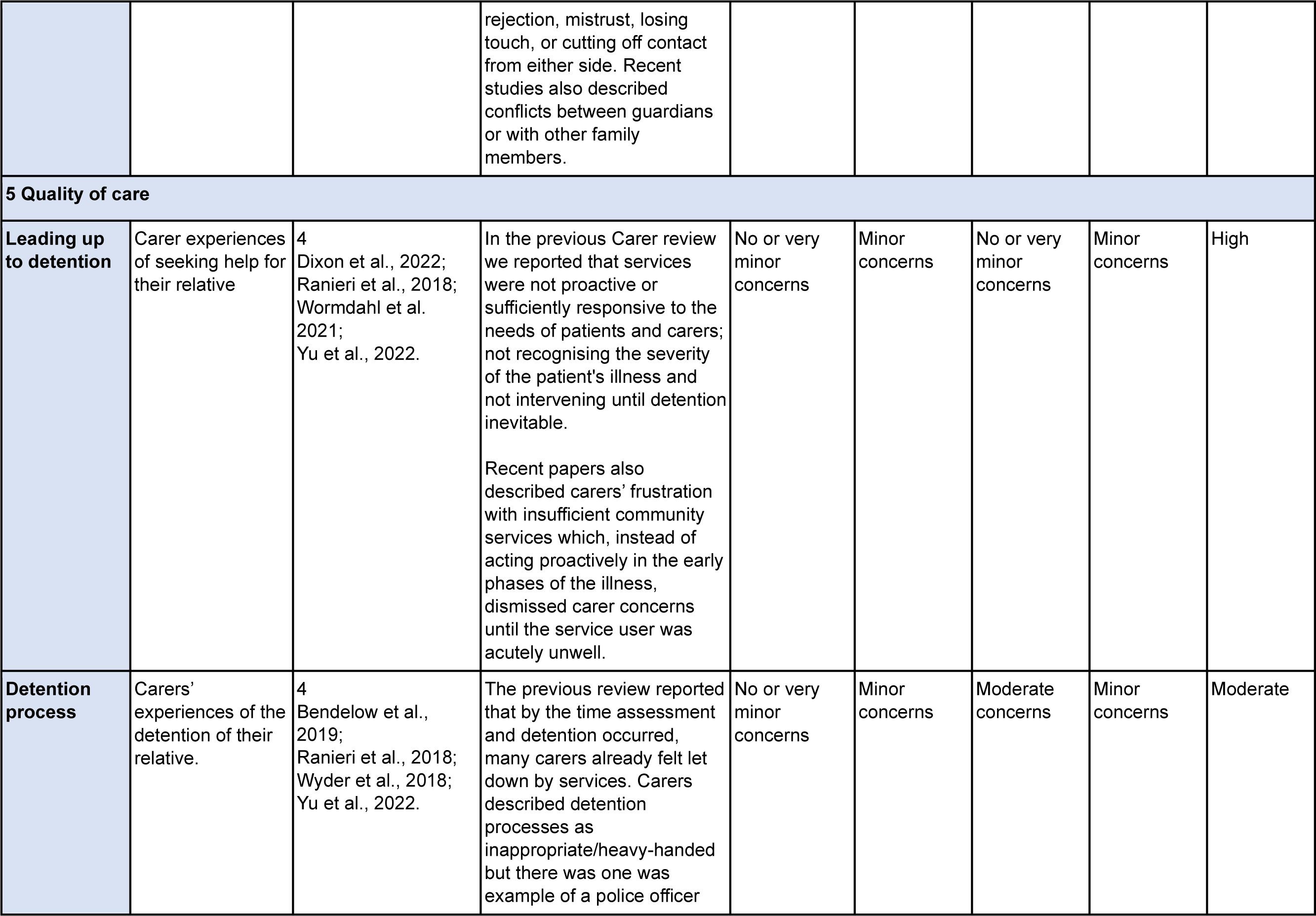
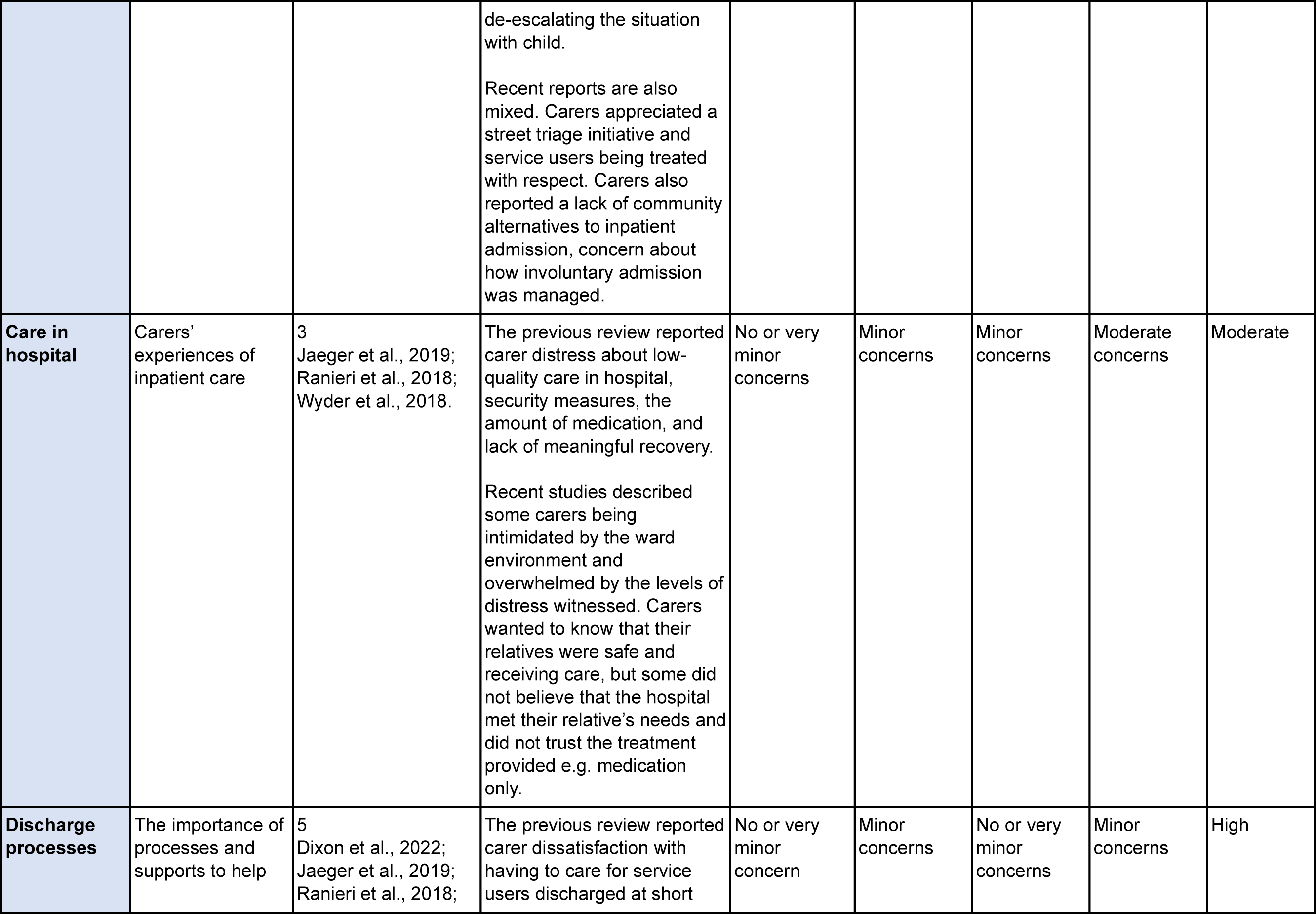
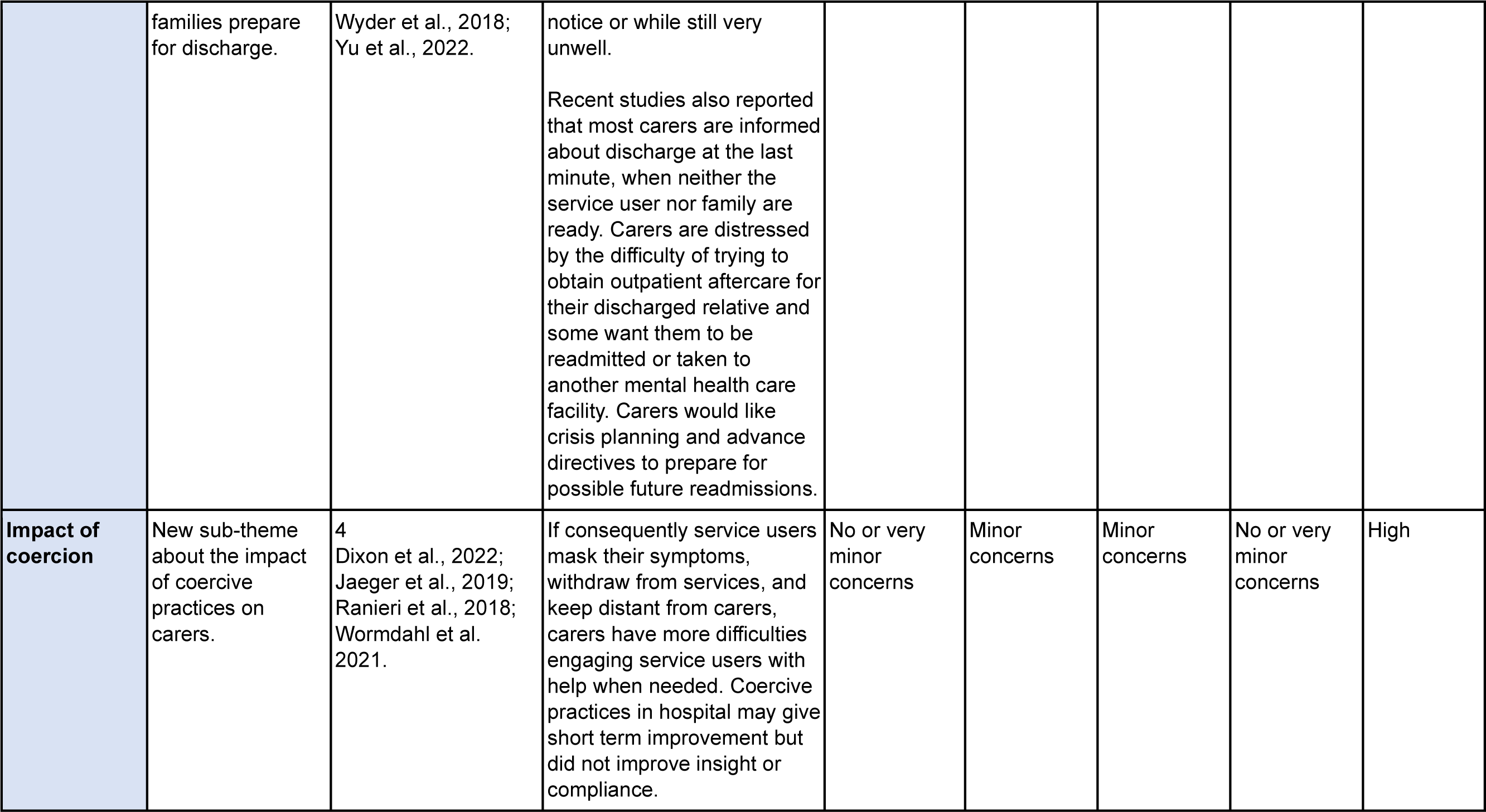
Summary of findings: Thematic framework, carer themes, sub-themes, certainty of evidence assessment.

| Themes and sub-themes | Brief description | Number of papers contributing | Finding | CERQual Quality Assessment |  |  |  |  |
| --- | --- | --- | --- | --- | --- | --- | --- | --- |
|  |  |  |  | Methodological limitations | Relevance of evidence | Coherence of finding | Adequacy of data | Overall confidence |
| 1 Emotional impact |  |  |  |  |  |  |  |  |
| Negative emotions | Carers' experience of a range of negative emotions around detention. | 4<br>Dixon et al., 2022;<br>Jaeger et al., 2019;<br>Ranieri et al., 2018;<br>Wyder et al., 2018 | The previous review reported pervasive stress and distress about the deterioration in the health of their family member and the struggle to find help while looking after someone who was unwell. Anger and frustration about the lack of information and help not being available until the health of their family member had deteriorated to the extent that detention was necessary.<br>Anxiety and fear for the safety of the person they cared for during detention; about how their family member would cope in hospital; of being blamed by their family member or by health professionals; that the service user's health may deteriorate again following discharge; and of prejudice and stigma. Recent papers carers also reported fear about service users' symptoms and behaviour prior to detention, high stress and hypervigilance in the build-up to a service user's crisis; and frustration when health professionals ignored or "failed to grasp the gravity of the service user's illness". Carers felt bad about initiating coercive measures and found assessment distressing, and admission traumatizing. | No or very minor concerns | Minor concerns | No or very minor concerns | Moderate concerns | High |
| **Relief** | Carers' experiences of relief following detention. | 3<br>Dixon et al., 2022;<br>Ranieri et al., 2018;<br>Wyder et al., 2018 | The previous review reported that carers experienced relief that the severity of the illness was recognised, that they were believed, that the patient was in a safe place, and to receive some respite and shared responsibility with health services.<br>Recent papers also reported some carers experiencing some relief when the person they cared for was safe, the risk of harm to self or others had been averted, and carers could let go of some worries and receive information about the illness. Carers were also relieved when someone else initiated detention. | No or very minor concerns | No or very minor concerns | No or very minor concerns | No or very minor concerns | High |
| **Adverse effect on carer wellbeing** | Effects of detention on carer wellbeing. | 4<br>Dixon et al., 2022;<br>Jaeger et al., 2019;<br>Ranieri et al., 2018;<br>Wyder et al., 2018 | In the previous review this theme included carers' moral distress from initiating the detention. Carers experienced isolation because of keeping the<br>illness and detention confidential, and from not disclosing their caring responsibilities. Carers reported depression and suicidal thoughts as they struggled to prevent health crises in households with multiple and complex needs.<br>Recent papers also describe carers carrying a “burden of disease” for years without the prospect of improvement; personal uncertainty, disagreements with other family members about the right course of action, and differences between what the person they care for wanted and what was in their best interests. After admission, some carers remained fearful about what would happen if their relative refused treatment or for their safety amongst other inpatients. | No or very minor concerns | Minor concerns | No or very minor concerns | Minor concerns | High |
| **2 Availability of support for carers** |  |  |  |  |  |  |  |  |
| **Carers’ own health** | Availability of support for carers’ own health needs. | 3<br>Dixon et al., 2022; Jaeger et al., 2019; Ranieri et al., 2018. | The previous review included needs arising from carers’ own physical and mental health problems (e.g., anxiety and depression). Some carers attributed these problems to the strain of their caring responsibilities. For others health problems limited their capacity to visit<br>hospital. Needing support and, finding it lacking over successive detentions, carers described a *'progressive loss of emotional strength'*.<br>Recent papers include carers wanting proactive support from health services for themselves and the whole family, before and after detention, to make sense of the illness, accept their situation, and cope with the stress. | No or very minor concerns | Minor concerns | Minor concerns | Moderate concerns | Moderate |
| **Lack of information** | Carers' difficulty accessing information prior to detention. | 3<br>Dixon et al., 2022; Ranieri et al., 2018; Wormdahl et al. 2021. | In the previous review we reported that prior to detention relatives did not know where to get information, especially if it was their first contact with services. Carers were not always present during detention. Patient confidentiality left some carers feeling they did not have enough information to protect themselves. Staff did not recognise them as partners and feelings of being disregarded and excluded exacerbated carers' fears for patient well-being.<br>Recent papers similarly refer to carers lacking information about their role, the illness and how best to help. There<br>are separate subthemes about Information sharing and Confidentiality. | No or very minor concerns | Minor concerns | Minor concerns | Moderate concerns | Low |
| **Too much responsibility** | Carers responsibilities before, during and after involuntary admission. | 5<br>Dixon et al., 2022; Jaeger et al., 2019; Ranieri et al., 2018; Wormdahl et al. 2021; Wyder et al., 2018. | The previous review reported carers being overwhelmed by too much responsibility, isolated from sources of support, and expected to manage situations they were ill-equipped to deal with prior to detention. Help to initiate admission was often needed out of hours when access difficult. When assessment did not lead to admission, carers were fearful about managing risk and reported a lack of service responsibility when aggression and violence were present. Carers' concern did not diminish during admission. Some carers had multiple caring responsibilities.<br>Recent papers similarly refer to carers having to manage a family member with deteriorating health and needing to advocate for admission with no guidance from health services. Carers also reported feelings of duty and a lack of support from other family members. Following admission some carers spoke about still feeling like the main carer and remaining vigilant. Some<br>carers resumed 24-hour responsibility following discharge. | No or very minor concerns | No or very minor concerns | No or very minor concerns | No or very minor concerns | High |
| **3 Carer involvement in decision making and the provision of care** |  |  |  |  |  |  |  |  |
| **Recognising carer expertise** | Carers' knowledge of the relative they care for when they were well, during previous episodes of illness, and leading up to the latest admission. | 4<br>Dixon et al., 2022; Jaeger et al., 2019; Ranieri et al., 2018; Wyder et al., 2018. | In the previous carer review we reported that carers had useful knowledge and experience to share but they did not have opportunities or were not listened to by staff. They wanted to be treated as a resource and provide information confidentially so health professionals could make better and more informed treatment decisions.<br>Recent studies similarly report that carers want their knowledge and experience of caring to be recognised by health professionals and to be included as partners in care, informing treatment decisions and continuing to provide emotional support. | No or very minor concerns | Minor concerns | No or very minor concerns | Minor concerns | High |
| **Maintaining dialogue** | Carer expectations of communication with health professionals | 3<br>Dixon et al., 2022; Jaeger et al., 2019; Wyder et al., 2018. | In the previous carer review we reported that carers' expectations for dialogue were often not met. Information was often given during times of chaos and stress. No one talked to young next of kin about detention even when they had witnessed their family<br>member being detained. Carers were frustrated not to be involved in discharge planning.<br>Similarly, recent studies reported that carers tried to maintain dialogue with hospital staff and participate in treatment and discharge decisions but were often not being heard. In England, even carers formally identified as the Nearest Relative were not always consulted. | No or very minor concerns | Minor concerns | No or very minor concerns | Minor concerns | High |
| **Sharing information** | Carers' experiences of information sharing following admission. | 4<br>Dixon et al., 2022; Jaeger et al., 2019; Ranieri et al., 2018; Wyder et al., 2018. | The previous carer review reported information is often lacking. Legal status (e.g. nearest relative/nominated person) afforded carers different rights to information and involvement which affected their experience. Carers need accessible information about their relative's illness, medication, and needs; plans for their care and discharge; and about the legal rights and entitlements of patients and carers.<br>Similarly, recent studies confirm that carers want to maximise treatment and recovery by sharing information and described the importance of receiving<br>information from clinicians about the illness and the service user's progress. Carers frequently reported the absence of information about inpatient treatment decisions, medications, transfers, or discharge. Carers who received more information about the condition of the person they care for were more confident in dealing with symptoms after discharge. | No or very minor concerns | Minor concerns | Minor concerns | Minor concerns | High |
| **Confidentiality** | Carers' experiences of patient confidentiality as a barrier to accessing information. | 2<br>Dixon et al., 2022;<br>Ranieri et al., 2018; | The previous carer review reported that patient confidentiality left some carers feeling that they did not have enough information to optimize care or protect themselves.<br>Again, in recent papers, carers recognised staff were constrained by confidentiality policies but felt that confidentiality prevented them from receiving information that would help them take care of their relatives. Some carers were frustrated they were given no sense of their relative's progress or informed of major decisions about care following compulsory admission. Some carers thought that confidentiality<br>was being used to inhibit family involvement and was against their rights. | No or very minor concerns | Moderate concerns | Minor concerns | Moderate concerns | Low |
| **Power dynamics** | New sub-theme based on references to power. | 3<br>Dixon et al., 2022;<br>Jaeger et al., 2019;<br>Ranieri et al., 2018; | A mixed finding. Many carers felt powerless, but some viewed their relative's admission as forced by them and some used their relative's economic dependency as leverage to engage in treatment. Some carers felt distressed that their powers as a Nearest Relative unbalanced their relationship with their family member. | No or very minor concerns | Minor concerns | Minor concerns | Moderate concerns | Moderate |
| **4 Carer relationships** |  |  |  |  |  |  |  |  |
| **Relationships with health professionals** | Carer relationships with healthcare professionals prior to/during detention. | 4<br>Dixon et al., 2022;<br>Jaeger et al., 2019;<br>Ranieri et al., 2018;<br>Wyder et al., 2018. | In the previous review carers described being disregarded or treated as strangers by staff. Staff did not engage with carers in an effective partnership or acknowledge the impact of detention on the family. Positive relationships with members of staff were infrequently reported but had a powerful impact.<br>Recent reports were also mixed. Carers wanted to be able to work in partnership with health professionals and appreciated professionals who were kind and respectful. Others experienced staff as lacking empathy, compassion, or time to talk; and failing to listen or take carers seriously. | No or very minor concerns | Minor concerns | Minor concerns | Minor concerns | High |
| **Mediation** | New sub-theme about carers' experiences of mediating between service users and health professionals. | 3<br>Dixon et al., 2022; Jaeger et al., 2019; Wyder et al., 2018. | Carers who did not believe that their family members were safe in hospital felt that they had to actively advocate on their behalf. Carers took an intermediary role when the relatives refused to speak directly to hospital staff. Carers formally identified as Nearest Relative found themselves torn between honest communication with health professionals and loyalty to their family members' wishes. | No or very minor concerns | Minor concerns | No or very minor concerns | Minor concerns | High |
| **Family relationships** | Carers' perspective of relationships following the involuntary admission of a family member. | 4<br>Dixon et al., 2022; Jaeger et al., 2019; Ranieri et al., 2018; Yu et al., 2022. | In the previous review, carers described the breakdown of relationships following detention. Relationships needed to be renegotiated and trust regained There was a reduction in contact during admission and geographical distance hampered visits. Young carers' felt the loss of being with a loved parent or sibling.<br>Recent studies similarly report relationships between carers and service users breaking down following admission, including rejection, mistrust, losing touch, or cutting off contact from either side. Recent studies also described conflicts between guardians or with other family members. | No or very minor concerns | Minor concerns | Minor concerns | No or very minor concerns | High |
| **5 Quality of care** |  |  |  |  |  |  |  |  |
| **Leading up to detention** | Carer experiences of seeking help for their relative | 4<br>Dixon et al., 2022;<br>Ranieri et al., 2018;<br>Wormdahl et al. 2021;<br>Yu et al., 2022. | In the previous Carer review we reported that services were not proactive or sufficiently responsive to the needs of patients and carers; not recognising the severity of the patient's illness and not intervening until detention inevitable.<br>Recent papers also described carers' frustration with insufficient community services which, instead of acting proactively in the early phases of the illness, dismissed carer concerns until the service user was acutely unwell. | No or very minor concerns | Minor concerns | No or very minor concerns | Minor concerns | High |
| **Detention process** | Carers' experiences of the detention of their relative. | 4<br>Bendelow et al., 2019;<br>Ranieri et al., 2018;<br>Wyder et al., 2018;<br>Yu et al., 2022. | The previous review reported that by the time assessment and detention occurred, many carers already felt let down by services. Carers described detention processes as inappropriate/heavy-handed but there was one was example of a police officer<br>de-escalating the situation with child.<br>Recent reports are also mixed. Carers appreciated a street triage initiative and service users being treated with respect. Carers also reported a lack of community alternatives to inpatient admission, concern about how involuntary admission was managed. | No or very minor concerns | Minor concerns | Moderate concerns | Minor concerns | Moderate |
| **Care in hospital** | Carers' experiences of inpatient care | 3<br>Jaeger et al., 2019;<br>Ranieri et al., 2018;<br>Wyder et al., 2018. | The previous review reported carer distress about low-quality care in hospital, security measures, the amount of medication, and lack of meaningful recovery.<br>Recent studies described some carers being intimidated by the ward environment and overwhelmed by the levels of distress witnessed. Carers wanted to know that their relatives were safe and receiving care, but some did not believe that the hospital met their relative's needs and did not trust the treatment provided e.g. medication only. | No or very minor concerns | Minor concerns | Minor concerns | Moderate concerns | Moderate |
| **Discharge processes** | The importance of processes and supports to help | 5<br>Dixon et al., 2022;<br>Jaeger et al., 2019;<br>Ranieri et al., 2018; | The previous review reported carer dissatisfaction with having to care for service users discharged at short | No or very minor concern | Minor concerns | No or very minor concerns | Minor concerns | High |
|  | families prepare for discharge. | Wyder et al., 2018; Yu et al., 2022. | notice or while still very unwell.<br>Recent studies also reported that most carers are informed about discharge at the last minute, when neither the service user nor family are ready. Carers are distressed by the difficulty of trying to obtain outpatient aftercare for their discharged relative and some want them to be readmitted or taken to another mental health care facility. Carers would like crisis planning and advance directives to prepare for possible future readmissions. |  |  |  |  |  |
| **Impact of coercion** | New sub-theme about the impact of coercive practices on carers. | 4<br>Dixon et al., 2022; Jaeger et al., 2019; Ranieri et al., 2018; Wormdahl et al. 2021. | If consequently service users mask their symptoms, withdraw from services, and keep distant from carers, carers have more difficulties engaging service users with help when needed. Coercive practices in hospital may give short term improvement but did not improve insight or compliance. | No or very minor concerns | Minor concerns | Minor concerns | No or very minor concerns | High |

##### 1 Emotional impact of detention

Carers experienced a range of conflicting emotions around detention.

###### Negative emotions

Carers reported fear about service users’ symptoms and behaviour prior to detention (56, 57), high stress and hypervigilance in the build-up to a service user’s crisis (58); and frustration when health professionals ignored or ‘*failed to grasp the gravity of the service user’s illness’* (56). Carers felt bad about initiating coercive measures (45) and found assessment distressing (56), and admission traumatizing (57).

> *‘The actual sectioning process was about as horrible as it ever could have been. It was possibly the worst experience of my life…’* (carer, England, (56))

###### Relief

Following admission, some carers felt some relief because they were reassured that the risk of harm to self or others had been averted, and the service user was now safe and in receipt of care (56, 57, 58). Other examples of relief arose from receiving information about the illness (58); being named as Nearest Relative (a defined close relative in the English Mental Health Act whom clinicians have a duty to consult during the process of assessment for compulsory admission) rather than another family member (56); and when someone else initiated detention (57).

> *‘A member of the public called in once. I felt relieved it wasn’t us that time. When he recovered, we could face him and tell him we had nothing to do with it.’* (carer, Ireland, (57)

###### Adverse effect on carer wellbeing

Carers talked about carrying a ‘*burden of disease’* for years without the prospect of improvement (45). Personal uncertainty, disagreements with other family members about the right course of action, and differences between what the person they cared for wanted and what was in their best interests, all took a toll on carer wellbeing (56, 57).

> *‘As I’m leaving he’s crying out, “Mom, why you doing this to me? I don’t want to be here, don’t leave me here.”’* (mother, Ireland, (57))Carers were fearful about what would happen if the service user refused treatment (Jaeger) and, in contrast to the relief following admission reported above, some carers remained fearful for their relative’s safety amongst other inpatients (58).

##### 2 Availability of support for carers

###### Carers own health

For their own health, carers need support to be offered proactively, before, during and after detention, to help them make sense of the illness, deal with stress, and accept their situation (45, 56).

> *‘One minute there were police cars and half a dozen doctors and lots of shouting and kind of stuff going on and then the next minute I was just here on my own and that was a bit kind of challenging, difficult.’* [Carer participant, (56)]One carer suggested involving the whole family as a ‘*more well-rounded approach to recovery’* because the whole family can be affected by the illness and detention (57).

###### Lack of information

Carers reported that there was a lack of information and guidance from health services as they managed a family member with deteriorating health and sought help (53, 57).

> *‘The whole [assessment] experience was pretty traumatic really I suppose. There should be more support actually for me or actually tell me what I need to do to support him.’* (carer, England, (56))Other sub-themes below, Information sharing and Confidentiality, reflect the provision of information following involuntary admission.

###### Too much responsibility

Carer participants expressed a sense of duty towards their family member which they willingly took on (56). While some carers initiated or advocated for admission (45), some also remained vigilant to protect their relative following admission and resumed 24-hour responsibility following discharge (58). For example, several service users in one study depended on their carer as their only social network (53).

> *‘The phone calls, oh, they’ve taken an overdose or I’m going to go under a bus or— they [health professionals] don’t seem to have any real regard for the fact that there are people out here that are having to deal with all this stuff.’* (carer, Brisbane, (58))

##### 3 Carer involvement in decision making and care

###### Recognising carer expertise

Carers’ familiarity with the relative they care for, having been with them when they were well and during previous episodes of illness, and knowing the circumstances of the latest admission, gives carers expertise (45, 56, 57).

> ‘*He’s been with me all his life*.’ (participant 9, (56))

> *‘I know what she was like before.’* (sister, (45))Carers said that they want their knowledge and experience of caring to be recognised and valued by health professionals and to be included as partners in care, informing treatment decisions and continuing to provide emotional support (57, 58).

> *‘Participants described the importance of the treating team recognising the critical role they played in their relative’s life. Many reported being treated as a nuisance.’* (58)

###### Maintaining dialogue

Following involuntary admission carers reported that they tried to maintain dialogue with hospital staff and participate in treatment and discharge decisions but were often not being heard (45, 58). In England, even carers formally identified as the Nearest Relative were not always consulted. This left them uncertain about the purpose of the role and feeling it was not taken seriously (56).

> *‘Nearest Relatives are not routinely consulted or provided with information once the hospital admission takes place.’* (56), England)

###### Sharing information

To maximise treatment and recovery, carers want to share information and described the importance of receiving information from clinicians about the illness and the service user’s progress. Carers frequently reported the absence of information about inpatient treatment decisions, medications, transfers, or discharge, even when the service user was being discharged to their care (45, 56, 57, 58).

> *‘While the treatment team relied on them for information, they were not informed about their relatives’ progress in return.’* (58)

Carers who received more information about the condition of the person they care for were more confident in dealing with symptoms after discharge (58).

###### Confidentiality

Like carers in the previous review (25), carers in recent studies recognised that staff were constrained by service users’ rights to confidentiality, but expressed frustration that they were given no sense of their family member’s progress during the period following involuntary admission to hospital (56). Carers stated that confidentiality prevented them from receiving information that would help them take care of their relative (57).

> *‘Some disclosed that this [confidentiality] clause was being used purposively to inhibit family involvement. Furthermore, a subsection of these caregivers stated that being denied access to such information went against their rights.’* (57)

###### Power dynamics

Carers described experiences of power in their role. For example, leveraging a service user’s economic dependency to make them engage in treatment (45), or initiating the detention.

> *‘Caregivers predominantly viewed their relative’s admission as forced and imposed by them rather than led by the service user or mental health services.’* (57)

Some carers felt distressed that their powers as a Nearest Relative (to request a mental health assessment or discharge) unbalanced their relationship with their relative and hoped that the imbalance would be temporary (56). Many carers felt an overall sense of powerlessness as they supported their family member through episodes of illness.

##### 4 Carer relationships

###### Relationships with health professionals

Reports of the relationships between carers and health professionals were mixed. Some carers appreciated experiences of staff generosity, patience, and kindness towards service users (56, 57).

> *‘One participant praised the AMHPs [Approved Mental Health Professionals – clinicians involved in assessment for detention in hospital in England] she had spoken to as being ‘fair’ and ‘lovely’’* (56)

Other carers struggled to get medical attention in the community and spoke of health professionals failing to listen and lacking empathy. Following admission carers wanted to work in partnership with hospital staff, but some described staff as not having enough time to talk to them (45, 56, 57, 58).

###### Mediation

Some carers described mediating between the person they care for and health professionals. In Brisbane, carers who did not believe that their family members were safe in hospital felt that they had to actively advocate on their behalf.

> *‘If you haven’t got someone fighting from the outside inwards, well you’re just left to your own devices. […] If she did not have any visitors, she could be bruised and bullied.’* (carer, Brisbane, (58)).

In Germany, carers took an intermediary role, “*translating the perspectives of each side to the other”* when the relatives they cared for refused to speak directly to hospital staff (45). Carers in England who had the role of Nearest Relative found themselves torn between honest communication with health professionals and loyalty to their family members’ wishes (56).

###### Family relationships

Carers spoke of trying to make sense of the illness in the context of family relationships.

> *‘Some interviewees reflected on their family relationships. They suspected that this also played a role in the behaviour of their ill family member.’* (45)

In some instances, the failure of other family members (other than the carer) to recognise the illness was a barrier to seeking help (57).

As in the previous carer review (25), some studies described communication breaking down between carers and service users following involuntary admission: mistrust and rejection by service users; losing touch; or carers cutting off contact after admission (41, 45, 57).

In England, no carer participant with the role of Nearest Relative thought that the role should be given to anyone outside the family, even if their relationship was no longer a close one (56).

##### 5 Quality of care

###### Leading up to detention

Several studies describe insufficient community services which, instead of acting proactively in the early phases of the illness, dismissed carer concerns until the service user was acutely unwell (53, 56, 57). One participant was concerned that ‘*undue focus on the service users’ rights and wishes may have acted against their best interests in the long run’* (56).

Some carers reported that pathways to care were inadequate. In Norway, carers and other stakeholders said that GPs had neither the time to properly conduct mental health assessments nor sufficient knowledge of primary care alternatives to hospital admission (53). Another study (41) also describes a lack of community alternatives to admission in South Korea.

As well as lack of access to community crisis services, cumbersome processes for initiating involuntary admission were seen as a barrier to timely detention:

> ‘*the need for a full medical assessment in service users who are acutely mentally unwell and have a standing history of mental illness’*, and in Connecticut ‘*caregivers argued against the need for probate court hearings in order to obtain a commitment order’* (57).

Other systemic barriers to treatment were reported: poor collaboration between primary and secondary services in Norway (53); disagreements between public and private services in Ireland; and catchment areas for specialist services in Ireland and the United States (57).

###### Detention process

Carers and service users in England were appreciative of the ‘*swift, effective, and compassionate interventions’* made by a Street Triage pilot in Sussex: a car crewed by a police officer and a mental health nurse (35). Carers in Ireland said emergency departments were unsuitable for mental health assessments. ‘*Some caregivers reported that their relative was treated fairly and respectfully during the admission’* and others described the detention process ‘*as clinical and devoid of compassion’* (57). In Brisbane a carer expressed concern about how involuntary admission was managed:

> *‘I didn’t have problems with the involuntary treatment. Without the involuntary treatment […] how would she have gotten the treatment? But it’s just how the service provider carried it out [which] is probably the bigger issue.’* (carer, Brisbane, (58))

In South Korea, there was an example of delayed treatment because of the new detention requirement for the consent of two legal guardians (41).

###### Care in hospital

Some carers described being intimidated by the ward environment and overwhelmed by the levels of distress they witnessed. Carers wanted to know that their relatives are safe and receiving care to help them recover, but some carers did not believe that the hospital met their relative’s needs and did not trust the treatment provided. They thought their conceptualization of the problem to be treated differed to that of health professionals. Many carers were critical of the focus on and adequacy of medication without talking therapies and attributed ‘*impairments of functioning’* to medication (45, 58).

> *‘All she does is see a doctor once or twice a week. There’s no counsellor brought in […] She seriously needs to talk to somebody, not for 10 minutes, how you’re going, how you’re feeling, are you still seeing anything? That’s all she gets. She’s never actually sat down with anybody and just talked about anything.’* (carer, Brisbane, (58))

###### Discharge processes

Recent studies reported the importance of processes and supports to help carers prepare for discharge but, like the previous review (25), many carers reported that they were informed about discharge at the last minute when neither the service user nor family was ready (58).

> *‘No discharge plan or anything. They didn’t even explain the medications to us, how he would take it, what times to give it to him. Just said ‘Bye’. They gave us a bag of medication.’* (carer, Brisbane, (58))

Some carers did not believe that their relative was ready for discharge and were concerned about their own ability to cope with the service user’s ‘*continued impairment’* or the *‘additional burden’* when a service user who refuses medication is discharged without treatment. Some described the distressing difficulty of trying to obtain outpatient aftercare (45, 57, 58). In South Korea it was reported that:

> *‘family members often want the patient to be readmitted or taken to another mental health care facility, rather than living together.’* (41)

Carers in Connecticut and Ireland spoke of their desire for crisis planning and advance directives to prepare for possible future readmissions (57).

###### Impact of coercion

Service users’ experiences of being detained in public, or roughly handled, or being subjected to coercive measures during admission and treatment have consequential impacts on carers too. Carers had difficulties arising from their relatives’ subsequent fear and unwillingness to seek help and engage with services, withdrawing and keeping their distance from carers; being guarded and masking their symptoms during assessments (45, 53, 56, 57). In one study carers recognised that coercion might lead to short-term improvement but:

> ‘*the effect was unsustainable and had not changed the insight, and because of this experience, patients were even more negative regarding compliance with treatment recommendations*.’ (45)

### Certainty of evidence

Certainty of evidence has been reviewed and appraised for each sub-theme, reported separately for service user and carer findings in Tables 4. and 5. respectively. There were 28 service user sub-themes, in which we rated overall confidence as high (10), moderate (13), low (3), or very low (2). For the 19 carer sub-themes, we rated overall confidence as high (13), moderate (4), or low (2). Decisions for lowering confidences were most commonly due to minor or moderate concerns due to relevancy of evidence, coherence of finding, and/or adequacy of data, as documented in Tables 4. and 5.

## Discussion

### Main findings

In our two earlier reviews (24, 25) service users primarily reflected on inpatient experiences, whilst data from carers described struggles to find health service support prior to detention and following discharge. In the current update we found a greater overlap in content: both service user and carer data included reflections on experiences of pathways to admission, the extent of therapeutic benefit from inpatient treatment, the frequent experience of coercion and its consequences for future engagement with health services. Studies included in this update also report on recent developments in mental health practice such as crisis planning, advanced directives and street triage, and changes to mental health legislation.

Our findings suggest that the experience of involuntary treatment and compulsory admissions is an often predominantly negative, at times traumatic experience for service users and carers, not always achieving the expected therapeutic benefit. A variety of factors are reported to contribute to this, including the use of coercive practices, too much focus on pharmaceutical treatments, lack of access to psychological and other therapies, uncaring staff attitudes, or a lack of a calm, therapeutic ward environment. Compulsory admissions are often associated with experiences of coercion, and variable but often low levels of involvement in decision making processes, which can both affect the perceived effect of hospital treatment (13). Involuntary treatment may be a particular source of tension as staff are providing care to which service users may not have the capacity to consent or adamantly oppose. Thus, delivering acute mental health services in a collaborative, non-coercive manner in this environment may be a particular challenge.

Our findings indicate that reliance on the use of coercive methods can be therapeutically counterproductive. To minimise experiences of coercion, service users may be motivated to minimise their symptoms and difficulties in discussion with health professionals and family members. Experiences of coercion may in turn make it less likely for people to seek or engage with subsequent treatment, corresponding with other findings (e.g. (1)). Bad experiences of detention and coercive treatment impacts on carers too. If service users mask their symptoms, stay guarded and ‘keep their distance’ as a way of avoiding further bad experiences, it is more difficult for carers to get help for them when needed. In addition to an effect on therapeutic relationships, coercion may also affect family dynamics. Carers described a need for support for themselves at the moment of admission and over time as they assist their family member in managing their mental health for years.

Our synthesis also highlighted some positive experiences during compulsory treatment, for example feeling relief, receiving help to manage symptoms, or family members accessing support. The finding that service users’ views on therapeutic benefit were mixed mirrors previous findings with service users’ groups of varying diagnoses (e.g. (59, 60, 61). In our review, service users reflected positively on being listened to, experiencing flexibility, being offered options. At the same time, as in previous reports, the opposite was also commonly experienced: not being heard, and lacking control over treatment (62). Studies in our review highlighted how staff attitudes (e.g. being respectful, caring, communicating well, empowering) could have an impact on how people saw their treatment and the degree of coercion.

Service user and carer accounts also reflected on the importance of pathways leading to hospital admissions. Several instances were highlighted when either a lack of alternatives to inpatient services, or professionals’ lack of mental health awareness may have escalated a crisis and contributed to involuntary admissions. In contrast, efficient collaboration between services was also noted by participants as a positive experience. An example of this is increasing coordination between mental health and law enforcement services when responding to crises, which is seen as potentially reducing the use of place of safety and police custody facilities in the UK (63).

Some service user reports referred to experiences of racial discrimination, and other inequalities (e.g. due to age, dual diagnosis etc.) in accessing services. This finding is more explicit in our review of recent studies than in previous reviews (24, 25), although it is notable that only a few papers addressed or specifically investigated the experiences of people with marginalised ethnic backgrounds. We note that initiatives to reduce racial discrimination and imparity are seen as increasingly important (10, 46), and should be reflected in additional resources for research and changes to practice in this area.

Whilst our themes and subthemes reported here are focused on the experiences of formal compulsory admissions, we also noted some of the more recent reports (32, 45, 50) described instances when service users underwent treatment whilst technically voluntary service users, but due to fear of coercion and thus *de facto* detained.

### Implications for policy and practice

Although compulsory admissions are an inherently coercive and often aversive situation, service user and carer accounts suggest that there are ways of improving experience and therapeutic potential, and address other issues such as inequality and discrimination:

1. Power, agency and choice were highlighted as important aspects of compulsory admission experience. Increased focus on supporting decision making may be particularly effective, given that many involuntarily admitted service users may have capacity to consent (64), for example by initiatives to provide supported involvement in treatment decisions from early stages of compulsory admissions (65). Additionally, for people experiencing readmission, advance directives and similar tools may support discussion of previous treatment experiences, accessing early support, or reducing the use of coercive practices (46, 66). Evidence shows that care planning and advance statements are effective in reducing detentions (67, 68). Broader use of these initiatives could address this aspect of service user experiences, whilst research is also needed to understand their implementation, and how they work for marginalised groups who may need them most. Providing information in a timely manner during all key stages to the admission process can also have a positive, empowering effect on both service users and carers (2).

2. Service user and carer voices highlighted the importance of working with respectful, engaged, kind staff. This corresponds with earlier qualitative work identifying service users’ desired qualities in mental health staff (69), emphasising the importance of empathy, kindness, respect, effective communication and fostering hope. These qualities have also been identified as facilitators of meaningful service user and staff relationships, and thus supporting meaningful therapeutic engagement (70). Addressing staff shortages, and recruitment and retention of staff with suitable personal characteristics seems important, as it may ameliorate otherwise often negative treatment experiences in the context of compulsory admissions and frequent use of coercive means. Training for mental health and other emergency staff in de-escalation (10), or increasing primary care practitioners’ mental health awareness (71) may also improve access and the quality of initial contact with treatment providers.

3. Service users and carers both identified missed opportunities to avoid detentions. Community-based alternatives to admission such as crisis houses, crisis resolution teams and acute day services may be beneficial both for the individual and the wider community, and wider implementation may have potential to reduce involuntary admissions (5, 12). Both the quality and intensity of upstream community care and availability of community crisis alternatives have been linked to compulsory admission rates (7), highlighting the need for proactive, well-resourced community care. Initiatives to enhance both crisis planning and ongoing monitoring and support post-discharge are of high interest as a potentially effective means to prevent repeat detentions (72, 73).

4. Addressing any inequalities of access to services and reducing racial discrimination and disparities appears to be a key issue to address across several mental health systems (21, 22, 74, 75). Service users from minoritised ethnic groups reported experiences of racism and unfair treatment, thus efforts to promote equality, diversity and inclusion and anti-racist practice are needed, a recent example of which is the Patient and Carer Race Equality Framework in England and Wales (76).

### Implications for research

The identified literature reflected accounts and views of some, but not all groups of service users who undergo compulsory admissions. In addition to the low number of papers on experiences of people from ethnic minority backgrounds and young people, more research is needed to understand the experiences of other seldom-heard groups e.g. those with intellectual disabilities, autism, speech impairments, and/or in long-term inpatient care.

With the increase of community crisis service models in recent years (77, 78) we need more evaluation studies to determine which might be most effective in reducing admissions and detention. This could reduce the number of instances in which opportunities to avoid compulsory hospital admissions are missed, a possibility that was identified by papers we reviewed. A clear effect of such alternatives on involuntary admissions has, however, yet to be demonstrated (67), thus, refining such models to ensure a clear focus on people whose history or the severity of their difficulties suggests they are at risk of compulsory admissions is a potentially fruitful direction for service development and research.

Many of the expressed wishes of service users and carers summarised in this review – for clear communication and a chance to be heard, for fairness and respect and some choice and control – map on to the components of procedural justice (79). Procedural justice focuses on perceptions of perceived control and fairness in the process, rather than the outcomes of a situation. It is theorised as comprising four components: allowing citizens a voice; perceived neutrality in decision-making; demonstrating dignity and respect during interactions; and having trustworthy motives. In mental health inpatient contexts, perceived procedural justice is associated with less perceived coercion (80) and better therapeutic alliance with staff (81). Development and evaluation of interventions specifically designed to enhance procedural justice in the assessment and compulsory detention processes are of high interest. Recent examples of this include crisis planning and monitoring for detained patients initiated in hospital (72), or supporting involvement in treatment decisions from early stages of compulsory admissions (65).

### Strengths and limitations

We used robust systematic search methodologies and followed established guidelines both for newly published primary research and for synthesising results. We have included recent studies that allowed corroborating and refining findings from previous systematic reviews and identify relevant new themes. Our review also reported findings from service user and carer groups that were present less in previous works, for example people from marginalised ethnic backgrounds. Our team had a diverse set of skills to draw on including researchers with lived experience, which informed the depth of our analysis, interpretation, and writing of the manuscript.

The use of qualitative synthesis methods may have led to the loss of more nuanced information on service user and carer experiences available in individual primary studies. There were only a few new studies available on the compulsory admission experience of ethnic minorities, or young people. Similarly, only three papers focused specifically on carers’ experiences, suggesting a necessity of further research in these areas. Our review focused on the experience of compulsory admission under mental health legislation, thus it did not fully explore involuntary treatment experiences when formally voluntary service users encounter coercive practices and undergo forced treatment despite their status. We also acknowledge that lived experience involvement could have been heightened by involvement in the earlier stages of the project, for example when drafting the review protocol. Evaluation of certainty of evidence for pre-existing themes has been carried out on newer information only, as the use of GRADE-CERQual was not a PRISMA recommendation at the time of the original review.

## Conclusion

Findings from our updated qualitative synthesis suggest that service users’ and carers’ experiences of compulsory admission processes are varied, predominantly negative. The negative impact of coercive measures have been reported across most studies, and more recent literature also reflected experiences of racial discrimination, inequality of access, and quality community care being seen as an alternative to detention in hospital.

A staff approach to service users that is both collaborative and kind appears to be important even if in the context of significant coercion and/or in instances when the person does not have the capacity to take part in treatment decisions. Positive accounts reported suggest that the experiences of compulsory admissions are also improved by professionals doing their best to inform at all stages of the admission. Mental health staff can positively affect treatment experience by being kind, offering choices where they can even if the situation seems dire, and involving service users and carers proactively in treatment decisions as and when possible. Community alternatives of inpatient care may also contribute to and lead to better overall treatment experiences.

## Lived Experience Commentary

### by Karen Machin, Patrick Nyikavaranda and Tamar Jeynes

This revised review builds upon two previous reviews by the same team from 2018. These earlier reviews, enriched by lived experiences commentaries, identified a pressing need for enhancing patient and carer experiences. They also highlighted the sluggish pace of translating knowledge into practical action. Regrettably, this latest review offers minimal new insights. However, it does underscore the counterproductive nature of over-reliance on coercive methods in care. The review acknowledges that while some may find solace in any support, it’s deeply troubling that issues like racial discrimination, unequal access, and general dissatisfaction have become more pronounced in recent years.

Notably, research by Morris et al. (82) suggests that it can take up to 17 years for research findings to be implemented in clinical practice. We cannot continue to wait. Urgent action is needed to enhance the experience of detention and, or even, dare we say it, earlier supportive interventions. This could address the immediate needs of individuals and potentially reduce the frequency of detentions.

As authors, we might argue that one way to promote change might be for researchers, funders, policymakers, and practitioners to **finally** act on what people with lived experience tell them they need. The current review suggests several improvements, including the provision of community-based alternatives to detention and the availability of family support. Additionally, it highlights the critical need for genuine cultural awareness training, moving beyond mere procedural compliance.

The numerous television exposes and media reports depicting the harrowing experiences within inpatient wards have understandably deterred many from seeking voluntary support. It is disheartening that this systematic review merely reiterates what service users and carers have long known: significant change is imperative. Perhaps the time has come for service users and carer groups to be more active in setting research agendas and conducting research themselves.

## Supporting information

GRADE-CERQual protocol

## Data Availability

Additional data is available on request via the corresponding author.

## Acknowledgements

We gratefully acknowledge the feedback and support provided by the members of the NIHR Mental Health Policy Research Unit’s Lived Experience Working Group in all stages of the report, including providing expert commentary, analysing and interpreting data, advising on terminology and reviewing and revising the manuscript.

## Availability of data

Thematic frameworks and illustrative quotes are available in Supplementary Materials. Additional data is available on request via the corresponding author.

## Abbreviations

Not applicable.

## Funding

This paper is based on independent research commissioned and funded by the National Institute for Health and Care Research Policy Research Programme, though the NIHR Policy Research Unit for Mental Health. The views expressed are those of the authors and not necessarily those of the NHS, the National Institute for Health and Care Research, the Department of Health and Social Care or its arm’s length bodies, and other Government Departments. The authors declare no completing interests.

## Author contributions

NA and RS searched the databases and registered the protocol. RS de-duplicated in EndNote and exported records to Rayyan. GBr, KS, NA, RS, SL, TJ did title and abstract screening and searched reference lists of included papers. AR & GBa searched citations. AR, GBa, GBr, KS, NA, RS and SL extracted data. AR, GBa, GBr, HG, NA, RS and SL did the quality appraisal. AS, BLE, GBa, NA, RS and UF agreed the coding frame. BLE, GBa, NA, RS and UF coded the included data. GBa and RS tabulated and summarised all service user and carer references. AS, AG, BLE, KP, PN, RS, TJ and UF discussed the findings. GBa, NA and RS graded the findings. RS drafted the carer results and GBa drafted all other sections of the manuscript, with input from RS and NA. All co-authors edited, revised and commented on the draft manuscript.

## Ethics

This systematic review and secondary data analysis (qualitative synthesis) has been conducted according to its pre-registered protocol as described in the manuscript.

## Supplementary information

### Search strategy 1 March 2023

#### Medline (OvidSP)

1. service-user* or patient* or consumer* or carer* or famil* or caregiver* or caregivers/ or relative* or inpatient* or client* or ((lived or life) adj experience*) or survivor*
2. "mental health act" or section* or ’mental treatment act’ or ((compuls* or involuntar* or coer* or forced or detention or detained or refusal or mandat* or civil or appeal* or advoc*) adj2 (hospital* or admiss* or admit* or readmiss* or commit* or assess* or treat* or healthcare))
3. mental disorders/ or ((mental* or psychologic* or psychiatr*) adj2 (health or disorder* or disease* or deficien* or illness* or problem*)).ti,ab,sh.
4. qualitative research/ or interview/ or qualitative or (theme$ or thematic) or ’ethnological research’ or (humanistic or existential or experiential or paradigm$) or (field adj (study or studies or research)) or ((purpos$ adj4 sampl$) or (focus adj group$)) or ’observational method$’ or ’content analysis’ or ((discourse$ or discurs$) adj3 analys?s) or ’narrative analys?s’ or (grounded adj (theor$ or analys?s)) or ’action research’ or (account or accounts or unstructured or openended or open ended or narrative$) or (lived adj experience$)
5. 1 and 2 and 3 and 4
6. limit 5 to yr=“2018-Current”

#### Embase (OvidSP)

1. service-user* or patient* or consumer* or carer* or famil* or caregiver* or caregiver/ or relative* or inpatient* or client* or ((lived or life) adj experience*) or survivor*)
2. "mental health act" or section* or ’mental treatment act’ or ((compuls* or involuntar* or coer* or forced or detention or detained or refusal or mandat* or civil or appeal* or advoc*) adj2 (hospital* or admiss* or admit* or readmiss* or commit* or assess* or treat* or healthcare))
3. mental disease/ or ((mental* or psychologic* or psychiatr*) adj2 (health or disorder* or disease* or deficien* or illness* or problem*)).ti,ab,sh.
4. qualitative research/ or interview/ or qualitative or (theme$ or thematic) or ’ethnological research’ or (humanistic or existential or experiential or paradigm$) or (field adj (study or studies or research)) or ((purpos$ adj4 sampl$) or (focus adj group$)) or ’observational method$’ or ’content analysis ’ or ((discourse$ or discurs$) adj3 analys?s) or ’narrative analys?s ’ or (grounded adj (theor$ or analys?s)) or ’action research’ or (account or accounts or unstructured or openended or open ended or narrative$) or (lived adj experience$)
5. 1 and 2 and 3 and 4
6. limit 5 to yr=“2018-Current”

#### PsycINFO (OvidSP)

1. service-user* or patient* or consumer* or carer* or famil* or caregiver* or caregivers/ or relative* or inpatient* or client* or ((lived or life) adj experience*) or survivor*
2. "mental health act" or section* or ’mental treatment act’ or ((compuls* or involuntar* or coer* or forced or detention or detained or refusal or mandat* or civil or appeal* or advoc*) adj2 (hospital* or admiss* or admit* or readmiss* or commit* or assess* or treat* or healthcare))
3. mental disorders/ or ((mental* or psychologic* or psychiatr*) adj2 (health or disorder* or disease* or deficien* or illness* or problem*)).ti,ab,sh.
4. qualitative research/ or interview/ or qualitative or (theme$ or thematic) or ’ethnological research’ or (humanistic or existential or experiential or paradigm$) or (field adj (study or studies or research)) or ((purpos$ adj4 sampl$) or (focus adj group$)) or ’observational method$’ or ’content analysis’ or ((discourse$ or discurs$) adj3 analys?s) or ’narrative analys?s’ or (grounded adj (theor$ or analys?s)) or ’action research’ or (account or accounts or unstructured or openended or open ended or narrative$) or (lived adj experience$)
5. 1 and 2 and 3 and 4
6. limit 5 to yr=“2018-Current”

#### HMIC (OvidSP)

1. service-user* or patient* or consumer* or carer* or famil* or caregiver* or carers/ or relative* or inpatient* or client* or ((lived or life) adj experience*) or survivor*
2. "mental health act" or section* or ’mental treatment act’ or ((compuls* or involuntar* or coer* or forced or detention or detained or refusal or mandat* or civil or appeal* or advoc*) adj2 (hospital* or admiss* or admit* or readmiss* or commit* or assess* or treat* or healthcare))
3. mental disorders/ or ((mental* or psychologic* or psychiatr*) adj2 (health or disorder* or disease* or deficien* or illness* or problem*)).ti,ab,sh.
4. qualitative research/ or interviews/ or qualitative or (theme$ or thematic) or ’ethnological research’ or (humanistic or existential or experiential or paradigm$) or (field adj (study or studies or research)) or ((purpos$ adj4 sampl$) or (focus adj group$)) or ’observational method$’ or ’content analysis’ or ((discourse$ or discurs$) adj3 analys?s) or ’narrative analys?s’ or (grounded adj (theor$ or analys?s)) or ’action research’ or (account or accounts or unstructured or openended or open ended or narrative$) or (lived adj experience$)
5. 1 and 2 and 3 and 4
6. limit 5 to yr=“2018-Current”

#### Social Science Citation Index (Web of Science)

1. service-user* or patient* or consumer* or carer* or famil* or caregiver* or relative* or inpatient* or client* or ((lived or life) N0 experience*) or survivor*
2. "mental health act" or section* or "mental treatment act" or ((compuls* or involuntar* or coer* or forced or detention or detained or refusal or mandat* or civil or appeal* or advoc*) N2 (hospital* or admiss* or admit* or readmiss* or commit* or assess* or treat* or healthcare))
3. mental disorders or ((mental* or psychologic* or psychiatr*) N2 (health or disorder* or disease* or deficien* or illness* or problem*))
4. qualitative research or interview or qualitative or (theme* or thematic) or "ethnological research" or (humanistic or existential or experiential or paradigm$) or (field N0 (study or studies or research)) or ((purpos* N4 sampl*) or (focus N0 group*)) or "observational method*" or "content analysis" or ((discourse* or discurs*) N3 analys?s) or "narrative analys?s" or (grounded N0 (theor* or analys?s)) or "action research" or (account or accounts or unstructured or openended or "open ended" or narrative*) or (lived N0 experience*)
5. 1 and 2 and 3 and 4
6. limit from 01/01/2018-01/03/2023

**Supplementary information, Table 6.** Prisma 2020 Checklist (Page et al., 2021)

| Section and Topic | Item # | Checklist item | Location where item is reported |
| --- | --- | --- | --- |
| **TITLE** |  |  |  |
| Title | 1 | Identify the report as a systematic review. | Title page (currently P1) |
| **ABSTRACT** |  |  |  |
| Abstract | 2 | See the PRISMA 2020 for Abstracts checklist. | Abstract (currently P1-2) |
| **INTRODUCTION** |  |  |  |
| Rationale | 3 | Describe the rationale for the review in the context of existing knowledge. | Background section, paragraphs 3-6 |
| Objectives | 4 | Provide an explicit statement of the objective(s) or question(s) the review addresses. | Background section, paragraph 6 |
| **METHODS** |  |  |  |
| Eligibility criteria | 5 | Specify the inclusion and exclusion criteria for the review and how studies were grouped for the syntheses. | Methods/Eligibility criteria |
| Information sources | 6 | Specify all databases, registers, websites, organisations, reference lists and other sources searched or consulted to identify studies. Specify the date when each source was last searched or consulted. | Methods/Data sources |
| Search strategy | 7 | Present the full search strategies for all databases, registers and websites, including any filters and limits used. | Supplementary information: Search strategy |
| Selection process | 8 | Specify the methods used to decide whether a study met the inclusion criteria of the review, including how many reviewers screened each record and each report retrieved, whether they worked independently, and if applicable, details of automation tools used in the process. | Methods/Selection process |
| Data collection process | 9 | Specify the methods used to collect data from reports, including how many reviewers collected data from each report, whether they worked independently, any processes for obtaining or confirming data from study investigators, and if applicable, details of automation tools used in the process. | Methods/Data Extraction |
| Data items | 10a | List and define all outcomes for which data were sought. Specify whether all results that were compatible with each outcome domain in each study were sought (e.g. for all measures, time points, analyses), and if not, the methods used to decide which results to collect. | Methods/Eligibility criteria, paragraph 3 |
|  | 10b | List and define all other variables for which data were sought (e.g. participant and intervention characteristics, funding sources). Describe any assumptions made about any missing or unclear information. | Methods/Data extraction, and Methods/Data analysis and synthesis |
| Study risk of bias assessment | 11 | Specify the methods used to assess risk of bias in the included studies, including details of the tool(s) used, how many reviewers assessed each study and whether they worked independently, and if applicable, details of automation tools used in the process. | Methods/Risk of bias and quality appraisal |
| Effect measures | 12 | Specify for each outcome the effect measure(s) (e.g. risk ratio, mean difference) used in the synthesis or presentation of results. | Methods/Data analysis and synthesis, paragraph 1 |
| Synthesis methods | 13a | Describe the processes used to decide which studies were eligible for each synthesis (e.g. tabulating the study intervention characteristics and comparing | Methods/Data analysis and synthesis, and study characteristics in Tables |
|  |  | against the planned groups for each synthesis (item #5)). | 2. and 3. |
|  | 13b | Describe any methods required to prepare the data for presentation or synthesis, such as handling of missing summary statistics, or data conversions. | Methods/Data analysis and synthesis |
|  | 13c | Describe any methods used to tabulate or visually display results of individual studies and syntheses. | Methods/Data analysis and synthesis, and Tables 4. and 5. |
|  | 13d | Describe any methods used to synthesize results and provide a rationale for the choice(s). If meta-analysis was performed, describe the model(s), method(s) to identify the presence and extent of statistical heterogeneity, and software package(s) used. | Methods/Data analysis and synthesis |
|  | 13e | Describe any methods used to explore possible causes of heterogeneity among study results (e.g. subgroup analysis, meta-regression). | Methods/Data analysis and synthesis, paragraph 2 |
|  | 13f | Describe any sensitivity analyses conducted to assess robustness of the synthesized results. | Methods/Assessment of confidence in findings |
| Reporting bias assessment | 14 | Describe any methods used to assess risk of bias due to missing results in a synthesis (arising from reporting biases). | NA for Thematic Synthesis |
| Certainty assessment | 15 | Describe any methods used to assess certainty (or confidence) in the body of evidence for an outcome. | Methods/Assessment of confidence in findings |
| **RESULTS** |  |  |  |
| Study selection | 16a | Describe the results of the search and selection process, from the number of records identified in the search to the number of studies included in the review, ideally using a flow diagram. | Results/Service users/Overview of included studies<br>Results/Carers/Overview of included studies<br>Figure 1.<br>Table 2. and Table 3. |
|  | 16b | Cite studies that might appear to meet the inclusion criteria, but which were excluded, and explain why they were excluded. | - |
| Study characteristics | 17 | Cite each included study and present its characteristics. | Results/Service users/Thematic synthesis<br>Results/Carers/Thematic synthesis<br>Tables 2. and 3.<br>Tables 4. and 5. |
| Risk of bias in studies | 18 | Present assessments of risk of bias for each included study. | Results/Search results<br>Table 1. |
| Results of individual studies | 19 | For all outcomes, present, for each study: (a) summary statistics for each group (where appropriate) and (b) an effect estimate and its precision (e.g. confidence/credible interval), ideally using structured tables or plots. | NA for Thematic Synthesis |
| Results of syntheses | 20a | For each synthesis, briefly summarise the characteristics and risk of bias among contributing studies. | Results/Search results<br>Table 1.<br>Tables 4. and 5. |
|  | 20b | Present results of all statistical syntheses conducted. If meta-analysis was done, present for each the summary estimate and its precision (e.g. confidence/credible interval) and measures of statistical heterogeneity. If comparing groups, describe | Results/Service users/Thematic synthesis<br>Results/Carers/Thematic synthesis |
|  |  | the direction of the effect. | Tables 4. and 5. |
|  | 20c | Present results of all investigations of possible causes of heterogeneity among study results. | NA for Thematic Synthesis |
|  | 20d | Present results of all sensitivity analyses conducted to assess the robustness of the synthesized results. | NA for Thematic Synthesis |
| Reporting biases | 21 | Present assessments of risk of bias due to missing results (arising from reporting biases) for each synthesis assessed. | Results/Search results<br>Table 1.<br>Tables 4. and 5. |
| Certainty of evidence | 22 | Present assessments of certainty (or confidence) in the body of evidence for each outcome assessed. | Results/Certainty of evidence<br>Tables 4. and 5. |
| **DISCUSSION** |  |  |  |
| Discussion | 23a | Provide a general interpretation of the results in the context of other evidence. | Discussion/Main findings |
|  | 23b | Discuss any limitations of the evidence included in the review. | Discussion/Strengths and limitations |
|  | 23c | Discuss any limitations of the review processes used. | Discussion/Strengths and limitations |
|  | 23d | Discuss implications of the results for practice, policy, and future research. | Discussion/Implications for policy and practice<br>Discussion/Implications for research |
| **OTHER INFORMATION** |  |  |  |
| Registration and protocol | 24a | Provide registration information for the review, including register name and registration number, or state that the review was not registered. | Abstract<br>Methods/Protocol and Registration |
|  | 24b | Indicate where the review protocol can be accessed, or state that a protocol was not prepared. | Methods/Protocol and Registration |
|  | 24c | Describe and explain any amendments to information provided at registration or in the protocol. | NA |
| Support | 25 | Describe sources of financial or non-financial support for the review, and the role of the funders or sponsors in the review. | Funding |
| Competing interests | 26 | Declare any competing interests of review authors. | Funding |
| Availability of data, code and other materials | 27 | Report which of the following are publicly available and where they can be found: template data collection forms; data extracted from included studies; data used for all analyses; analytic code; any other materials used in the review. | Availability of data<br>Supplementary information |

## Notes

### Competing Interest Statement

The authors have declared no competing interest.

### Clinical Protocols

https://www.crd.york.ac.uk/PROSPERO/display_record.php?RecordID=423439

