## Supplementary material for "A qualitative meta-synthesis of service users’ and carers’ experiences of assessment and involuntary hospital admissions under mental health legislations: a five-year update": Table 7

**Supplementary information, Table 7.** Selected additional evidence for Service User findings.

| **Themes and Sub-themes** | **References** | **Finding** | **Quotes – all participant names are pseudonyms.** | |
| --- | --- | --- | --- | --- |
| **1 Emotional impact** | | | | |
| **Acceptance** | 8  Bendelow et al., 2019; Lawrence et al., 2019; MacDonald et al., 2020; O’Connor et al., 2021; Smyth et al., 2021; Vallarino et al., 2019, Yu et al., 2022; Aluh et al., 2022 | The previous review highlighted occasions when participants appreciated or accepted the involuntary admission as necessary in some cases. Additional data from more recent papers similarly referred to positive aspects like feeling safe, addressing an underlying need and accessing support. Acceptance was nevertheless motivated on occasions by wanting to avoid conflict and the threat of coercion. | A small number of interviewees expressed a preference for being detained in police custody, as it gave them a feeling of safety, but most participants who were not taken to the hospital suite felt very strongly that ending up in a police cell when they had committed no crime was extremely stigmatising and distressing. Sinita, aged 32, described her gratitude for the s136 intervention which she said ‘saved my life’ at a time of great despair, but she stressed that if she had been taken to custody rather than the hospital suite, she would never have recovered from the stigma or been able to ‘move on’ with her life.  Bendelow et al., 2019  Another participant said she knew she needed the hospitalization. One suggested that the overall treatment experience was more important than voluntary or involuntary status.  INTERVIEWER: “Did you feel like just the aspect of being here involuntarily had an effect on how you related with your treatment team?”  PATIENT: “Not once I realized the way that the psychiatrist and the team were approaching. I work in public health in terms of the degree I received, so I understand how care should be delivered, so once I saw that that was happening, it immediately eased my tension, and I felt a lot better with being here. So, although I was committed involuntarily, it didn’t really feel involuntary.”  Lawrence et al., 2019  Overall, the participants’ opinions about IT included a general consensus that IT is occasionally necessary. They elaborated on this, stating that sometimes it can be difficult to reach and reason with a patient with AN, although it is not necessarily too late at the time of IT. It was described that staff should not be too hesitant using IT in order to avoid further escalation or death.  MacDonald et al., 2020 | |
| **Impact of detention** | 18  Aluh et al., 2022; Bendelow et al., 2019; Blakley et al., 2022; Goodall et al., 2019; Jaeger et al., 2019; Jones et al., 2021a; Jones et al., 2021b; Lawrence et al., 2019; McDonnaugh et al., 2020; McGuinness et al., 2018; O’Connor et al., 2021; Pothoff et al., 2022; Smyth et al., 2021; Sondhi et al., 2018; Vallarino et al., 2019, Verstegen et al., 2022; Wormdahl et al., 2021; Yu et al., 2022 | The previous synthesis identified anger, confusion, fear, distress, resentment and defensiveness as common emotional impacts detention, exacerbated by lack of information, police involvement, and behaviour of staff. The update uncovered similarly frequent, traumatic experiences, undergoing complex triage processes, away from one’s usual environment and dealing with unfamiliar people. Arriving to a place of safety was seen as the beginning of receiving support, but often a distressing, chaotic, high-risk environment. Detention in hospital settings was often described as negative, seen at times as punishment, barbaric, or a source of violent confrontations. Positive experiences with staff could at times ameliorate emotional impact, for example encountering a caring attitude, being listened to, or being trusted and offered control. | Moreover, information provided by place-of-safety staff was perceived to be more about fulﬁlling their obligations than informing their patient what was happening to them, compounding the negative experience of the detention process:  She [the nurse] didn’t come near me. She stayed at the door. I probably told her not to come any further ... I’d already been in September and I’d walked out, discharged myself. I never got any follow-up or offer of follow-up either anymore because .. . I felt threatened by the situation, so she was just part of that. I’ve no idea what she wrote down, except [that] I was sectioned. [Interview #52]  Sondhi et al., 2018  For some individuals the involuntary admission had a signifi-  cant impact on their well-being and relationship with others. Some described feeling traumatised by the process ‘When I went to the psychiatrist after 3 months… I said look, I want to go and talk to someone myself…to help you with the post-traumatic stress of being in the hospital in the first place.’ (participant 45, woman).  McGuiness et al., 2018  Despite the decrease in coercive practices during transportation (e.g., bodily restraint and use of handcuff s by the police) since the revision of the law, patients who had experienced trauma during hospitalization complained of constant anxiety and fear about hospitalization. Th ese experiences are irreversibly ingrained in their memories; the interviewees continued to be afraid of and hostile toward the idea of inpatient psychiatric treatment: ‘I had been forcibly committed by my family in the past. When my condition worsens, a feeling of extreme terror rises in me. They tied me up as soon as I got there. Since then, I have hated the process of getting there. I mean, would you like it? Being tied up by men, being dragged away. You do not know how much they assaulted me.’  Yu et al., 2022 | |
| **Impact of coercive treatment** | 8  Aluh et al., 2022; Goodall et al., 2019; Kalagi et al., 2018; MacDonald et al., 2020; McGuinness et al., 2018; Pothoff et al., 2022; Solanki et al., 2023; Yu et al., 2022 | In both the original and updated data coercive treatments were often described as an abusive, violating experience contributing to a sense of lack of control and a strong negative emotional impact (e.g. anxiety, fear, distress and dehumanisation). For example, seclusion triggered feelings of anger, loneliness and shame. Coercive practices in newly identified data included physical (including mechanical) and chemical, constant observation, or a locked environment. The use of mechanical restraint (referred to in a couple of studies) was linked with the experience of pain, humiliation and perception of assault. Witnessing other service users being restrained could be perceived as similarly fear provoking. Coercive practices were perceived unnecessary by some, justified only in limited instances (e.g. serious risk to life). serious risks to life. Some studies also referred to a vicious cycle in which coercion leads to increased anger and aggression, which in turn may be responded to with more coercion (e.g. restraint) by staff. | Anti‑therapeutic and traumatic Many participants felt that coercion was not an effective way of managing people with mental health problems, but rather aggravated the distress that they felt in times of crisis. It made them feel worse about the diagnosis, and receiving care from the same people who subjected them to various coercive practices was traumatic. “To me, is abnormal because it deals with mental health. Okay. Chaining, injections and all that, it affects psychologically. So, it’s not proper.” (FGD1,  male with MBDPS)  Aluh et al., 2022  Many experienced coercive interactions with professionals and described feeling frightened as they did not know where they were going: ‘… they [assisted admission team] just dragged me…They put me against the floor, used violence…they handcuffed me and they put me in this plastic yellowblanket and put me in a van or something…Ididn’t know where I was going.’ (participant 40, woman).  McGuiness et al., 2018  As well as access to outside space and the ability to smoke, participants speciﬁcally referred to negative experiences of excessive restraint and coercion as a part of detention, both medical and physical.  Solanki et al., 2023 | |
| **Feelings following discharge** | 8  Jones et al., 2021a; MacDonald et al., 2020; McDonnaugh et al., 2020; McGuinness et al., 2018; O’Connor et al., 2021; Smyth et al., 2021; Sondhi et al., 2018; Yu et al., 2022 | In the original review service users reported often feeling worse following admission than before, corroborated by new data. The negative impact of detention could be long lasting and lead to an increase in symptoms (e.g. depression, stress). Fear of readmission could affect relationships with community mental health services as well as family members. Some reported having positive experiences following discharge, for example having an increased motivation for change, focusing more on self-care, and seeing part of treatment as helpful to recovery. Returning to community life was seen as difficult at times, for example coping with less support, lacking adequate information on or access to services post-discharge. Some facilitators of successful discharge were emotional and practical support, engagement with community and social support, and a staged step-down process. | In total, 70% (30/40) of participants described negative  impacts on their ability or willingness to trust others— most frequently mental health providers, but in some cases extending to broader authority figures (parents, teachers) and peers. Both the intensity of distrust, where present, and degree of behavioral impact varied:  “…it made me not really want to open up to anybody because I would still go through the motions, but before, I would be a lot more open about it to people close to me and the psychologists. But afterwards, I didn’t really want to talk about things anymore”. “[Afterwards] I would always think about, I don’t want to go back to [the hospital]. I don’t want to … I can’t tell anyone I’m feeling this way because they’re just going to send me back.”  Jones et al., 2021  There was a wide consensus that community life was challenging. Participants described finding it hard to adapt to the lack of structure after being in the highly restrictive MSU setting and subsequently feeling overwhelmed by the freedom and responsibility:  P4.[...] going out doing shopping, making yourself something to eat [.. .][.. .] cos it was all sort of done for you. You know, like when you’re in hospital [.. .][.. .]I found it quite hard to start with[.. .].  P3. I’ve not had a case of independent living for quite some time, and I think it would be quite scary[.. .][.. .][.. .] Yeah, it’s more than scary it’s a nightmare (laughs)[.. .][.. .]ifI don’t get any sleep, or my anxiety and stress keys up I need someone to talk to[...]  McDonnaugh et al., 2020  A lasting imprint The participants experienced that IT had a negative imprint on them, often for a long time. Most of the participants described effects bordering on trauma symptoms, such as dreaming about IT, fear of being touched, trying to forget the experiences, or trying to avoid particular IT measures including mechanical restraint, nasogastric tube feeding, and involuntary admission. Persistent attempts to avoid physical restraint could occur after having witnessed it. Moreover, IT as contributing to low mood was described, although infrequently.  MacDonald et al., 2023 | |
| **Therapeutic benefit (NEW)** | 11  Aluh et al., 2022; Jaeger et al., 2019; Jones et al., 2021a; Kalagi et al., 2018; MacDonald et al., 2023; McGuiness et al., 2018; O’Connor et al., 2021; Smyth et al., 2021; Solanki et al., 2023; Vallarino et al., 2019; Yu et al., 2022 | In addition to the impact on emotions and self-worth, in recently identified studies service users’ expressed views on whether they saw the assessment and involuntary treatment as necessary and therapeutic, and whether it helped their recovery. Service users’ experiences were mixed in this domain. In some cases, admissions were described as providing little meaningful help in addressing distress or psychological help and in some cases lead to feeling worse psychologically. The nature of experiences and interactions with staff during admission (e.g. coercive, or inclusive) could affect trust and engagement with therapy in later stages. At the same time, in some instances therapeutic value was reported, for example aiding recovery, contributing to an increased care for oneself, or changing perspectives on managing life after admission. | “…that’s the thing, it makes you feel worse afterwards than you did before. I’m sitting here, I’m more depressed and stressed coming out of that, and freaked out, than I was going in before”  “It was so unhelpful. There wasn’t any kind of psychological help really to it. It wasn’t like therapeutic in any way.”  Jones et al., 2021  Anti‑therapeutic and traumatic Many participants felt that coercion was not an effective way of managing people with mental health problems, but rather aggravated the distress that they felt in times of crisis. It made them feel worse about the diagnosis, and receiving care from the same people who subjected them to various coercive practices was traumatic. “To me, is abnormal because it deals with mental health. Okay. Chaining, injections and all that, it affects psychologically. So, it’s not proper.” (FGD1,  male with MBDPS)  Aluh et al., 2022  All participants had access to individual therapy, which included mindfulness, relaxation and exposure for anxiety. Participants found psychoeducation, which was provided mainly via groups, to be useful: ‘I’ve done a stack of groups . . . It’s gone good. It’s opened my eyes to schizophrenia.’ (P4, lines 194–204).  O’Connor et al., 2021 | |
| **2 Impact on self-worth** | | | | |
| **Dehumanised** | 12  Aluh et al., 2022; Goodall et al., 2019; Jaeger et al., 2019; Jones et al., 2021a; MacDonald et al., 2020; Pothoff et al., 2022; Smyth et al., 2021; Wormdahl et al (2021); Yu et al (2022); Sondhi et al (2018); Verstegen et al (2022); Solanki (2023) | Previously, service users reported feeling dehumanised by coercive interventions, supported by newly identified data. Some described feeling like a caged animal during involuntary treatment, a sense of loss of identity, with everyday behaviours or traits being interpreted as a sign of illness or symptom by others. Staff showing genuine concern, treating people with dignity could have a positive effect on self-esteem. | | For most participants, the process of being admitted was a harrowing experience. They felt dehumanised and manhandled in the process of being brought to the hospital to be admitted involuntarily. Several participants reported being injured by the ropes and chains used to restrict movement while being transported to the hospitals. “.... My experience with them is that … they were supposed to counsel us, ... they don’t have conscience, they didn’t us treat us like human. They did not treat me like as if I’m a human being, despite my complain in the office there.”  Aluh et al., 2022  All participants associated the experience of detention as  being at odds with their sense of normal life and their basic human rights. Participants described detention as being something that should not be happening in the way that it does because it compromises freedom and justice. As a result, participants often compared detention to being treated inhumanely, like an animal in prison.  “I felt like I was being treated like an animal. I wasn’t allowed to go outside, I wasn’t allowed to have fresh air.” (Participant 10)  Solanki et al., 2023  “Sometimes, when there has been an incident of aggression on the ward, the sociotherapists are going to comfort each other in their office and we are just sitting there. And, well, I kind of do not like that, actually. As if we are less” [Patient 3].  This patient feels like a lesser human being because of the way the aftercare  is organized.  Verstegen et al., 2022 |
| **Power** | 12  Aluh et al., 2022; Blakley et al., 2022; Goodall et al., 2019; Jones et al., 2021a; Jones et al., 2021b; Lawrence et al., 2019; McDonnaugh et al., 2020; McGuinness et al., 2018; O’Connor et al., 2021; Pothoff et al., 2022; Vallarino et al., 2019; Verstegen et al., 2022 | Service users across a number of previously reviewed papers reported having a lack of control over the treatment process, e.g. having limited choices, lacking autonomy, not having an impact on timelines, or arbitrary ward routines. There were multiple references to power differences, paternalistic attitude by staff, needing permission, leading to feelings of dependency and reduced self-efficacy. Not being listened to, coercion (e.g. locked doors) could have a negative effect, whilst instances of collaborative care, being provided with choices, advocacy by others could have a positive effect on perceived control and autonomy during admission. | | ‘Helping people to feel more empowered’ as part of the pro-  cess was something identified by participants as a wish-list item; opportunities that could help to empower participants included reducing the use of handcuffs and restraint, and more access to advocacy services. The provision of an independent advocate for the individual was something deemed to be an important wishlist item by one individual.  Goodall et al., 2019  “The barrage of three”  Participants described being interviewed by three or more professionals as “daunting” (Charlotte, Alice), “intimidating” (Katie, Charlotte), “oppressive” (Stephen) and “a terrible pressure” (George). Katie talked about “the panel” expressing the idea of being judged with the number of assessors linked to a feeling of powerlessness: The barrage of three… looking at you, you know it is oppressive. (Stephen)  Blakley et al., 2022  Seven people reported a prior history of distressing compulsory admissions. Some were completely unaware of what was happening and reported feelings of failure, social isolation, disempowerment and shame. Two persons did not want to give any detail because they did not wish to relive the experience.  Vallarino et al., 2019 |
| **Stigma** | 14  Aluh et al., 2022; Bendelow et al., 2019; Goodall et al., 2019; Jaeger et al., 2019; Jones et al., 2021a; Jones et al., 2021b; McDonnaugh et al., 2020; McGuinness et al., 2018; Smyth et al., 2021; Solanki et al., 2023; Sondhi et al., 2018; Vallarino et al., 2019; Wormdahl et al., 2021; Yu et al., 2022 | There were frequent reports of service users feeling labelled, tainted or criminalised as a consequence of detention. For example, experiencing shame when neighbours witness them being handcuffed and taken by police, or losing credibility and being treated as dangerous for having a mental illness both within the hospital but also by society. There were reports of fear of being excluded and marginalised post-discharge and feeling at a greater risk of being detained again. Newer data corroborated experiences of violation and loss of rights, with frequent experiences of criminalisation, and not being believed. A few factors were seen as having a positive effect on stigma, for promoting open discussion on mental health, self-disclosure by professionals and public figures, and changing public and legal perceptions of mental health. | | Across these narratives, what we came to conceptualize as perceptions of moral judgement were common—for example the feeling of having been judged to be “lesser than,” “intellectually deficient” or “[like a] criminal” (see Table 2). Those participants describing these experiences often expressed anger about the fact that providers ostensibly tasked with supporting struggling youth instead blamed or belittled them.  Jones et al., 2021  At the same time, participants with lived experience and carers discussed how some individuals with SMI withdrew from services because they had experienced former admissions as traumatic. Among other things, they talked about being roughly handled, and often the police had been involved. When this happened in public, the participants experienced additional strain and stigma. Some said that the services were not tailored to help people overcome this fear around receiving services.  Wormdahl et al., 2021  Feeling able to talk openly about mental health problem without fear of judgement or shame with professionals and on occasions the use of self-disclosure of professionals for those who were detained to relate their experiences to. Discussing mental health in relation to public figures and celebrities also helped to challenge stigma.  Goodall et al., 2019 |
| **Positive impacts** | 8  Jones et al., 2021a; Jones et al., 2021b; Kalagi et al., 2018; MacDonald et al., 2020; O’Connor et al., 2021; Pothoff et al., 2022; Smyth et al., 2021; Solanki et al., 2023 | Service users also referenced activities or experiences during detention that built confidence, self-esteem, self-respect, or a sense of achievement. In newly identified data, admission was at times described as a turning point, contributing to becoming more independent, motivation to return to everyday life, or receiving more recognition of mental health from family members. Social proximity of others, working with skilled, recovery-oriented professionals, compassionate, genuine approach from staff, meaningful activities were some of the identified facilitators of these positive experiences. | | Beneficial here is without question the communication you can have here; that you’re not put offbut rather that you can directly interact with the nursing staff, […] in the locked setting it’s rather that they withdraw, and with this observation there is a kind ofcare, you get attention and warmth and feel social proximity, and that’s very conducive to your health. (Patient 2)  Kalagi et al., 2018  Other features that were mentioned were good care resources (in terms of staff, therapy options and equipment), relief from everyday obligations, and protection against relapse in relation to addiction, mental health crises, homelessness and loneliness.  Pothoff et al., 2022  The theme “progress” pertained to references about making and maintaining progress. Participants described progress strategies, which included: ensuring structured activities while on […]; engaging in meaningful activity; and psychology input on the ward: P5. (referring to a training position in the community) “It gave me a routine and a reason to get up, because you have to do it, which I needed because I was laying in a lot in bed, which is what you do in hospital - that inertia of getting up in the morning is really, really difficult once you come out of hospital.”  McDonnaugh et al., 2020 |
| **3 Information and involvement in care** | | | | |
| **Coercion, consent, choice** | 16  Aluh et al., 2022; Blakley et al., 2022; Goodall et al., 2019; Jaeger et al., 2019; Jones et al., 2021a; Jones et al., 2021b; Kalagi et al., 2018; Lawrence et al., 2019; MacDonald et al., 2020; McGuinness et al., 2018; O’Connor et al., 2021; Pothoff et al., 2022; Smyth et al., 2021; Solanki et al., 2023; Yu et al., 2022, Vallarino et al., 2019 | In both reviews, service users reflected positively on being provided flexibility in their care, which reduced the perception of coercion. Some reported experiences of collaborative care, but others described that their wishes and care preferences were ignored. Service users at times felt that coercive treatment, or the threat of involuntary admission undermined their ability to meaningfully consent to care. Based on newly identified data, experiences with coercion were predominantly negative, often distressing, potentially affecting future engagement with treatments offered. Intrusive observation, chemical or physical restraint, excessive force were seen as particularly negative, whilst being given choices where possible, caring staff attitudes, and alternative (e.g. recovery-oriented) practices were identified as potentially moderating harmful effects. | | With respect to their relatives, some interviewees reported how they felt supported, and others how they felt controlled and under pressure. Some believed that they were seeing some kind of coalition or conspiracy between their parents and the doctors, leading to mistrust and secrecy. Some patients assumed that their relatives’ behavior was motivated by their intention to help. Others suspected ambitions to dominate the patient, or lack of understanding in their family.  Jaeger et al., 2019  Patients said the involuntary experiences were difﬁcult because they felt as though they had no control over decisions, they did not feel like full participants in treatment planning, and they were not able to manage their lives outside the hospital (e.g., not able to make money). Three patients said it was difﬁcult to engage in treatment because they did not want to be there.  Lawrence et al., 2019  The first category ‘losing control’ refers to the extent of loss of autonomy individuals experienced as a result ofvarying levels ofmental distress and the extent individuals felt coerced during involuntary admission and subsequent hospital stay. Losing control arose as a result of internal and external pressures on an individual’scapacity to control their own life. ‘losing control’ comprised ofthree subcategories, ‘diminishing self-mastery’, ‘feeling violated’ and ‘being confined’.  McGuiness et al., 2018 |
| **Rights** | 6  Goodall et al., 2019; Lawrence et al., 2019; O’Connor et al., 2021; Smyth et al., 2021; Verstegen et al., 2022; Yu et al., 2022 | In studies reporting on legal hearings related to involuntary admissions some service users were pleased with steps facilitating their involvement. Examples of this included being given time to articulate thoughts, advocacy by staff or family, and legal representation. Others felt excluded by the presence of unfamiliar people and the formal language used. Tribunals were viewed favourably by patients as a method of upholding human rights, but difficulties in accessing relevant information or discussing this with staff were often reported. Newly identified studies corroborated that court experiences were at times negative, and service users at times had to navigate complex, lengthy, e.g. forensic legal processes. Being provided with adequate information, access to advocacy, and enhanced rights to self-determination were valued when offered. | | All participants acknowledged that legal processes dictated their transitions between different facilities and to community leave. Some participants expressed frustration with the amount of time spent in court to amend legal conditions (e.g., for overnight leave): ‘It was a lot of being put in a cell waiting while you’re at the courts waiting  to appear, only to have that adjourned . . . You’ve gotta be fairly patient.’ (P2, lines 440–445); ‘I’ve probably been to court close to twenty times for this offence.’ (P8,  line 850)  O’Connor et al., 2021  Third, the process of pressing charges was experienced as more difficult for  patients than for staff members and this raised concern among patients. Hospital staff not always promoted the patients’ rights to press charges. For instance, a patient was advised to shift his attention from the wrongdoing of others to his own treatment and thereby, not press charges. Furthermore, relations between patients become more complicated after pressing charges and patients are condemned to each other. For instance, patient 1 was warned that he could report the situation to the police, but then the other patient will, most likely, also press charges against him and due to national regulations, the liberty of the patient to leave the hospital will be suspended for one year if charges are brought against someone. This means that the patient would risk losing his liberties if he pressed charges against the patient that harmed him.  Verstegen et al., 2022  ‘Helping people to feel more empowered’ as part of the pro-  cess was something identified by participants as a wish-list item; opportunities that could help to empower participants included reducing the use of handcuffs and restraint, and more access to advocacy services. The provision of an independent advocate for the individual was something deemed to be an important wishlist item by one individual:  “I would ask for a mental health care worker. But when you’re like that, you don’t think of anything. You see when you’re vulnerable like that, you need somebody of authority on that, on the situation…”  Goodall et al., 2019 |
| **Information about what’s happening** | 11  Aluh et al., 2022; Blakley et al., 2022; Goodall et al., 2019; Jaeger et al., 2019; Jones et al., 2021b; Lawrence et al., 2019; McDonnaugh et al., 2020; Pothoff et al., 2022; Smyth et al., 2021; Sondhi et al., 2018; Vallarino et al., 2019 | Patients described wanting information about the reason and length of their admission, and about legal rights. Those in forensic settings described receiving conflicting information about their length of stay resulting in feelings of hopelessness, corroborated by data from newer studies. In many studies, patients reported that they were not given basic information about medication or perceived progress. Provision of clear, relevant information was less frequently mentioned, but in these cases appeared to empower service users, reduce fear, and improve relationships with staff. Newly identified studies highlighted several steps in the admission process where information to service users could be lacking: formal assessments for admission, accessing place of safety, taking medication, or discharge. At the same time, being provided with too much information at the wrong time (e.g. when distressed) could potentially be overwhelming. | | Deceived and kept in the dark Many participants narrated how they were tricked into going to the psychiatric hopitals and then admitted against their will. They complained about being taken unawares and, in some cases, it seemed to them like they were being kidnapped. “You cannot just come into deceiving, you know. In my own case I was deceived. I was, they told me I’m going for my business work, but on seeing this sign board, I now notice that they have lied to me. So that’s what I’m saying that the whole system is illegal. Because if they had told we are going to psychiatric hospital for evaluation, I will know and that’s what I thought. They told me that I’m coming for work that they will pay me after my work, with that in mind, I started coming.” (FGD4, female participant with schizophrenia )  Aluh et al., 2022  Information and options  No participant completely understood the MHAA process, even those who had had multiple experiences of assessment. Participants explained in numerous differing ways how they lacked information and options. Some participants did not recognise the process as a MHAA: I did not know why they were there (Thomas) It’s like it’s deliberately secretive…They don’t say “we are a group of 3 people one of us is going to be the main person to ask questions and what we are doing is analysing you to see…if you need to be detained under the mental health act”. Why don’t they say that? (Stephen)  Blakley et al., 2022  ‘I was given plenty of information . . . they [nurses] gave me the mental health booklet. Somebody talked to me about the tribunal. . . they always made sure that I was aware of what was coming up next’ (Female 214 High).  Smyth et al., 2021 |
| **Involvement in treatment decisions** | 12  Aluh et al., 2022; Bendelow et al., 2019; Blakley et al., 2022; Jaeger et al., 2019; Jones et al., 2021a; Jones et al., 2021b; Lawrence et al., 2019; O’Connor et al., 2021; Pothoff et al., 2022; Smyth et al., 2021; Solanki et al., 2023; Yu et al., 2022 | The original review and new data described similar service users in this domain. In many studies patients described wanting to be involved in decisions about their care; very often more than was offered. Newly identified studies contained frequent reports of similar experiences: lacking control, not being listened to nor offered options during assessment or treatment. Good relationships with staff, being part of the planning process, discussing options with friends and family facilitated involvement in decision-making. Flexibility in care, involvement in creating treatment plans also reduced the perception of coercion. Raising feedback was also reported as difficult in several new studies, where patients’ concerns were not followed up satisfactorily by staff. | | One option that some participants felt was not fully explored was agreeing to admission (informal) with one the majority of participants talked about not feeling involved and the outcome being a fait accompli: There’s no point saying anything when it’s not going to have much difference (Natasha)  Decision to be hospitalised was inevitable whether I liked it or not. (Bridget).  I wasn’t involved… I got the feeling made mind up before coming through the door. I was going to hospital. They weren’t going to listen to me (George) I was the subject of it but it didn’t feel like a two-way process (Stephen)  Blakley et al., 2022  They didn’t even allow me in my ward round. How would you feel about that? You’d be unhappy right? :: : Who wants their destiny to be decided without them there? (Participant 8)  Solanki et al., 2023  I don’t want people who have got tired of the job and tired of everything that’s happening and sort of, have given up. Because .. . that’s not very motivating for us as patients. I would like a proper care plan. I’ve never seen my care plan in 13 years. I’d like a proper care plan. I’d like to be part of what that care plan was. (Suzanne, aged 41)  Bendelow et al., 2019 |
| **Medication** | 10  Aluh et al., 2022; Jaeger et al., 2019; Jones et al., 2021a; Lawrence et al., 2019; O’Connor et al., 2021; Pothoff et al., 2022; Smyth et al., 2021; Solanki et al., 2023; Vallarino et al., 2019, Yu et al., 2022 | In papers previously reviewed, forced medication was a source of particular distress. Some patients a lack of opportunity to make a fully informed decision, being offered what they perceived to be a false choice and threatened with punishment. Side-effects of medication were at times difficult to tolerate and could restrict participation in other therapeutic activities. In contrast, others felt medication could reduce some symptoms and contribute to recovery. In both previous and more recent studies, treatment during detention was described as predominantly pharmacological, despite the demand for psychological therapies. Rejection of medication was also frequently discussed: turning down prescribed treatment could lead to more coercion, longer inpatient stays, or family disagreements. Levels of continuation after the end of involuntary treatment was reported as varied. | | Experience of Chemical Restraint Undesirable effects Many participants (43.3%, n = 13) complained that the chemical restraints had negative effects on them, from sleeping for prolonged periods, inability to walk because they were injected into their thighs, and dyskinesia. Many participants described the process of administering the chemical restraint as humiliating because their clothes were forcibly removed to administer the injections.  “E get some kind of injection, you give some patients here, the body will change automatic, some will start turning their eye, some will start shaking their body and I want the hospital look into... it.” (FGD3, male participant with MBDPS)  “I was injected with sleeping injection, so I slept off that night. When I woke up in the morning, they’ve injected my both laps. I can’t even walk for like a week and some days.” (FGD3, male participant with MBDPS) “So, I was held down, pinned down and my... covering was physically and manually pulled down off my waist and the syringe was inserted into my body and alongside I had some bruises on my fist. I think it’s a  very bad experience.” (FGD2, male participant with  MBDPS) “...and when they injected me, I slept immediately and those that were with me said that is for two consecutive days that I didn’t wake up. So that’s the effect of the drugs.” (FGD4, male participant with schizophrenia)  An attitude which did not differ by insight scores was the perception of the excessive centrality of pharmacotherapy. A large number of participants were critical of the focus on medical interventions and perceived an insufficient provision of psychotherapeutic options. ‘I just felt that I was having medication thrown at me . . . there was a lot of psychological aspects to it that aren’t really addressed. Everyone seemed to be focusing on drugs as a solution’ (Male 28 High).  Smyth et al., 2021  Interviewees in all four groups similarly reported negative  consequences of medication refusal or discontinuation. Positive consequences were rarely reported. A few patients claimed to feel more energetic and alive without medication. But even these patients told of negative consequences.  Jaeger et al., 2019 |
| **4 Quality of relationships** | | | | |
| **Police and emergency department staff** | 8  Bendelow et al., 2019; Goodall et al., 2019; Jones et al., 2021b; Lawrence et al., 2019; McGuinness et al., 2018; Solanki et al., 2023; Sondhi et al., 2018; Wormdahl et al., 2021 | Initial contact and experience on service entry was described as varied in the previous review. People at times experienced kind and gentle treatment, but at times staff were felt to be dismissive or lacking training in mental health. Newly analysis studies reported a similarly mixed experience. Forceful treatment, inadequate responses, rejection, poor communication with service users and/or between professionals have been particularly unhelpful. Instances of the opposite: examples of caring, kind, emotionally supportive treatment have also been highlighted. Police were at times seen as helpful and taking distress seriously, but several papers reported experiencing the involvement of this service as stigmatising, with staff on occasions dismissive or using excessive force. | | ‘Staff who use excessive physical restraint or force’, such as by  clinicians during admission, or the use of handcuffs by police during the initial phases of detention, was reported as an unhelpful CI by a small number of participants. The use of handcuffs on one participant by police upon arriving at a public place was something that was held in memory by that individual: “Yeah, but that was with… They put handcuffs on to do that, then they did that after onto my shoulder. And I was screaming out in pain. Researcher: So when you think about being put in handcuffs, what’s your thoughts about that? Well it’s…its not… If you’re put in handcuffs, you’re put in handcuffs there. You’re not put in handcuffs behind your back. Why did they put me handcuffs behind my back?”  Goodall et al., 2019  Moreover, information provided by place-of-safety staff was perceived to be more about fulﬁlling their obligations than informing their patient what was happening to them, compounding the negative experience of the detention process:  She [the nurse] didn’t come near me. She stayed at the door. I probably told her not to come any further ... I’d already been in September and I’d walked out, discharged myself. I never got any follow-up or offer of follow-up either anymore because .. . I felt threatened by the situation, so she was just part of that. I’ve no idea what she wrote down, except [that] I was sectioned. [Interview #52]  Sondhi et al., 2018  Subcategory 2.2: ‘encountering humanising care’ This describes how participants began to regain control through positive and supportive interactions with people who were in control (for example police and clinicians) and who were willing to take a  risk and giving participants some agency and control, despite their legal status. For example, being provided with choice and options such as being allowing to decide on the means of transport to hospital was viewed by participants as an opportunity to regain some control over their situation. ‘I had my car with me…He [Police man]…had me follow him, so he was actually trusting me…the independence…the trust …that was important (participant 38, man)’;  McGuiness et al., 2018 |
| **Inpatient staff** | 13  Aluh et al., 2022; Bendelow et al., 2019; Blakley et al., 2022; Goodall et al., 2019; Jaeger et al., 2019; Jones et al., 2021a; Kalagi et al., 2018; Lawrence et al., 2019; McGuinness et al., 2018; Pothoff et al., 2022; Smyth et al., 2021; Solanki et al., 2023; Verstegen et al., 2022 | Experiences of relationships with inpatient staff were similarly varied. Service users in both reviews highlighted staff qualities that contributed to building positive relationships: making time to talk to patients, building a connection, being approachable talking openly about mental health, and providing emotional support. Some staff were described as disrespectful, bullying, or unavailable which was seen as detrimental to the therapeutic relationship and leading to feelings of anger and distrust. Newly identified studies highlighted the continued experience of bullying or abusive behaviour, infliction of fear of physical pain, lack of transparency in communication, not being listened to in relation to some staff. The involuntary nature of admission, high turnover and tired, overworked staff with little resources could also lead to tension and negatively affect relationships. | | Helpful critical incidents  Being detained under S136 whilst experiencing significant emotional distress is not likely to be a positive experience, however, participants were able to identify a number of factors which eased, or helped with the experience of being detained. These are outlined in Table 2. One of the most prominently reported categories was ‘staff  who provide emotional support’. This appeared to be a common experience for many participants and was the most frequently identified helpful critical incident. This involved experiences where staff emotionally supported those who were detained through providing verbal reassurance, validating emotions and enquiring about how the person was coping with the experience. One participant described the way in which staff at the place of safety treated them upon their arrival: “No, just being kind. Researcher: Okay. How were they kind? They was just kind to me. They spoke gentle to me, told me what was going on, and just telling me that they were there for me.”  Goodall et al., 2019  What do mental people who are going through a very hard time need? Kindness . . . A touch of reassurance . . . I would have preferred a cancer diagnosis than that because my fear of all of that . . . There was no hint of ‘look, we really want to help you’ . . . (Female 217 Low)  Smyth et al.., 2021  The quality of the personal relationship The data suggested that perceived psychological pressure depends on whether personal relationships with professionals or informal caregivers are experienced as supportive or discouraging. Discouraging interpersonal relationships were characterized by service users as involving a lack of transparency, a lack of emotional  support, a feeling of being unknown to each other, unfair treatment and strong dependence. Communicative interactions in the context of discouraging relationships were more likely to be perceived as involving psychological pressure. One participant  reported discouraging per-  sonal relationships with professionals in the context of an admission process:  When I look back on it in retrospect, one could have said, “Mr. X1, there is something wrong with you, you are in a manic episode, you are out of line, you are … not in command of your powers, … or you are not sane at the moment … we have to keep you here and if you don’t want that, then unfortunately we have to restrain you.” Something like that, a clarifying conversation or something. That somehow didn’t happen at all. But it was all just like this, “Here, Mr. X1, now stay here and sit down and … let’s do something like this.” … Run-of-the-mill exchanges, according to the motto ‘The main thing is that we calm him down.’ (Service user 1).  Potthoff et al., 2022 |
| **Family and friends** | 11  Aluh et al., 2022; Bendelow et al., 2019; Blakley et al., 2022; Jaeger et al., 2019; Jones et al., 2021a; McDonnaugh et al., 2020; McGuinness et al., 2018; O’Connor et al., 2021; Pothoff et al., 2022; Smyth et al., 2021; Solanki et al., 2023 | Both the previous and new data revealed similar aspects of the relationship with family and friends. Their role was seen positively by many service users, due to the emotional support, discussions over help-seeking, help with speaking up, providing a reminder of own identity, and continued role following discharge. At the same time, due to the role in involuntary admission these relationships could be accompanied by feelings of distrust and abandonment. Newer studies reported on admissions could at times lead to closer relationships, but also concerns over service users fearing being misinterpreted by family members, thus deciding to disguise their true feelings, and on occasions distancing themselves. | | There was a call for more support within this process  from family or others and Katie feeling the process would then feel fairer and help to give a voice:  If my mum was in the room, I know that I’ve got somebody who knows me and who’s not going to let anything happen that shouldn’t happen. So you don’t get… doctors …talk over you or anything.  Blakley et al., 2022  All ten of the intact trust participants described at least one significant “indirect” positive impact of hospitalization (compared to a minority of the distrust sub-group). Examples included participants’ parents or families taking their mental health challenges more seriously after discharge; experiencing less blame from family or friends; or ultimately receiving mental health services (including medications) as a result of their hospitalization that they felt were helpful.  Jones et al., 2021  Subcategory 3.2: ‘preserving sense of self’ This describes the strategies used to minimise stigma, and deal and contend with other’s perception of them. Individuals were mindful not to state or engage in anything that could be construed as a reason for readmission, opting to conceal certain thoughts or deliberately try to behave in a socially acceptable manner: ‘I’m really afraid to say anything to my husband… I don’t give out about people… I think I couldn’t start saying any of those things I was saying before that led me to be brought in …’ (participant 44, woman). ‘I have to mind by Ps and Qs because my husband… he’d probably sign me in again’ (participant 39, woman).  McGuiness et al., 2018 |
| **Other patients** | 8  Jones et al., 2021a; Lawrence et al., 2019; McDonnaugh et al., 2020; O’Connor et al., 2021; Pothoff et al., 2022; Solanki et al., 2023; Vallarino et al., 2019; Verstegen et al., 2022 | Previously reviewed papers described people gaining encouragement and support from contact with peers, for example when witnessing recovery, but also tension on occasions with other service users, partly due to staying on overcrowded wards. Newly identified data corroborated these experiences, with peer support and experience valued in managing one’s own well-being. Specifically negative aspects resulting in fear, avoidance or mistrust were encountering conflict (e.g. verbal, physical aggression or sexual transgressions), or witnessing others being subjected to coercion and forceful treatment. | | Participants described the value of sharing experiences and forming peer friendships. For example, P7 said, ‘I’ve made some friends while I’ve been here . . . that’s something positive I can take away with me.’ (P7, lines 319–321). Social contact was facilitated through community meals, group leave and treatment groups. The community meal, for example, was identified as a safe environment where residents could discuss concerns and individual achievements.  O’Connor et al., 2021  You can be around other people who are going through the same stuff as you, so you don’t feel :: : awkward about saying, “Oh I felt like killing myself” or “I had these thoughts yesterday.” So you got people who’s going through the same experience. (Participant 6)  Solanki et al., 2023  Fear was a common response to aggression. Patients, for instance, reported that they feared that previously violent patients will reengage in violent behavior, that tense situations between patients escalate into violence or that patients would use equipment that is available in the hospital as a weapon. In all these situations, patients feared that they, or staff members or other patients that they care about, would get hurt. Patient 3 described how another patient threatened to assault her in an empty room on the ward. Afterward, she was very afraid to be alone.  Verstegen et al., 2022 |
| **Playing ball (NEW)** | 6  Jones et al., 2021a; McDonnaugh et al., 2020; McGuinness et al., 2018; Pothoff et al., 2022; Smyth et al., 2021; Verstegen et al., 2022 | Both previous and new studies addressed the negative emotional impact that the assessment and admission process may inflict on service users. Some patients referred to developing various strategies in order to cope with aspects of their care and treatment environment. This included increased self-regulation, changing the way people communicate or handle potential conflict with others, disclosing their symptoms and mental health more cautiously to professionals to avoid different, or longer inpatient treatment. | | “The mindset you get into there, at least what I got into was like, ‘Okay, I need to pretend I’m okay so that they’ll let me out.’ Because you aren’t going to get better in that situation. You’re just gonna pretend to be better, so they’ll let you out, so you can go back to an easier life.”  “[One of the other patients said] ‘Hey, you need to stop crying,’ and I was like, ‘Why? I don’t care. Why?’ And one of them is like, ‘Well, they won’t let you out unless you show emotional stability,’ and I was like, ‘Oh my God. Okay.’  Jones et al., 2021  Participants talked of how they used self-monitoring and self-correction in order not to arouse doubt over their readiness for discharge. Participants used different strategies to achieve this, including: masking negative emotions, carefully managing interactions with both staff and patients on the ward, complying with all requests from staff, and isolating themselves to avoid trouble:  P1. Sometimes you just wanna put your head in your hands and just say “when am I getting out of here!?”[.. .]. and you can’t really do that in front of other people .. . it gets reported saying, saying, oh “X was a bit depressed or something”; but it never did get reported back cos I was good at putting on a front.  McDonnaugh et al., 2020  Subcategory 2.4: ‘playing ball’ This refers to other strategies adopted by some individuals to limit the extent of coercion exerted upon them. Such individuals saw no benefit from being involuntarily admitted and deliberately monitored what they said to professionals, did not disagree or ask questions of professionals. They conformed to the system by reluctantly agreeing to comply with professionals and treatment, as one participant commented ‘you learn keep your head down, say nothing’ (participant 39, woman).  McGuiness et al., 2018 |
| **5 Quality of environment** | | | | |
| **Police cells** | 4  Bendelow et al., 2019; Goodall et al., 2019; Pothoff et al., 2022; Sondhi et al., 2018 | The material environment in these facilities were often found to be cold, noisy and distressing, where lack of treatment could contribute to worsening of symptoms. Newer reports also highlighted that, whilst at times this was experienced as a place of safety, being taken to police custody was often associated with a prison-like environment, and feelings of shame and stigma. | | The absence of meaningful information provided throughout the process from detention to arrival at a place of safety was a major concern for many interviewed:  It’s just one big black hole that assessment room they keep you in, nothing, no information as to what is going to happen, by when and who is doing it. Dump you in the room to be stared at like some sort of strange animal. [Interview #88]  Sondhi et al., 2018  Through being provided with tangible or practical support  such as the provision of food or drink, several participants reported that this helped to ‘make the experience more tolerable’ when detained under S136. One participant recalled how she was advised by a healthcare support worker about the availability of food and drink during her detention at the place of safety: “She even went out of her way, and she goes well sorry erm… lunch has gone, teatime has gone and all the rest of it, but I will see what I can do for you. And she came back erm, like 15 minutes later. She brought me tea, erm sorry a coffee, sandwiches and biscuits and erm… she was so helpful. So helpful.”  Goodall et al., 2018  The material surroundings The analysis of the data indicated that the material surroundings of communication also influence perceived psychological pressure. The following quote shows exemplarily how the material surroundings of communication shape the meaning of communication, especially by influencing service users’ inferences about the consequences of noncooperation:  Then I came into a room … was supposed to sit at the table there. First I got something to eat. It was around noon and that probably wasn’t a normal patient room, but it was, I don’t know if you call it an admission room, where there is actually no office, but a room, but with windows so that you can look inside. Where there was a table with chairs, but also a patient bench, and these belts were attached to this patient bench and they are used if necessary … but that was in there by default, and that irritated  Potthoff et al. BMC Psychiatry (2022) 22:186 Page 10 of 13  me because I thought, why is this patient bench there now or why is it there with these devices? (Service user 10)  Potthoff et al., 2022 |
| **Hospital wards** | 8  Jones et al., 2021a; Kalagi et al., 2018; Lawrence et al., 2019; McGuinness et al., 2018; Pothoff et al., 2022; Smyth et al., 2021; Solanki et al., 2023; Vallarino et al., 2019 | Both old and new reports described the physical environment as important for recovery, but at times not meeting expectations from a therapeutic environment. Rigid routines, punitive methods, locked doors contributed to a prison-like feel to many service users. In newer studies, there were accounts of service users who valued having a tranquil, well-equipped safe space, increased freedoms, and access to therapeutic activities whilst on hospital wards. Seclusion rooms were seen as bare, cold, uncomfortable, or in forensic settings akin to a prison cell. | | Unmet expectations “My only experience with inpatient stuff was from movies, which was not accurate. I expected it to be more like [‘To the Bone’ on Netflix]. You were just kind of at a hotel, and you were allowed to do your own thing and…in the movie they had their phones. So I expected, I was like I’ll be able to have my computer and all this stuff. It shouldn’t be too bad. I won’t get behind in school. [But] It was not like the movie at all, obviously.”  “I didn’t know what involuntary inpatient even was. What I thought something like that would be, would be more like a lot of counseling, a lot more help in that sense. But, it was less of help and more of like, ‘Oh, we’re just going to keep you for 72 [hours] …’”  Jones et al., 2021  Our data indicated that psychological pressure is used to increase adherence to not only treatment but also social norms. The latter type of psychological pressure was reported predominantly in relation to house rules concerning mealtimes, television times and wake up times. Other rules in relation to which service users typically experienced psychological pressure concerned smoking or drinking coffee or tea between meals.  Potthoff et al., 2022  I felt like I was being treated like an animal. I wasn’t allowed to go outside, I wasn’t allowed to have fresh air. (Participant 10)  I feel like I’m more in prison than I am in a mental institution. I do, it feels like a prison :: : There shouldn’t be restrictions on smoking, not in a mental institution or prison ‘cause that’s the only thing they’ve got. (Participant 6)  Solanki et al., 2023 |
| **Forensic wards** | 2  O’Connor et al., 2021; Verstegen et al., 2022. | In the previous review service users reflected on strict security measures reminiscent of prison. In newer studies some of the risk management processes were seen as intensive but sometimes acceptable. Service users valued forensic services that were designed in a step-down fashion, embracing recovery-oriented approaches as these were seen as aiding people’s progress and preparation for life following admission. | | Participants contrasted the FSDRU with previous settings, in that there was  more privacy and greater freedom of movement. They identified physical security measures as less restrictive: “I’ve been in the system for years, so this is a good place to end up . . . It’s a good stepping stone before I get back in the community . . . A lot of people class (the  high security forensic unit) as jail, ‘cause you’re locked up. Here, you haven’t got that. You’re not locked up as such.” (P1, lines 71-77).  O’Connor et al., 2021  Participants’ responses concerned the requirements outlined in the relevant legislation concerning their detention and supervision, including requirements for periodic assessment by psychiatrists. For some participants, the assessment process was a difficult experience: ‘A lot of raw topics come up, involving your offence . . . It’s very, very confronting . . . It brings things back up again.’ (P1, lines 109–111).  O’Connor et al., 2021  As patients were confronted with their aggressors and repeatedly had to talk  about the incidents, these aggressive incidents continued to influence the atmosphere on the ward, even if the acute situation was resolved. This means that patients were confronted with the consequences for a long time and were not able to escape it  Verstegen et al., 2022 |
| **Meaningful activities** | 4  Jones et al., 2021a; Kalagi et al., 2018; McDonnaugh et al., 2020; Wormdahl et al., 2021 | The importance of recreational, education, occupational activities in helping maintain routine and progress, and lowering tension has been highlighted in both review stages. At times access to these were affected by fears for personal safety, or low staffing levels. One study highlighted that having a diverse range of meaningful activities is needed to successfully match the needs of different groups. | | Patients and nurses critically note that having too little time to engage in activities or conversations with patients gives patients the feeling that they are not being taken seriously. As a result, tension builds up.  Kalagi et al., 2018  While only a few ‘trust intact’ participants described their experiences as positive overall, many more noted positive aspects of hospitalization that appeared to soften or counterbalance negative experiences. Examples included beneficial therapy groups or activities, “stand out” staff who left them feeling cared about and heard, and roommates or other youth with whom they bonded.  Jones et al., 2021  I think a more diverse oﬀer of activities to those who need it would be good, because there is not much to choose from now, especially for men. We have a day center but they oﬀer mostly knitting, crocheting and reading the newspaper and stuﬀlike that. They should organise things like data, golf, bowling and outdoor activities. It is time for some innovation. It is important to have good arenas to meet, generally in the community, in the city, or where you live, but the municipality here has no other activities to oﬀer outside the day center. (Individual with lived experience)  Wormdahl et al., 2021 |
| **Personal safety and security** | 6  Bendelow et al., 2019; Jones et al., 2021a; Kalagi et al., 2018; Pothoff et al., 2022; Verstegen et al., 2022; Yu et al., 2022 | This domain has been previously identified as key to service users evaluating the quality of their environment. Whilst places of safety and hospital wards were seen as helping averting risk and protection from harm, there were many accounts of fear for personal safety in both sets of studies. Commonly reported risks were aggression, verbal or physical assault, sexual transgression, or threat to property. Newly identified studies contained reports of fear, hypervigilance, and withdrawal as a response to fear of violence. Friendly, reassuring presence of staff were seen as promoting feeling of safety. | | A small number of interviewees expressed a preference for being detained in police custody, as it gave them a feeling of safety, but most participants who were not taken to the hospital suite felt very strongly that ending up in a police cell when they had committed no crime was extremely stigmatising and distressing. Sinita, aged 32, described her gratitude for the s136 intervention which she said ‘saved my life’ at a time of great despair, but she stressed that if she had been taken to custody rather than the hospital suite, she would never have recovered from the stigma or been able to ‘move on’ with her life.  Bendelow et al., 2019  almost half (14/30) reported feeling ‘terrified,’ ‘scared’ or ‘unsafe’, while confined  Turning to narratives of more specific incidents of harm, one participant reported a sexual assault and two others sexual harassment by other inpatients; in all three cases, participants felt that staff took only minimal steps to ensure their safety and superficial de-briefing.  Jones et al., 2021  Fear was a common response to aggression. Patients, for instance, reported that they feared that previously violent patients will reengage in violent behavior, that tense situations between patients escalate into violence or that patients would use equipment that is available in the hospital as a weapon. In all these situations, patients feared that they, or staff members or other patients that they care about, would get hurt. Patient 3 described how another patient threatened to assault her in an empty room on the ward. Afterward, she was very afraid to be alone.  Verstegen et al., 2022 |
| **6 Discrimination** | | | | |
| **Racial discrimination** | 3  Jones et al., 2021b; Solanki et al., 2023; Verstegen et al., 2022 | Whilst only a few studies explicitly addressed this topic, experiences of discrimination based on race and ethnicity have been reported from three separate contexts. These included a paper on young adults’ views on police involvement in detention for psychiatric assessments, one on the experiences of service users of a Black Ethnic background admitted under the MHA in the UK, and one taking place in a Dutch forensic setting. In terms of police involvement in US, Florida, some people felt that police conduct was disrespectful in general, whilst some others felt that they were treated differently specifically because of their race (Jones et al., 2021b). In the UK inpatient context service users reported experiencing abuse and discrimination because of their race, both during their treatment and in society in general (Solanki et al. 2023). In the same paper, a service user also described being perceived as stronger, and subjected to harsher treatment by staff due to being a Black man. In the Dutch forensic setting, ethnicity was described as one of several characteristics that made it more likely that a service user would be targeted by peers in a confrontative, violent manner (Verstegen et al., 2022). | | When directly asked about the potential role of racial-ethnic discrimination in their handling by police ofﬁcers, most participants of color expressly conveyed that they felt that race-ethnicity had not directly inﬂuenced their treatment, although these statements were often paired with broadly cynical views of the police (e.g., “[Race-ethnicity] . . . not really. I just don’t really think the cops really treat anybody with respect. They always, to me, always treat you like a criminal ﬁrst no matter who you are.” [participant 14, Latino male high school student]). Meanwhile, others felt or at least suspected that race ethnicity had a role in their treatment:  Yes. I feel because I’m a Black Hispanic and I’m a minority that they just maybe took it differently, maybe judged me in that character that . . . because you know if you see that . . . I don’t know if it just goes with the police too . . . just because I look like that, because of my race, that’s why [what] happened [happened]. (participant 18, Black Hispanic female high school student)  Jones et al., 2022  I Am Not a Person—I Am a Black Patient The second theme is a quote from one participant and brings  together reports of how various aspects of a participants’ experience of detention were linked to their ethnicity. This was described as an additional component to the experience of being detained, with participants reporting abuse and discrimination associated with their ethnicity. Participants did not always specify who they experienced this from, however, patients, staff, and society as a whole were identiﬁed or mentioned in the same passages.  There’s a few foes, few racists [unspeciﬁed] in there calling me “nigger,”“monkey” and whatever, but, I get that every day anyway so it don’t really bother me anymore. (Participant 6)  I mean we all know, there’s no point kidding ourselves, this is generally a racist country :: : from my experience of ﬁfteen years of having a mental illness. Being black, you are treated as if you’re superhuman, you’ve got superhuman powers ::: you just get treated differently because you’re black. They [staff] assume because you’re black that you’re stronger :: : you can take it. (Participant 2)  That’s a racial thing. The less, the least of us on the street, the better. Especially the men. That’s what I think. I think they want it. I think they get us in, drug us up, some of us never come back :: : That’s what I think, it’s racial. (Participant 6)  Solanki et al., 2023  According to the interviewed patients, not all patient groups have equal chances of being targeted. Patients who have committed a sexually violent offense toward children, patients from an ethnic minority groups, homosexual patients and female patients were pointed as groups that were more often subjected to inpatient aggression.  Verstegen et al., 2022 |
| **Inequality of access** | 4  Bendelow et al., 2019, Goodall et al., 2019; Sondhi et al., 2018, Yu et al., 2022 | Several newly identified studies contained accounts of service users receiving insufficient treatment or being unable to access care in a timely manner due to their age, gender, demographic or personal characteristics, or medical history. Examples included delayed service entry due to having an additional addiction diagnosis (Bendelow et al., 2019; Sondhi et al., 2018), young people’s mental health services not well placed to support young women with experiences of past trauma/abuse (Bendelow et al., 2019), or wishing to see more diverse staff to facilitate better communication, e.g. people of all genders in the police services (Goodall et al., 2019). | | Younger women were very critical of support from Child and Adolescent Mental Health Services, especially with regard to sexual abuse, and nearly all the women with this history felt that statutory adult mental health services were unable to offer the help they needed to manage their dissociative episodes or address the traumas underpinning their mentalhealth problems: ‘[My community mental-health team] are underresourced, and in my most recent meeting with them, I was told that if I’m in crisis, the only option is to call the police!’ (Nina, aged 18). A similar theme emerged from our qualitative data. Several interviewees who had not been previously known to services described self medicating with alcohol to cope with their distress. Other participants who had a known addiction were referred back to substance misuse services when they were in crisis, in some cases with a three-month wait for an appointment. These long waits for mental-health assessments or substance-misuse referrals had escalated the impending crisis in some cases.  Bendelow et al. (2019) UK, Sussex  UK, views on the S136 process (place of safety, police custody) by people experiencing distress Given the relative high usage of alcohol and illicit drugs prior to their mental-health episode, many participants also expressed frustration at being denied access to a place of safety to be ‘medically cleared’ at an emergency department, which often required additional travel across hospital sites.  Sondhi et al. (2018)  South Korea, involuntary admission and discharge experiences following legal change Most older adult patients who had been hospitalized for long periods ended up moving from one mental health care facility to another because they did not have families to go home to, or were unable to use residential facilities due to age restrictions: If you reduce the number of hospital beds to achieve deinstitutionalization and forcibly discharge patients, where are the elderly patients who had remained at hospitals supposed to go? Their families will not take them in. Th ere are no facilities either, are there? I once heard that the patients end up going to a nursing home for the homeless in the rural areas.  Yu et al. (2022) |
| **7 Pathway to admission** | | | | |
| **Pathway to admission** | 7  Bendelow et al., 2019; McGuiness et al., 2018; Smyth et al., 2021; Solanki et al., 2023; Vallarino et al., 2019; Wormdahl et al., 2021, Yu et al., 2022 | This theme reflects service users’ reflections on their experience of the care pathway leading to assessment or involuntary treatment, e.g. whether it was felt that the admission was necessary or avoidable, and whether alternatives were available and explored. Some reported a lack of access or availability of services that would be less restrictive than hospital admissions. Others did not feel they received the right support from their GP, community or early intervention service despite seeking help, leading to symptoms getting worse, and resulting in admission or readmission. For example, police involvement and detention followed due to inadequate responses from A&E and health emergency call centres (Bendelow et al., 2019), or mental health crises escalated otherwise due to either lack of sufficient support from social services (Solanki et al., 2023), insufficient knowledge of low-threshold services by primary care physicians (Wormdahl et al., 2021), or insufficient capacity of specialist outpatients services to support those with serious mental health issues (Wormdahl et al., 2021). | | When seeking help from other sources out of hours, our interviewees were often advised to present to an emergency department or to call 999. Rejection by hospital staff or inadequate responses to help-seeking often appeared to have escalated desperate behaviour in many of these accounts and culminated in situations that resulted in s136 detention. Barry, aged 25, described how being told by emergency department staff that asking for help because he felt suicidal was inappropriate and a waste of their time prompted him to drive to a notorious suicide spot where he was eventually detained.  Bendelow et al., 2019  The impact of admission and diagnosis. I want people around me to actually recognise it [illness] first and go for early intervention rather than ever having to walk into an archaic situation where men and women are insane. That was barbaric. (Female 217 Low)  Smyth et al., 2021  This included experiences of seeking help when vulnerable and receiving unhelpful responses from primary care and social services.  Months ago when I approached my general practitioner1 and I said to him that I was feeling depressed, I should have got help then. Rather than when it becomes too late, so that’s where I feel I’ve been let down :: : , I think, at that time, I feel he should have taken it more seriously. (Participant 7)  Solanki et al., 2023  Many participants with lived experience and carers said that GPs often relied on medication as the main treatment option for people with SMI. In addition, participants from all stakeholder groups, including GPs, mentioned that GPs had limited knowledge of the available low-threshold services in primary mental health care. Several participants with lived experience and carers stated that GPs did not have suﬃcient time to conduct comprehensive assessments of their needs and match them with available services. This was also mentioned in relation to other services, such as when specialist outpatient mental health services only allocated a 1-h follow-up each week; according to participants with lived experience and carers, this was insuﬃcient to help someone with SMI who deteriorated.  Wormdahl et al., 2021 |
