## Supplementary material for "A qualitative meta-synthesis of service users’ and carers’ experiences of assessment and involuntary hospital admissions under mental health legislations: a five-year update": Table 8

**Supplementary information, Table 8.** Selected additional evidence for Carer findings.

| **Themes and Sub-themes** | | **References** | **Finding** | **Quotes – all participant names are pseudonyms.** |
| --- | --- | --- | --- | --- |
| **1 Emotional impact** | | | | |
| **Negative emotions** | | 4 studies  Dixon 2022  Jaeger 2019  Ranieri 2018  Wyder 2018 | Recent papers carers also reported fear about service users’ symptoms and behaviour prior to detention, high stress and hypervigilance in the build-up to a service user’s crisis, and frustration when health professionals ignored or “*failed to grasp the gravity of the service user’s illness”*. Carers felt bad about initiating coercive measures and found assessment distressing, and admission traumatizing.  The previous review reported pervasive stress and distress about the deterioration in the health of their family member and the struggle to find help while looking after someone who was unwell. Anger and frustration about the lack of information and help not being available until the health of their family member had deteriorated to the extent that detention was necessary.  Anxiety and fear for the safety of the person they cared for during detention; about how their family member would cope in hospital; of being blamed by their family member or by health professionals; that the service user’s health may deteriorate again following discharge; and of prejudice and stigma.  Recent papers carers also reported fear about service users’ symptoms and behaviour prior to detention, high stress and hypervigilance in the build-up to a service user’s crisis; and frustration when health professionals ignored or “failed to grasp the gravity of the service user’s illness”. Carers felt bad about initiating coercive measures and found assessment distressing, and admission traumatizing. | “Families talked about their efforts to engage the patient in treatment, through pressure or empathy and gentle persuasion. They sought help in the professional system. Some initiated involuntary hospitalization and then felt insecure and bad about this step.”  **Jaeger**  “Admission was a distressing and traumatizing experience for caregivers, who felt disheartened at leaving a relative in the hospital against their will.”  **Ranieri**  “The crisis build-up generally was experienced as a difficult and intense time characterized by high stress and chaos. Many described that they had become hypervigilant about their family member’s behaviours.”  **Wyder** |
| **Relief** | | 3 studies  Dixon 2022  Ranieri 2018  Wyder 2018 | The previous review reported that carers experienced relief that the severity of the illness was recognised, that they were believed, that the patient was in a safe place, and to receive some respite and shared responsibility with health services.  Recent papers also reported some carers experiencing some relief when the person they cared for was safe, the risk of harm to self or others had been averted, and carers could let go of some worries and receive information about the illness. Carers were also relieved when someone else initiated detention. | *“...being frank, but it’s a sense of relief when it [detention] happens almost, because, you know, she’s not going to end up doing something horrible to someone or herself. It’s literally that extreme I guess*.” (Participant 5)  **Dixon**  “All, at least initially, experienced a sense of relief when their family member was admitted to hospital under an ITO, as it not only provided an explanation for their loved one’s behaviours, but it also meant that they were going to receive the care they needed. Emma described these feelings as follows: “*It felt like relief that someone else was aware and giving him help*” (Emma, mother, Brisbane).  **Wyder** |
| **Adverse effect on carer wellbeing** | | 4 studies  Dixon 2022  Jaeger 2019  Ranieri 2018  Wyder 2018 | In the previous review this theme included carers’ moral distress from initiating the detention. Carers experienced isolation because of keeping the illness and detention confidential, and from not disclosing their caring responsibilities. Carers reported depression and suicidal thoughts as they struggled to prevent health crises in households with multiple and complex needs.  Recent papers also describe carers carrying a “*burden of disease”* for years without the prospect of improvement; personal uncertainty, disagreements with other family members about the right course of action, and differences between what the person they care for wanted and what was in their best interests. After admission, some carers remained fearful about what would happen if their relative refused treatment or for their safety amongst other inpatients. | “Most of the relatives described feelings of helplessness and talked about their burden of disease, which they often had faced for years without the prospect of change for the better.”  **Jaeger**  “However, in cases where Nearest Relatives felt unsure about the need for treatment, they voiced acute feelings of guilt. For example:  *The trouble is, being that close, maybe you don’t always do the right thing, which is what you feel guilty about, because you’re not too sure what the hell it is. But what is the right thing? I don’t know. But whether he’s- he has felt let down by me for allowing it [detention] to happen. On a daily basis I get texts and emails. Please get me out, this is awful…I can’t bear it here. I’m going to be attacked by one of these loonies or whatever it is that, you know, he’s texted. And I think I can’t, I can’t help you. There’s nothing I can do. And that’s horrendous.* (Participant 6).  In the above example, the participant’s angst arose from her son’s distress as well as her own uncertainty about what the right course of action was. Additionally, feelings of conflict could be compounded by disagreement amongst family members after detention as to whether detention in hospital was the correct outcome.”  **Dixon**  “When I found that out that a fellow patient was a convicted rapist: Oh my God, I felt more scared for my daughter. I couldn’t sleep at night.”  **Wyder** |
| **2 Availability of support for carers** | | | | |
| **Carers’ own health** | 3 studies  Dixon 2022  Jaeger 2019  Ranieri 2018 | | The previous review included needs arising from carers’ own physical and mental health problems (e.g., anxiety and depression). Some carers attributed these problems to the strain of their caring responsibilities. For others health problems limited their capacity to visit hospital. Needing support and, finding it lacking over successive detentions, carers described a ‘progressive loss of emotional strength’.  Recent papers include carers wanting proactive support from health services for themselves and the whole family, before and after detention, to make sense of the illness, accept their situation, and cope with the stress. | *“We don’t feel that the hospital is particularly proactive in terms of saying we appreciate the pressure you’re under, or the stress you must feel under, and we’ve got this for you. They’ve told us about carer support but that tends to be, in our experience, more to do with practical day-to-day looking after someone after hospital, rather than psychologically, how do you deal with the predicament you find yourself in.”* (Participant 4)  **Dixon**  “An important issue for almost all family members was their own coping with the disease and the current tense situation, and how they tried to take care of themselves (e.g., trying to remember the “real” person with a biography behind the alienated son or daughter, trying to find a healthy distance, seeking support for oneself, struggling for an inner acceptance of the situation, trying to get information and make sense of the disease).”  **Jaeger**  *“If the family could be more involved . . . it would be a more well-rounded approach to recovery and the whole family could recover then as a result because the whole family is affected by it, not just the service user.* (Jennifer, daughter, Ireland)”  **Ranieri** |
| **Lack of information** | 3 studies  Dixon 2022  Ranieri 2018  Wormdahl 2021 | | In the previous review we reported that prior to detention relatives did not know where to get information, especially if it was their first contact with services. Carers were not always present during detention. Patient confidentiality left some carers feeling they did not have enough information to protect themselves. Staff did not recognise them as partners and feelings of being disregarded and excluded exacerbated carers’ fears for patient well-being.  Recent papers similarly refer to carers lacking information about their role, the illness and how best to help.  There are separate subthemes below about Information sharing and Confidentiality following involuntary admission, and about Recognising carer expertise and Maintaining dialogue with health professionals. | “Nearest Relatives spoke about the need to be offered more information about the specifics of the NR role. This included information both about what the role entailed as well as emotional support both prior to and following the compulsory admission.”  **Dixon**  “Feeling unsupported was a common adversity for caregivers, linked to an excess in responsibility and a lack of support within the family structure, or guidance and protection from the mental health services.”  **Ranieri**  “According to the carers, there was little, if any, service approach or support for them as carers to help them manage these situations.”  **Wormdahl** |
| **Too much responsibility** | 5 studies  Dixon 2022  Jaeger 2019  Ranieri 2018  Wormdahl 2021  Wyder 2018 | | The previous review reported carers being overwhelmed by too much responsibility, isolated from sources of support, and expected to manage situations they were ill-equipped to deal with prior to detention. Help to initiate admission was often needed out of hours when access difficult. When assessment did not lead to admission, carers were fearful about managing risk and reported a lack of service responsibility when aggression and violence were present. Carers’ concern did not diminish during admission. Some carers had multiple caring responsibilities.  Recent papers similarly refer to carers having to manage a family member with deteriorating health and needing to advocate for admission with no guidance from health services. Carers also reported feelings of duty and a lack of support from other family members. Following admission some carers spoke about still feeling like the main carer and remaining vigilant. Some carers resumed 24-hour responsibility following discharge. | “Some family members may feel compelled by societal expectations/obligations in relation to duty, i.e. that families should care for one another, even where the relationship is absent or has become strained. In other instances, relatives made a connection between the emotional bonds between themselves and their family member, and a duty to act on their behalf.”  **Dixon**  “They reported on how they had tried to support their ill family member for a long time in coping with everyday life and with the disease, through close control or by supporting the patient’s desire for independence.”  **Jaeger**  “Feeling unsupported was a common adversity for caregivers, linked to an excess in responsibility and a lack of support within the family structure, or guidance and protection from the mental health services.”  **Ranieri**  “Participants with lived experience and carers especially emphasized loneliness, and many experienced that the stigma around SMI in the wider society heightened the individuals’ loneliness; several participants said the carers became the only social network for the individual.”  **Wormdahl** |
| **3. Carer involvement in decision making and the provision of care** | | | | |
| **Recognising carer expertise** | 4 studies  Dixon  Jaeger 2019  Ranieri 2018  Wyder 2018 | | In the previous carer review we reported that carers had useful knowledge and experience about the relative they care for: when they were well, during previous episodes of illness, and leading up to the latest admission. Carers did not have opportunities to share this or were not listened to by staff. They wanted to be treated as a resource and provide information confidentially so health professionals could make better and more informed treatment decisions.  Recent studies similarly report that carers want their knowledge and experience of caring to be recognised by health professionals and to be included as partners in care, informing treatment decisions and continuing to provide emotional support. | “A major influence on most family members was the experience of a long history of repeated illness episodes with varying degrees of hope and frustration.”  **Jaeger**  “Caregivers expressed a need for sharing information in order to maximize their relative’s treatment in the shape of both informing the services of the service user’s history of illness and circumstances at admission and receiving information to help bring about recovery for the service user and the family as a whole.”  **Ranieri**  “Partnership was not described as needing to be involved in all aspects of care, however it did involve being included in the care so that they were able to continue to provide emotional support to their loved ones.”  **Wyder** |
| **Maintaining dialogue** | 3 studies  Dixon 2022  Jaeger 2019  Wyder 2018 | | In the previous carer review we reported that carers’ expectations for dialogue were often not met. Information was often given during times of chaos and stress. No one talked to young next of kin about detention even when they had witnessed their family member being detained. Carers were frustrated not to be involved in discharge planning.  Similarly, recent studies reported that carers tried to maintain dialogue with hospital staff and participate in treatment and discharge decisions but were often not being heard. In England, even carers formally identified as the Nearest Relative were not always consulted. | “Following the admission of their family member to the hospital, they tried to stay in touch with doctors or to participate in treatment decisions, with mixed results. Some felt rejected by the professionals, and they missed more detailed information about the treatment process, mainly due to a lack of time.”  **Jaeger**  “Many families did not believe that their relative was ready for discharge and that staff did not take into account their concerns. Family members were concerned about their ability to look after their relative. As a result, they felt unheard and invisible.”  **Wyder** |
| **Sharing information** | 3 studies  Dixon 2022  Jaeger 2019  Ranieri 2018  Wyder 2018 | | The previous carer review reported information is often lacking. Legal status (e.g. nearest relative/nominated person) afforded carers different rights to information and involvement which affected their experience. Carers need accessible information about their relative’s illness, medication, and needs; plans for their care and discharge; and about the legal rights and entitlements of patients and carers.    Similarly, recent studies confirm that carers want to maximise treatment and recovery by sharing information and described the importance of receiving information from clinicians about the illness and the service user’s progress. Carers frequently reported the absence of information about inpatient treatment decisions, medications, transfers, or discharge. Carers who received more information about the condition of the person they care for were more conﬁdent in dealing with symptoms after discharge. | “One participant noted that whilst ward staff would speak to them if he initiated contact, in his view, his daughter could ‘*break a leg’* without the ward ringing him up to inform him (Participant 4). Similarly, others spoke about not being informed of major decisions such as their relative being moved to another unit.”  **Dixon**  “In addition, many family members complained that they were not involved in therapy decisions. Some missed getting a basic understanding about the disorder, the treatment, and the proceedings in the ward.”  **Jaeger**  “Caregivers expressed a need for sharing information in order to maximize their relative’s treatment in the shape of both informing the services of the service user’s history of illness and circumstances at admission and receiving information to help bring about recovery for the service user and the family as a whole.”  **Ranieri** |
| **Confidentiality** | 2 studies  Dixon 2022  Ranieri 2018 | | The previous carer review reported that patient confidentiality was a major barrier to carers accessing information and left some carers feeling that they did not have enough information to optimize care or protect themselves.  Again, in recent papers, carers recognised staff were constrained by confidentiality policies but felt that confidentiality prevented them from receiving information that would help them take care of their relatives. Some carers were frustrated they were given no sense of their relative’s progress or informed of major decisions about care following compulsory admission. Some carers thought that confidentiality was being used to inhibit family involvement and was against their rights. | *“I could tell them [information], but they weren’t able to tell me, which I can understand with confidentiality and all that, but actually that’s quite frustrating as a relative when you’re shut out* (Participant 4).”  **Dixon**  “Many caregivers stated that the service user’s right to confidentiality prevented them from receiving information that they felt would help them take care of their relative.”  **Ranieri** |
| **Power dynamics**  **(NEW)** | 3 studies  Dixon 2022  Jaeger 2019  Ranieri 2018 | | A mixed finding of recent carer expressions of power and powerlessness. Many carers felt powerless, but some viewed their relative’s admission as forced by them and some used their relative’s economic dependency as leverage to engage in treatment. Some carers felt distressed that their powers as a Nearest Relative unbalanced their relationship with their family member. | “…the experience of powerlessness was omnipresent.”  **Jaeger**  “Some parents reported that their children’s economic dependency was an efficient leverage to make them engage in treatment.”  **Jaeger**  “Behaviour perceived as coercive at admission was justified by their fear that the service user would self- harm or suffer a further decline in health had their relative not admitted them. Furthermore, caregivers stated that service users did not choose to be admitted, nor did the idea originate from them.”  **Ranieri**  “Nearest Relatives were distressed by the change in power dynamics that the NR brought about; but remained hopeful that these changes were temporary.”  **Dixon** |
| **4. Carer relationships** | | | | |
| **Carer relationships with health professionals** | 4 studies  Dixon 2022  Jaeger 2019  Ranieri 2018  Wyder 2018 | | In the previous review we described carer relationships with healthcare professionals prior to/during detention as unsatisfactory, strained, invalidating, and a terrible battle. Following admission, carers described being disregarded or treated as strangers by staff. Staff did not engage with carers in an effective partnership or acknowledge the impact of detention on the family. Positive relationships with members of staff were infrequently reported but had a powerful impact.  Recent reports were also mixed. Carers wanted to be able to work in partnership with health professionals and appreciated professionals who were kind and respectful. Others experienced staff as lacking empathy, compassion, or time to talk; and failing to listen or take carers seriously. | “They reported their experience that doctors or staff did not have enough time to talk to them and respond to their questions and concerns.”  **Jaeger**  “Caregivers expressed difficulty in gaining medical attention from both the mental health and police services, both of which failed to listen, provide guidance, or recognize the need for their relative’s hospitalization.”  **Ranieri**  “Another predominant theme was the importance of working in partnership with health care professionals.”  **Wyder** |
| **Mediation**  **(NEW)** | 3 studies  Jaeger 2019  Wyder 2018  Dixon 2022 | | Some carers in recent studies described mediating between the person they care for and health professionals. Carers who did not believe that their family members were safe in hospital felt that they had to actively advocate on their behalf. Carers took an intermediary role when the relatives refused to speak directly to hospital staff. Carers formally identified as Nearest Relative found themselves torn between honest communication with health professionals and loyalty to their family members’ wishes. | “Others took on a mediating role between their family member who refused to speak and the professionals by translating the perspectives of each side to the other, in order to strengthen cooperation between the two.”  **Jaeger**  “*With my mum, because my mum’s very paranoid...whenever I go to meetings and things, she says, don’t say anything to the doctors to make me stay here longer...And I would say it [the Nearest Relative role] puts me in an awkward position, because as much as I hate to see my Mum in hospital, equally I would not want her to come out before she was ready and then have to go in again* (Participant 3).”  **Dixon** |
| **Family relationships** | 4 studies  Dixon 2022  Jaeger 2019  Ranieri 2018  Yu 2022 | | In the previous review, carers described the breakdown of relationships with the people they care for following detention. In many cases service users blamed carers for their detention and felt betrayed. Relationships needed to be renegotiated and trust regained. There was a reduction in contact during admission and geographical distance hampered visits. Young carers’ felt the loss of being with a loved parent or sibling.  Recent studies similarly report relationships between carers and service users breaking down following admission, including rejection, mistrust, losing touch, or cutting off contact from either side. Recent studies also described conflicts between guardians or with other family members.  In England, no carer participant with the role of Nearest Relative thought that the role should be given to anyone outside the family, even if their relationship was not a close one. | “… many citing a breakdown in communication following admission. When admission was led by someone external to the family, families expressed relief over a prevented rupture in their relationship with the service user.”  **Ranieri**  “In some instances, Nearest Relatives identified that a close relationship with the person was not important. For example, when Participant 5 was asked whether the Nearest Relative role had affected his relationship with his sister, he responded: *There isn’t one...I’ll do anything I need to do to help her, but to say there is a relationship – my relationship is with the person she was...* (Participant 5).”  **Dixon**  “Notably, no Nearest Relatives in our sample suggested that the Nearest Relative function should be given to another person outside of the family unit.”  **Dixon**  “They [carers] talked about continued rejection and mistrust of the patient, and how much they were affected.”  **Jaeger**  “Some interviewees reflected on their family relationships. They suspected that this also played a role in the behavior of their ill family member.”  **Jaeger**  “…after the requirements for involuntary admission were strengthened, the burden of care on guardians increased in the process of seeking cooperation from the police. This increased burden can lead to conflicts between legal guardians who attempt to have patients committed and patients who reject their attempts, at times leading to severe acts of patient violence. The worst situation occurred in which family members cut off contact with a patient who had assaulted them during the admission process:  *I thought, “I’m going to have to let my daughter hit me today,” and then, “I guess I should call [the police] now.” That is why I let her hit me. After that, she was trying to run away because the police were coming, so the police officers immediately restrained her, and we went to the hospital. I had bruises all over the place, not just on my knees.* (F1)”  **Yu** |
| **5. Quality of care** | | | | |
| **Leading up to detention** | 4 studies  Dixon 2022  Ranieri 2018  Wormdahl 2021  Yu 2022 | | In the previous Carer review we reported that services were not proactive or sufficiently responsive to the needs of patients and carers; not recognising the severity of the patient's illness and not intervening until detention inevitable.  Recent papers also described carers’ frustration with insufficient community services which, instead of acting proactively in the early phases of the illness, dismissed carer concerns until the service user was acutely unwell. | *“I felt that we did not get help fast enough when the crisis appeared. It was like there was nothing between no help and coercion. My wife had to become very, very, ill before they understood the severity of her condition, and then it ended in an involuntary admission. I believe that if the doctor had taken better time to hear us out and gotten more insight into her problems, she could have gotten better help and recovery before she got so ill that she had to be involuntarily admitted.* (Carer, spouse)”  **Wormdahl**  “Those who believed that staff should been more proactive in using compulsory powers also felt that their relatives’ mental health problems could have been ‘*nipped in the bud’* (Participant 2) through detention at an earlier stage. In doing so, they expressed concerns that an undue focus on the service users’ rights and wishes may have acted against their best interests in the long run.”  **Dixon**  “Due to the strengthening of admission requirements based on family consent, persons in need of psychiatric treatment were left unattended in the community without receiving appropriate treatment in a timely manner. Lack of an alternative measure to inpatient treatment for persons with mental illness in the community meant that they had no choice but to be involuntarily committed by the police if their symptoms worsened.”  **Yu** |
| **Detention process** | 4 studies  Bendelow 2019  Ranieri 2018  Wyder 2018  Yu 2022 | | The previous review reported that by the time assessment and detention occurred, many carers already felt let down by services. Some carers described detention processes as inappropriate or heavy-handed but there was one was example of a police officer de-escalating the situation with child.  Recent reports are also mixed. Carers appreciated a street triage initiative and service users being treated with respect. Carers also reported a lack of community alternatives to inpatient admission, concern about how involuntary admission was managed. | “The street triage team also received praise and thanks from a number of relatives and carers for their swift, effective and compassionate interventions.”  **Bendelow**  “Caregivers’ accounts of treatment at admission tended to vary. Some caregivers reported that their relative was treated fairly and respectfully during the admission. Admissions viewed in a positive light were characterized by the staff ’s generosity, patience, and kindness toward the service user. Those viewed negatively were described as clinical and devoid of compassion.”  **Ranieri**  “Lack of an alternative measure to inpatient treatment for persons with mental illness in the community meant that they had no choice but to be involuntarily committed by the police if their symptoms worsened. In this situation, admissions by legal guardians have been replaced with administrative admissions enforced by the police under local government approval.”  **Yu** |
| **Care in hospital** | 3 studies  Jaeger 2019  Ranieri 2018  Wyder 2018 | | The previous review reported carer distress about low-quality care in hospital, security measures, the amount of medication, and lack of meaningful recovery.    Recent studies described some carers being intimidated by the ward environment and overwhelmed by the levels of distress witnessed. Carers wanted to know that their relatives were safe and receiving care, but some did not believe that the hospital met their relative’s needs and did not trust the treatment provided e.g. medication only. | “The main motives for refusal [of medication] according to patients and family members were different conceptualizations of what was the problem to be treated. Some patients did not think they had a problem at all. Some thought they had other problems and medication treatment was not the adequate therapy. They did not feel understood by professional helpers.”  **Jaeger**  “Others felt that the quality of the care their relative received and caregiver interaction was poorer if their relative was admitted to a public rather than private hospital.”  **Ranieri**  “… the vast majority (n = 14) did not believe that the hospital met their relative’s needs. Many of the families in the study were critical of the strong focus on medication as the only form of treatment. They were also concerned that their relative did not have any opportunity to talk about their experiences.”  **Wyder** |
| **Discharge processes** | 5 studies  Dixon 2022  Jaeger 2019  Ranieri 2018  Wyder 2018  Yu 2022 | | The previous review reported carer dissatisfaction with having to care for service users discharged at short notice or while still very unwell.  Recent studies also reported that most carers are informed about discharge at the last minute, when neither the service user nor family are ready. Carers are distressed by the difficulty of trying to obtain outpatient aftercare for their discharged relative and some want them to be readmitted or taken to another mental health care facility. Carers would like crisis planning and advance directives to prepare for possible future readmissions. | “In several cases, participants stated that they had not been informed that leave or discharge was due to be granted and had been given no information about support arrangements.”  **Dixon**  “As a consequence of the changed legal situation, some patients had been discharged prematurely when they were no longer at risk of harming themselves or others. The families found this an additional burden, especially as they had hoped to hand over responsibility and get help.”  **Jaeger**  “Many caregivers, however, appraised this time as difficult due to the service user’s continued impairment following discharge.”  **Ranieri**  “Finally, caregivers highlighted the possibility of future admissions. In doing so, some stated a desire for crisis planning and helping service users feel less nervous about admission by letting them familiarize themselves more comfortably with hospital settings and choose their own admitting preferences by means of advanced directives. These ideas are proposed as ways to prepare both service users and families for possible subsequent hospital readmissions.”  **Ranieri**  “It was equally important for participants to have information about the timing of the discharge of their relative. This time often represented another important transition point where families described the need to reconnect. It was also a time where families needed to balance facilitating independence with providing the necessary support to their family members. But, for this to occur it was important that families felt prepared for the discharge and had processes and supports in place. While a structured and planned discharge process appeared to facilitate their relative’s reintegration into the community, the majority of the participants (n = 14) described the discharge process as ad hoc, where they were only informed at the last minute that their relative was going to be discharged.”  **Wyder**  “Most psychiatric inpatients can be discharged after their legal guardians have paid their hospital bills and arranged a place for them to stay when they leave the hospital. Even if the Mental Health Review Board orders long-term inpatients to be discharged, family members often want the patient to be readmitted or taken to another mental health care facility, rather than living together.”  **Yu** |
| **Impact of coercion**  **(NEW)** | 4 studies  Dixon 2022  Jaeger 2019  Ranieri 2018  Wormdahl 2021 | | Service user experiences of being detained roughly or in public or being subjected to coercive measures during admission and treatment in hospital, have consequential impacts on carers too. Carers in recent studies reported difficulties arising from their relatives’ subsequent fear and unwillingness to seek help and engage with services, such as withdrawing and keeping their distance from carers; and being guarded and masking their symptoms during assessments. In one study carers acknowledged that coercion might lead to short-term improvement in hospital but regarded the effect as unsustainable and ultimately negative. | “*And so, I was raising concerns continually with the mental health team saying, look he’s not well and in the end, I said to the mental health team guy, I said, you need to stay with him for more than ten minutes, because you don’t – he puts up – he’s very guarded and he puts up that he’s fine and doesn’t need any help* (Participant 2).”  **Dixon**  “Caregivers often stated that service users masked their symptoms during medical assessments to avoid admission.”  **Ranieri**  *“Participants with lived experience and carers discussed how some individuals with severe mental illness withdrew from services because they had experienced former admissions as traumatic. Among other things, they talked about being roughly handled, and often the police had been involved. When this happened in public, the participants experienced additional strain and stigma. Some said that the services were not tailored to help people overcome this fear around receiving services.”*  **Wormdahl** |
