## Supplementary material for "A qualitative meta-synthesis of service users’ and carers’ experiences of assessment and involuntary hospital admissions under mental health legislations: a five-year update": GRADE-CERQual protocol

### GRADE Protocol v02: Certainty of evidence for each outcome measure

**Method:** We applied a version of the Grading of Recommendations Assessment, Development and Evaluation (GRADE) system (Guyatt et al., 2008), adapted to assess confidence in findings from qualitative evidence synthesis (CERQual, Lewin et al., 2015). Two people independently assessed the certainty of evidence for each research question and met to resolve any inconsistencies.

Each GRADE-CERQual domain was assigned one of the four ratings below, as per NICE Guidelines (N216 supplement – insert reference):

1. **None or very minor concerns**: unlikely to reduce confidence in the review finding.
2. **Minor concerns**: may reduce confidence in the review finding.
3. **Moderate concerns**: will probably reduce confidence in the review finding.
4. **Serious concerns**: very likely to reduce confidence in the review finding.

The four GRADE-CERQual domains were defined and operationalized in the following way, according to Lewin et al. (2015).

1. **Methodological limitations**: The extent to which there are problems in the design or conduct of the primary studies that contributed evidence to a review finding. For this item, ratings differing from ‘None or very minor concerns’ were defined as follows. *Serious concerns*: 50% or more of the studies were of low quality (score less than 4)*. *Moderate concerns*: 50% or more of the contributing studies were moderate or low quality (50% with score of less than 7). *Minor concerns*: some concerns exist with individual studies but not meeting criteria for moderate or serious concern defined above.

- *Go to Study Information workbook in the MHA Review/GRADE/Ratings folder.*
- *Use filter for the finding being evaluated.*
- *See CASP quality rating column for High, Medium, Low.*
- *Check number of each, and match against criteria above.*

2. **Relevance**: The extent to which the body of evidence from the primary studies supporting a review finding is applicable to the context (perspective or population, phenomenon of interest, setting) specified in the review question. For this item, assessors evaluated indirectness, partiality and uncertainty of evidence, defined as follows. *Indirectness*: if a review domain, e.g. setting, perspective or population has been substituted by another in the primary study. E.g. experience of coercion of service users who are in hospital voluntarily, not involuntarily). *Partiality*: studies identify only some of the relevant review domains (e.g. data on one setting only, when review interest is broader). *Uncertainty*: the extent to which the focus of the included studies reflects the phenomenon of interest is uncertain, because of deficiencies in the reported details of the population, intervention, or settings. E.g. a study reports on involuntary treatment experiences, but also treatment experiences in general, and assessors are unsure to what extent results relate to involuntary experiences. Detailed justification for any rating reductions in this domain are described in Table X.

- *Go to Study Information workbook in the MHA Review/GRADE/Ratings folder.*
- *Use filter for the finding being evaluated.*
- *See and assess the information in the following columns for each finding: Study place, Study aims, Study focus, Study setting, Diagnoses of patients.*

3. **Coherence**: The extent to which the review finding is well grounded in data from the contributing primary studies and provides a convincing explanation for the patterns found in these data. Review findings are sometimes challenged by outlying, contrasting, or even disconfirming data from the primary studies that do not support or that directly challenge the main finding. Domain ratings were reduced if: a) data were considered too ‘thin’ (*Is it clear what the context, meaning of the finding is? E.g. ‘patients experienced discrimination’ only.* i.e. the phenomenon, e.g. coercive practice is described narrowly, as opposed to in richness of context, meaning, and interpretation), b) data also contained evidence disconfirming the finding (*Does data support the sub-theme description, or are there data that point in another direction?*, without potential theoretical explanation, or c) studies with limited samples (*Studies with good quotes, but may only come from a small/not reflective sub-sample? Do items below in blue cover the population of interest?*) were included.

**For a) and b):**

- *Go to the Findings file in the GRADE/Ratings folder.*
- *For each finding, evaluate the Quotes/quote summaries column.*
- *Consult NVivo Master file if information is insufficient.*

**For c):**

- *Go to Study Information workbook in the MHA Review/GRADE/Ratings folder.*
- *Use filter for the finding being evaluated.*
- *See and assess the information in the following columns for each finding: Number patients in sample, Gender patients, Age patients, Ethnicity.*

4. **Adequacy of data**: An overall determination of the degree of richness and quantity of data supporting a review finding. Domain ratings were reduced a) if there were insufficient number of studies reporting on the finding, or b) there was insufficient data in primary studies supporting the finding.

- *Go to the Findings file in the GRADE/Ratings folder.*
- *For each finding, a) evaluate the number of studies contributing to the finding in the References column.*
- *For each finding, b) evaluate richness of data based on Quotes/quote summaries column.*

**Overview of GRADE CERQUAL assessment:** Overall confidence in evidence supporting each finding has been graded as one of the four categories below. For each finding, ‘High’ confidence in evidence was assumed, unless the following reductions were made:

- If there were two or more domains rated as ‘Minor concerns’: assessors considered downgrading confidence by one category, documenting the justification for their decision.
- If there was any domain rated as ‘Moderate concerns’: assessors considered downgrading confidence by one category, documenting the justification for their decision.
- If there was any domain rated as ‘Serious concerns’: assessors downgraded confidence by one category, or considered downgrading by two (justifying reasons if so).

Thus, final confidence ratings were one of the following categories:

1. **High.** It is highly likely that the review finding is a reasonable representation of the phenomenon of interest.
2. **Moderate.** It is likely that the review finding is a reasonable representation of the phenomenon of interest.
3. **Low.** It is possible that the review finding is a reasonable representation of the phenomenon of interest
4. **Very low.** It is unclear whether the review finding is a reasonable representation of the phenomenon of interest

*Quality ratings here and henceforth are based on the CASP tool (Table X.) using the same rating system as in the previous review Akther et al . (2019). Studies scoring 7 or higher were rated as ‘high’, 4 or higher as ‘moderate’, and below 4 as ‘low’ quality.

**Resolving disagreements:** Ratings were carried out by two study members independently (RS, NA, and/or GB). All domain ratings and confidence ratings that were rated differently were discussed until agreement was reached. If reaching agreement was inconclusive, a senior author (BLE, UF and/or AS) was consulted until agreement was reached.

**Table X.** CERQual confidence in evidence supporting findings (template example only: to update from GRADE assessors agreed ratings)

| Study information | Description of review finding | CERQual Quality Assessment | | | | |
| --- | --- | --- | --- | --- | --- | --- |
|  |  | Methodologi-cal limitations | Relevance of evidence | Coherence of finding | Adequacy of data | Overall confidence |
| **Theme 1a): Emotional impact: Acceptance** | | | | | | |

| Study information | Description of review finding | CERQual Quality Assessment | | | | |
| --- | --- | --- | --- | --- | --- | --- |
|  |  | Methodologi-cal limitations | Relevance of evidence | Coherence of finding | Adequacy of data | Overall confidence |
| **Theme 1b): Emotional impact: Impact of detention** | | | | | | |
